# Associations Between Major Psychiatric Disorder Polygenic Risk Scores and Blood-Based Markers in UK Biobank

**DOI:** 10.1101/2020.11.06.20227066

**Authors:** Michael D.E. Sewell, Xueyi Shen, Lorena Jiménez-Sánchez, Amelia J. Edmondson-Stait, Claire Green, Mark J. Adams, Andrew M. McIntosh, Donald M. Lyall, Heather C. Whalley, Stephen M. Lawrie

## Abstract

**Background:** Major depressive disorder (MDD), schizophrenia (SCZ), and bipolar disorder (BD) have both shared and discrete genetic risk factors and abnormalities in blood-based measures of inflammation and blood-brain barrier (BBB) permeability. The relationships between such genetic architectures and blood-based markers are however unclear. We investigated relationships between polygenic risk scores for these disorders and peripheral biomarkers in the UK Biobank cohort.

**Methods:** We calculated polygenic risk scores (PRS) for samples of *n* = 367,329 (MDD PRS), *n* = 366,465 (SCZ PRS), and *n* = 366,383 (BD PRS) individuals from the UK Biobank cohort. We examined associations between each disorder PRS and 62 blood markers, using two generalized linear regression models: ‘minimally adjusted’ controlling for variables including age and sex, and ‘fully adjusted’ including additional lifestyle covariates such as alcohol and smoking status.

**Results:** 12/62, 13/62 and 9/62 peripheral markers were significantly associated with MDD, SCZ and BD PRS respectively for both models. Most associations were disorder PRS-specific, including several immune-related markers for MDD and SCZ. We also identified several BBB-permeable marker associations, including vitamin D for all three disorder PRS, IGF-1 and triglycerides for MDD PRS, testosterone for SCZ PRS, and HDL cholesterol for BD PRS.

**Conclusions:** This study suggests that MDD, SCZ and BD have shared and distinct peripheral markers associated with disorder-specific genetic risk. The results implicate BBB permeability disruptions in all three disorders and inflammatory dysfunction in MDD and SCZ, and enrich our understanding of potential underlying pathophysiological mechanisms in major psychiatric disorders.

## Introduction

Major depressive disorder (MDD), schizophrenia (SCZ), and bipolar disorder (BD) are potentially debilitating psychiatric disorders with significant morbidity and economic impact worldwide (1). Despite this, the pathogenesis of these disorders remains elusive (2), with diagnosis reliant upon symptom presentation (3). All three disorders are heritable and of polygenic nature (4), with strong evidence suggesting they share some genetic risk (5, 6, 7, 8) and phenotypic traits (9, 10, 11). Conversely, other studies have identified disorder-specific genetic associations (12, 13, 14) and neurobiological characteristics (15, 16, 17, 18), alluding to variations in the genetic pathophysiology between conditions.

There is also compelling evidence demonstrating that MDD, SCZ and BD are accompanied by genetically and environmentally influenced alterations in peripheral blood-based marker levels (19, 20, 21). Similarities and differences in the nature of these, such as an elevated inflammatory response, have been observed in patients across the three disorders (22, 23, 24, 25). Although these peripheral perturbations, including in inflammatory measures, have also differentiated cases from healthy controls (22, 26, 27), their potential role in disorder pathophysiology has not yet been ascertained, and currently there are no markers available that can predict the risk of any disorder (28). Conflicting data concerning associations in case-control studies, such as those investigating the relationship between inflammatory marker C-reactive protein (CRP) and schizophrenia (29), may be attributable to limitations such as selection bias, small sample sizes, and illness related confounding factors such as medication.

Progress in identifying blood-based markers for MDD, SCZ and BD may be advanced by examining peripheral profiles of large samples of individuals according to their genetic predisposition for each disorder, as denoted by their polygenic risk score (PRS) (30). PRS are constructed by summing the number of risk alleles, weighted by effect sizes derived from genome-wide association studies (GWAS) of independent samples (31). Previous work by our group and others have demonstrated neurobiological associations for individuals with higher PRS for all three disorders (32, 33, 34, 35). Here, we extend this to compare blood-based markers.

We have employed a data-driven exploratory approach to examine relationships between PRS for all three disorders (MDD, SCZ, BD) with peripheral markers using data from the UK Biobank (UKB) cohort in a total of over 500,000 participants (36). We were primarily interested in identifying associations with markers that traverse the blood-brain barrier (BBB), due to repeated observations of BBB impairment in patients with MDD, SCZ and BD (37). This is in keeping with studies showing altered brain or peripheral levels of BBB-permeable markers including glucose, high-density lipoprotein (HDL) cholesterol and triglycerides in patients (38, 39, 40, 41). Additionally, it is increasingly clear that inflammatory and immune dysregulation are characteristics of MDD, SCZ and BD (25, 42), which suggests that increased PRS may also be associated with deviations in inflammatory and immune system markers. We therefore hypothesized that higher disorder PRS for MDD, SCZ and BD would be related to the levels of these markers. However, we did not restrict our analysis to BBB-permeable, inflammatory and immune system components, as the high statistical power generated by our large population-based sample could also yield novel associations with other markers not yet implicated in disorder pathology.

## Methods and Materials

### UK Biobank

The UKB study involved 502,617 community-based individuals recruited between 2006 and 2010 in the United Kingdom (https://www.ukbiobank.ac.uk/) (36, 43). UKB received ethical approval from the research ethics committee (REC reference 11/NW/0382), under application #4844, with all participants providing written consent.

### Study Population

The study was conducted using the latest release of UKB blood marker data including 502,536 participants (*n*_*male*_ *=* 229,134, *n*_*female*_ = 273,402, mean age at time of blood sampling = 56.53 ± 8.10 years, age range = 37-73 years). We firstly removed participants who had supplied no biomarker data (*n =* 9833). Subsequently, as part of the genetic quality checking procedure described in Howard *et al* (44) we performed prior to PRS generation, we excluded participants who had non-white British ancestry based on self-reported ethnicity, gender mismatch, relatedness using kinship coefficients calculated from the KING toolset (r > .044), and genotype missingness greater than 2%. For both SCZ and BD PRS analyses, individual identifier information was not available from the Psychiatric Genomics Consortium (PGC) GWAS summary statistics used to compute SCZ and BD PRS (45, 46). To minimize any potential overlap between UKB and PGC SCZ and BD cohorts, we excluded subjects who had received SCZ or BD diagnoses, identified through self-declaration and hospital records, and defined using international classification of disease (ICD10) codes F20-F29 for SCZ, and F30-F31 for BD (11^th^ revision) (http://biobank.ctsu.ox.ac.uk/crystal/field.cgi?id=41202). The same measures were not implemented for MDD PRS analysis as we were able to directly identify UKB subjects who had also participated in MDD PGC studies in order for them to be removed in the current analysis (See Genotyping and PRS Generation in Methods). This gave us final sample sizes of *n =* 367,329 for MDD PRS, *n =* 366,465 for SCZ PRS and *n =* 366,383 for BD PRS analyses respectively (see Supplementary Figure S1).

### Genotyping and PRS Generation

Genotyping of 488,377 blood samples from UKB participants was performed using either the UK BiLEVE http://biobank.ctsu.ox.ac.uk/crystal/refer.cgi?id=149600 or UKB Axiom arrays http://biobank.ctsu.ox.ac.uk/crystal/refer.cgi?id=149601 (43). Further information about the quality control procedures applied to participant genomic data are described in http://www.ukbiobank.ac.uk/scientists-3/genetic-data, https://www.ukbiobank.ac.uk/wp-content/uploads/2014/04/UKBiobank_genotyping_QC_documentation-web-1.pdf, Hageneaars *et al* (47) and Bycroft *et al* (43).

Prior to MDD PRS derivation, we removed any subjects who had participated in the UKB study from the MDD GWAS summary statistics used to construct MDD PRS (48, 49). Individual PRS for all disorders at five *p* value thresholds (.01, .05, .1, .5 and 1.0) were computed using PRSice software (50) and PLINK (version 1.9; https://www.cog-genomics.org/plink/1.9/), by summing risk alleles weighted on the strength of their association with a certain trait. Association strength has previously been estimated through GWAS conducted by PGC working groups (https://www.med.unc.edu/pgc/pgc-workgroups/). In this manuscript, an SNP inclusion threshold was applied at a significance level of p ≤ .5, in line with previous work showing this threshold is the most predictive of case-control status (45, 51, 52, 53), containing 141,802 SNPs for MDD, 138,505 SNPs for SCZ and 145,975 SNPs for BD after clump-based linkage-disequilibrium (LD) pruning (*R*^2^ = .25, 250-kb window). In addition, SNPs with a minor allele frequency of less than 1%, Hardy-Weinberg equilibrium (p < 1×10^−6^) or variant call rate (< 98%) were removed prior to analysis. The numbers of unpruned and pruned SNPs, and results for other SNP inclusion thresholds p ≤ .01, p ≤ .05, p ≤ .1, and p ≤ 1.0 are reported in Tables S1 and S11-34 in the Supplement.

### Blood-Based Markers

UKB performed quality checking procedures for blood sampling data, including assays described in https://biobank.ndph.ox.ac.uk/showcase/showcase/docs/biomarker_issues.pdf https://biobank.ndph.ox.ac.uk/showcase/showcase/docs/serum_biochemistry.pdf http://biobank.ndph.ox.ac.uk/showcase/showcase/docs/haematology.pdf and https://www.ukbiobank.ac.uk/wp-content/uploads/2018/11/BCM023_ukb_biomarker_panel_website_v1.0-Aug-2015-edit-2018.pdf. Details of the raw data available concerning 61 peripheral markers and how they were subdivided into six groups: ‘BBB-permeable’, and five non-BBB-permeable groups: ‘inflammatory and hematological’, ‘liver’, ‘renal’, ‘cardiovascular’ and ‘other’, are reported in the Supplement (Supplementary Methods, Table S2). In addition, we calculated the neutrophil-to-lymphocyte (NLR) ratio for subjects who supplied neutrophil and lymphocyte counts. Previous studies have demonstrated the use of NLR in predicting acute and chronic inflammatory status (54), showing higher levels in patients for all three disorders (55, 56, 57).

Baseline data for each marker were standardized using the ‘scale’ function in R (version 3.6.1; R Foundation, Vienna, Austria; https://cran.r-project.org/bin/windows/base/old/3.6.1/), and subsequently values that were further than five standard deviations away from the mean standardized value (“zero”) were removed (see Table S3 in the Supplement).

### Statistics

Statistical analysis was performed using R (version 3.6.1) in a Linux environment. To investigate associations between disorder PRS at each SNP inclusion threshold and blood markers, we employed generalized linear regression ‘minimally adjusted’ (MA) and ‘fully adjusted’ (FA) models (‘glm’ function in R; “stats” package version 3.6.1). The following covariates were included for our MA model: age, age^2^, sex, genotyping array used, UKB assessment center the participant attended for blood sample collection, and 15 genetic principal components (see Supplementary Methods). Our FA model additionally controlled for body mass index (BMI), alcohol and smoking status, to account for previous associations identified between all three disorders and these covariates (58, 59, 60, 61, 62, 63) (see Supplementary Methods). This approach enabled us to distinguish biomarker associations attributed to a higher disorder genetic risk from those more likely related to these particular lifestyle factors. We also performed an additional sensitivity analysis for MDD PRS, by testing for peripheral marker associations for our UKB cohort excluding participants who had received an MDD diagnosis (ICD10 classification codes F32-F39, 11^th^ revision, http://biobank.ctsu.ox.ac.uk/crystal/field.cgi?id=41202), (*n* = 10,518) (see Supplementary Figure S1). Multiple testing correction was applied to all tests conducted across all traits at each individual SNP inclusion threshold, using the Bonferroni multiple comparison test (64), utilizing the ‘p.adjust’ function in R (“stats” package version 3.6.1). Significant associations were determined using a threshold of *p*_corr_ < .05. Associations are reported as standardized Beta (ß) values throughout this manuscript.

## Results

### Associations Between MDD PRS and Peripheral Markers

We observed significant associations between MDD PRS and 7/8 BBB-permeable markers in our MA model, although only 3/8 were robust to controlling for lifestyle factors (see Figure 1 Tables S4 and S7 in the Supplement). These were: insulin-like growth factor 1 (IGF-1) (MA: ß = -.016, *p*_corr_ = 6.70×10^−22^, FA: ß = -.010, *p*_corr_ = 2.98×10^−7^), triglycerides (MA: ß = .023, *p*_corr_ = 7.28×10^−44^, FA: ß = .010, *p*_corr_ = 5.83×10^−9^) and vitamin D (MA: ß = -.019, *p*_corr_ = 1.22×10^−25^, FA: ß = -.010, *p*_corr_ = 1.68×10^−7^).

**Figure 1.**
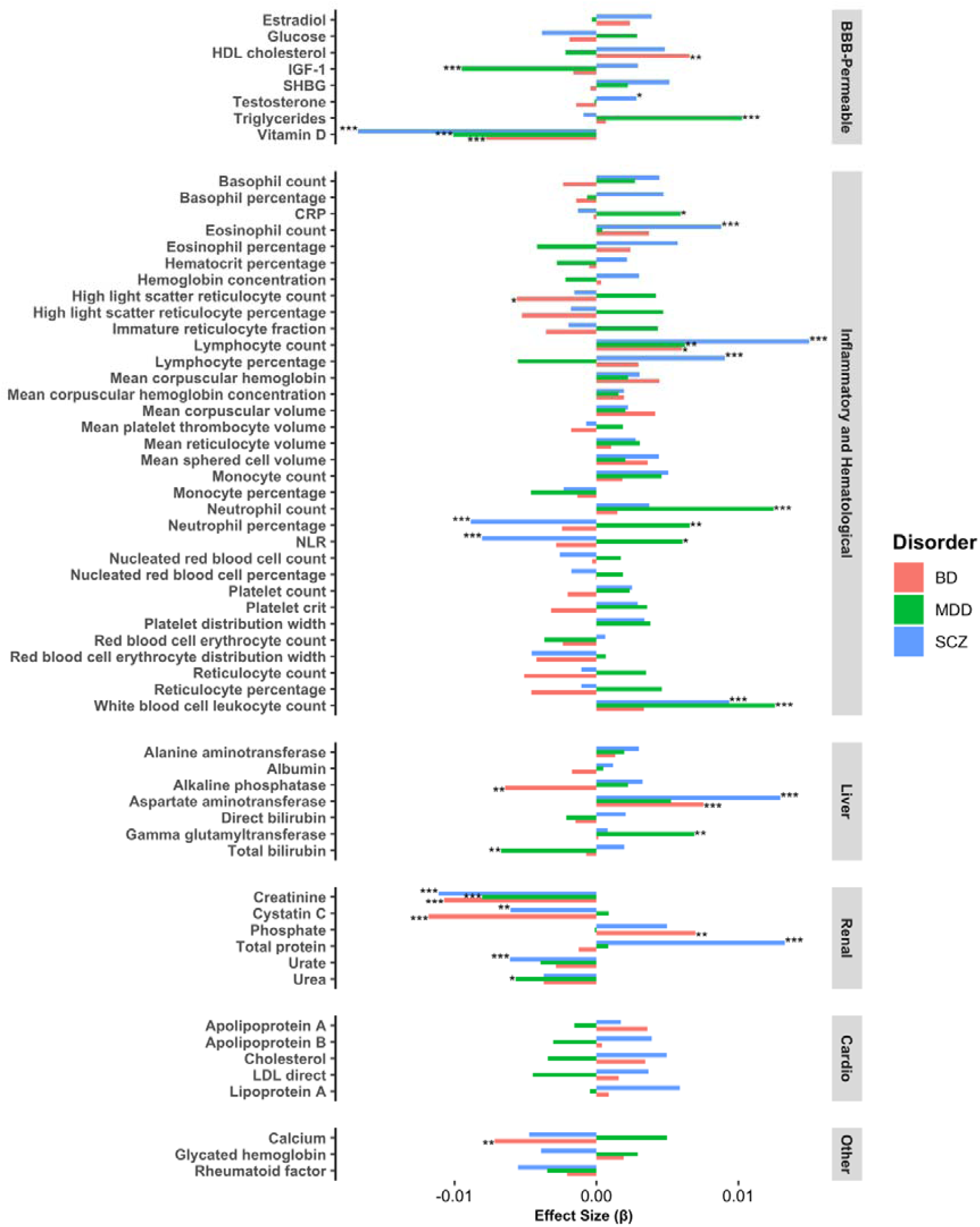
Standardized Effect Sizes (ß) for fully adjusted blood marker associations with each disorder polygenic risk score at threshold ≤ .5. BBB, blood-brain barrier; BD, bipolar disorder; Cardio, cardiovascular; CRP, C-reactive protein; HDL, high-density lipoprotein; IGF-1, insulin-like growth factor 1; LDL, low-density lipoprotein; MDD, major depressive disorder; NLR, neutrophil-to-lymphocyte ratio; Other, other non-BBB-permeable markers; SCZ, schizophrenia; SHBG, sex hormone binding globulin. ***p corr. ≤ .001, **p corr. ≤ .01, *p corr. ≤ .05

We also noted many associations with various non-BBB-permeable markers which survived Bonferroni correction in both models. These included positive associations with several hematological and immune indices: white blood cell (MA: ß = .028, *p*_corr_= 3.67×10^−61^, FA: ß = .013, *p*_corr_ = 1.91×10^−13^), lymphocyte (MA: ß = .019, *p*_corr_ = 2.45×10^−27^, FA: ß = .006, *p*_corr_ = .008) and neutrophil counts (MA: ß = .025, *p*_corr_ = 1.70×10^−47^, FA: ß = .012, *p*_corr_ = 1.13×10^−12^) and neutrophil percentage (MA: ß = .007, *p*_corr_ = 1.02×10^−3^, FA: ß = .007, *p*_corr_ = .006).

Additionally, positive associations were detected for inflammatory markers CRP (MA: ß = .022, *p*_corr_ = 7.19×10^−37^, FA: ß = .006, *p*_corr_ = .012) and NLR (MA: ß = .006, *p*_corr_ = .012, FA: ß = .006, *p*_corr_ = .018). Furthermore, associations were identified with liver and renal function markers, with a positive association for gamma glutamyltransferase level (MA: ß = .016, *p*_corr_ = 2.00×10^−21^, FA: ß = .007, *p*_corr_ = 1.20×10^−3^) and negative associations for total bilirubin (MA: ß = -.014, *p*_corr_ = 5.28×10^−15^, FA: ß = -.007, *p*_corr_ = 2.44×10^−3^) and creatinine (MA: ß = -.008, *p*_corr_ = 1.71×10^−6^ FA: ß = -.008, *p*_corr_ = 2.30×10^−7^),. All of these associations continued to be significant, with the nature of their effect sizes unchanged, after conducting a sensitivity analysis in which we excluded MDD patients from our sample, with the exception of CRP and NLR (see Table S10 in the Supplement).

### Associations Between SCZ PRS and Peripheral Markers

We initially observed SCZ PRS associations with 6/8 BBB-permeable markers in our MA model, (see Table S5 in the Supplement), but this was reduced to only two markers upon implementing the FA model: testosterone (MA: ß = 4.74×10^−3^, *p*_corr_ = 3.58×10^−7^, FA: ß = 2.81×10^−3^, *p*_corr_ = .027) and vitamin D (MA: ß = -.014, *p*_corr_ = 1.16×10^−12^, FA: ß = -.017, *p*_corr_ = 1.26×10^−19^) (see Figure 1 Table S8 in the Supplement). Associations with hematological and immune parameters were also observed across both models, including positive associations for white blood cell (MA: ß = .010, *p*_corr_ = 1.46×10^−6^, FA: ß = .009, *p*_corr_ = 1.13×10^−6^), lymphocyte (MA: ß = .015, *p*_corr_ = 4.37×10^−16^, FA: ß = .015, *p*_corr_ = 4.15×10^−17^) and eosinophil (MA: ß = .008, *p*_corr_ = 1.05×10^−4^, FA: ß = .009, *p*_corr_ = 2.64×10^−5^) counts, and lymphocyte percentage (MA: ß = .009, *p*_corr_ = 2.58×10^−5^, FA: ß = .009, *p*_corr_ = 1.15×10^−5^).

In contrast to MDD PRS, a negative association was detected for neutrophil percentage (MA: ß = -.008, *p*_corr_ = 1.10×10^−4^, FA: ß = -.009, *p*_corr_ = 2.76×10^−5^), and inflammatory marker NLR (MA: ß = -.007, *p*_corr_ = 1.28×10^−3^, FA: ß = -.008, *p*_corr_ = 2.32×10^−4^). Other non-BBB-permeable markers that exhibited positive associations with SCZ PRS included liver function marker aspartate aminotransferase (MA: ß = .010, *p*_corr_ = 7.48×10^−7^, FA: ß = .013, *p*_corr_ = 2.41×10^−12^), and renal function marker total protein (MA: ß = .011, *p*_corr_ = 1.68×10^−7^, FA: ß = .013, *p*_corr_ = 3.56×10^−11^). Negative associations were identified however with renal function markers creatinine (MA: ß = -.014, *p*_corr_ = 4.56×10^−21^, FA: ß = -.011, *p*_corr_ = 3.63×10^−13^), cystatin C (MA: ß = -.010, *p*_corr_ = 1.31×10^−8^, FA: ß = -.006, *p*_corr_ = 4.05×10^−3^) and urate (MA: ß = -.013, *p*_corr_ = 2.45×10^−17^, FA: ß = -.006, *p*_corr_ = 7.08×10^−4^).

### Associations Between BD PRS and Peripheral Markers

Only 19 associations were initially detected with BD PRS for our MA model, only two were with BBB-permeable markers, although a greater proportion of markers remained significant upon adjusting for lifestyle factors compared to MDD and SCZ PRS (see Figure 1 Tables S6 and S9 in the Supplement). Similarly to MDD and SCZ PRS, we observed negative association with vitamin D (MA: ß = -.006, *p*_corr_ = .022, FA: ß = -.008, *p*_corr_ = 5.80×10^−4^). However, a distinct positive association was detected with HDL cholesterol (MA: ß = .010, *p*_corr_ = 4.38×10^−7^, FA: ß = .007, *p*_corr_ = 1.45×10^−3^).

Two non-BBB-permeable marker associations detected for SCZ PRS were also found to occur for BD PRS: liver function marker aspartate aminotransferase (MA: ß = .006, *p*_corr_ = .027, FA: ß = .008, *p*_corr_ = 5.78×10^−4^), and renal function marker cystatin C (MA: ß = -.014, *p*_corr_ = 8.89×10^−16^, FA: ß = -.012, *p*_corr_ = 3.04×10^−13^), along with creatinine (MA: ß = -.012, *p*_corr_ = 2.59×10^−16^, FA: ß = -.011, *p*_corr_ = 2.29×10^−12^) which also was associated with MDD PRS. Other non-BBB-permeable marker associations that were unique to BD PRS included a positive association with renal function marker phosphate (MA: ß = .009, *p*_corr_ = 5.07×10^−5^, FA: ß = .007, *p*_corr_ = .005), and negative associations for hematological parameter high light scatter reticulocyte count (MA: ß = -.010, *p*_corr_ = 4.54×10^−7^, FA: ß = -.006, *p*_corr_ = .027), bone function marker calcium (MA: ß = -.007, *p*_corr_ = .014, FA: ß = -.007, *p*_corr_ = .006), and liver function marker alkaline phosphatase (MA: ß = -.007, *p*_corr_ = 8.67×10^−4^, FA: ß = -.006, *p*_corr_ = .009), which along with vitamin D, can also be indicative of bone health (65). Unlike MDD and SCZ PRS, no significant associations for both MA and FA models were detected between hematological immune indices and BD PRS.

## Discussion

Here, we have demonstrated that MDD, SCZ and BD PRS display varying associations with blood markers across multiple physiological systems. Notably, a large proportion of non-BBB-permeable marker associations for MDD and SCZ PRS were immune and inflammatory related, supporting previous work showing that aberrant immune and inflammatory processes contribute to and possibly underlie these disorders (66). We also report several BBB-permeable marker associations, including vitamin D for all three disorders, IGF-1 and triglycerides for MDD PRS, testosterone for SCZ PRS, and HDL cholesterol for BD PRS.

In keeping with their shared polygenic architecture, we reported some overlapping peripheral traits across all three disorder PRS, notably negative associations with renal marker creatinine, a by-product of creatine metabolism (67), and vitamin D. Lower vitamin D levels (68, 69, 70) and abnormal creatine/creatinine levels or metabolism (71, 72, 73) have been previously reported in cases for all three disorders. However, the majority of associations detected appear to be disorder-specific. This important finding highlights differences in peripheral markers of genetic pathophysiology, and relevance of considering each disorder separately when attempting to identify risk biomarkers.

Moreover, we found several associations with MDD and SCZ PRS that did not mirror those of smaller-scale patient studies, notably lymphocyte count (74, 75) and IGF-1 (76) for MDD. The positive associations we observed with both lymphocyte and neutrophil counts point to elevated chronic inflammation, previously implicated in MDD pathogenesis (77), in individuals with higher MDD PRS. Whilst neutrophil activity is thought to be critical for both acute and chronic inflammation (78), roles have also been proposed for certain lymphocyte subpopulations that secrete pro-inflammatory cytokines such as interleukin-17 (IL-17) and interferon-γ in systemic inflammation (79). Several of these T-lymphocyte-released cytokines, notably IL-17, are thought to disrupt BBB integrity in other disease models (80, 81), potentially also predisposing or precipitating MDD onset. Additionally, inflammation may account for associations identified with non-immune cell parameters, such as gamma glutamyltransferase and total bilirubin. These classical liver function indicators also display associations with chronic inflammation markers that resemble those reported in our study (82, 83).

Elevated systemic inflammation may additionally account for the negative association between MDD PRS and with IGF-1, a marker shown to exert anti-inflammatory effects (76, 84), with lower brain levels in several *in vivo* models associated with a MDD phenotype (85, 86). Chronic inflammation could also explain the insignificant associations with acute inflammatory markers CRP and NLR in our FA model upon patient exclusion. Although both CRP and NLR are considered reliable indicators of inflammatory status, they may be more strongly associated with acute inflammation (87). Our sensitivity analysis results could therefore suggest that increased levels of these markers, and associated acute inflammation seen in MDD patients (57, 88) are downstream of disease onset.

However, we noted a negative association between NLR and increased genetic risk for SCZ, another condition linked to chronic inflammation (89). Reduced inflammation in subjects with higher SCZ PRS was corroborated by the negative association with CRP in our MA model, which may have become insignificant upon lifestyle factor corrections due to its strong association with obesity (90), a common comorbidity of SCZ (59). Associations between lowered inflammation and elevated SCZ PRS have been suggested by two Mendelian Randomization studies showing that people genetically predisposed to lower CRP levels have an increased risk of SCZ, potentially due to a weakened ability to fight early-life infection (91, 92). Viral infections in particular have been hypothesized to play a causative role in SCZ pathogenesis (93), which may explain the association between SCZ PRS and increased lymphocyte counts, a trait replicated in some SCZ patients (94). T-lymphocytes are involved in viral responses (95), and their viral-induced activation may drive SCZ pathophysiology in a similar pro-inflammatory manner to that suggested for MDD development. Future studies investigating blood-based markers of psychiatric conditions should therefore seek to incorporate additional inflammatory marker measurements such as pro-inflammatory cytokines, to possibly elicit more information surrounding the MDD and SCZ disorder-associated inflammatory response.

The various associations between SCZ PRS and renal markers are consistent with studies showing higher chronic kidney disease incidence in SCZ patients (96, 97). Although these are regarded as canonical renal function markers, some, such as cystatin C, have also been shown to exert neurobiological influences. In several *in vivo* models subjected to brain injury, cystatin C was found to be neuroprotective and helped restore BBB integrity (98, 99). These findings possibly pertain to the negative association found between cystatin C and increased SCZ PRS, and several associations with BBB-permeable markers. A potential role for cystatin C in SCZ pathogenesis has yet to be thoroughly investigated, and this observation demonstrates the power of big data studies like ours to identify novel markers possibly involved in disorder etiology.

Although genetically driven perturbations to markers such as cystatin C may contribute to BBB dysfunction, our findings suggest that BBB impairment cannot solely be attributed to increased genetic risk. A link between genetic liability and BBB dysregulation in MDD and SCZ was initially implied by the associations detected for most BBB-permeable markers in our MA model. The subsequent loss of associations upon controlling for lifestyle factors suggests BBB function may be mediated by both genetic and environmental cofounders. Obesity, smoking and alcohol status have all been previously implicated in BBB dysregulation (100, 101, 102). However, the individual genetic and environmental contributions to psychiatric disorder-associated BBB impairment remain unclear.

Interestingly, fewer associations were detected with inflammatory, immune and BBB-permeable markers in subjects with higher BD PRS. Although inflammatory and immune abnormalities have been observed in BD patients (103), these may well be effects of the condition, rather than causative. However, it is also possible that fewer associations were detected due to the lower sample sizes employed in the Stahl *et al* (46) GWAS used to determine BD PRS, reducing their predictive power.

Nonetheless, we still observed unique associations for BD, including with alkaline phosphatase and calcium, which along with vitamin D are bone health markers, and may relate to bone abnormalities seen in some BD patients (104). The higher proportion of markers that remained significant after correcting for lifestyle factors may also suggest genetic alterations have a greater influence on the peripheral profile for BD in comparison to MDD and SCZ.

## Limitations

Our study has several limitations, perhaps most notably that in UKB we are examining associations in a predominantly healthy population with an older age range. This could also explain why some of our results do not reflect those of patient studies, such as one showing that increased CRP levels predispose to SCZ onset in adolescent participants (105), a sample less likely to be cofounded by long-standing illness and disorder related medications. Moreover, our analyses are based on single cross-sectional assessments, as repeated measurements of peripheral marker levels are planned for only a subset of the UKB cohort (43). Consequently, we cannot determine whether the changes in peripheral markers we observed are stable over time.

In addition, peripheral marker levels may be significantly influenced by environmental and lifestyle factors unaccounted for by our models, which may play a role in the pathogenesis of MDD, SCZ and BD in conjunction with genetic risk variants (106). This is especially relevant to MDD, which has been reported to have a lower heritability than both schizophrenia and bipolar disorders (107), and demonstrates the importance of other approaches such as Mendelian randomization to infer genetic causality. Additionally, lifestyle factors controlled for in our FA model such as obesity are influenced by both genetics and environment, the individual contributions of which we cannot directly determine (108).

Moreover, our study does not provide direct confirmation of BBB attenuation in participants, and we are only examining peripheral BBB-permeable marker associations, reaffirming the value of *in vivo* models that permit simultaneous blood and brain measurements of these markers. In addition, BBB-permeable markers such as glucose are involved in a myriad of physiological processes, some of which may influence associations more than BBB dysfunction. Direct evidence for BBB impairment could be obtained in future studies by taking cerebrospinal fluid measurements from participants and incorporating non-invasive BBB-imaging techniques, as well as advancement in BBB laboratory models.

We also cannot exclude the possibility of some participant overlap between UKB and SCZ and BD GWAS cohorts used to determine PRS and associations, as summary statistics from the PGC SCZ and BD GWAS do not provide this information. However the effects of this limitation may be considered minor and confined to controls, due to our efforts to minimize overlap through removal of cases in our UKB sample.

Finally, our results may have limited generalizability as we only included Caucasian participants. Future studies should therefore seek to independently replicate these findings in different datasets and such populations with larger sample sizes than ours. Greater participation in BD GWAS may also yield more associations for BD PRS.

## Conclusions

In summary, we have demonstrated varying associations between SCZ, MDD and BD PRS with BBB-permeable, inflammatory and other blood-based markers, influenced by genetic risk and robust to controlling for lifestyle factors. This reaffirms that despite their shared genetic risk, aspects of their underlying neurobiology can be differentiated. Our study also reinforces the potential value and importance of examining peripheral components of disorder pathogenesis. Incorporation of additional markers, laboratory models, longitudinal data and analyses of other datasets will facilitate testing for causal inferences.

## Data Availability

Details concerning access to UK Biobank data can be found in the link provided.

https://www.ukbiobank.ac.uk/principles-of-access/

## Acknowledgements and Disclosures

This study was supported by a Wellcome Trust Strategic Award (“Stratifying Resilience and Depression Longitudinally”; Grant No. 104036/Z/14/Z; principal investigator, AMM). This research was conducted using the UKB Resource under approved project 4844. MDES, LJS (both of whom supervised by XS, HCW and SL throughout the duration of the project) and AJES all receive support from the University of Edinburgh Wellcome Trust Translational Neuroscience 4-year PhD programme (Grant No. 108890/Z/15/Z). We would like to thank UKB participants for their time and UKB team members for collating the data.

