## Supplement for "Associations Between Major Psychiatric Disorder Polygenic Risk Scores and Blood-Based Markers in UK Biobank"

**Supplementary Information**

**Table of Contents**

Supplementary Methods

-Participants

-Covariates

-PRS Generation and Table S1

-Blood Markers and Tables S2-S3

Supplementary Results

- PRS Associations with blood markers at threshold ≤ 0.5 for each disorder, Tables S4-S9

- Sensitivity analysis of major depressive disorder polygenic risk score associations, Table S10

- Disorder PRS Associations with blood markers at all other PRS thresholds, Tables S11-S34

Supplementary References

**Supplementary Methods**

***Participants***

The flow chart below (**Figure S1**) demonstrates how we derived our final sample sizes employed to examine blood marker associations with each disorder PRS.

**Figure S1.** Outline of scheme of work employed to generate final participant sample from UK Biobank for disorder polygenic risk scores (PRS) association analysis with peripheral markers. ICD, International Classification Disease codes; PGC, Psychiatric Genomics Consortium; GWAS, genome-wide association studies.

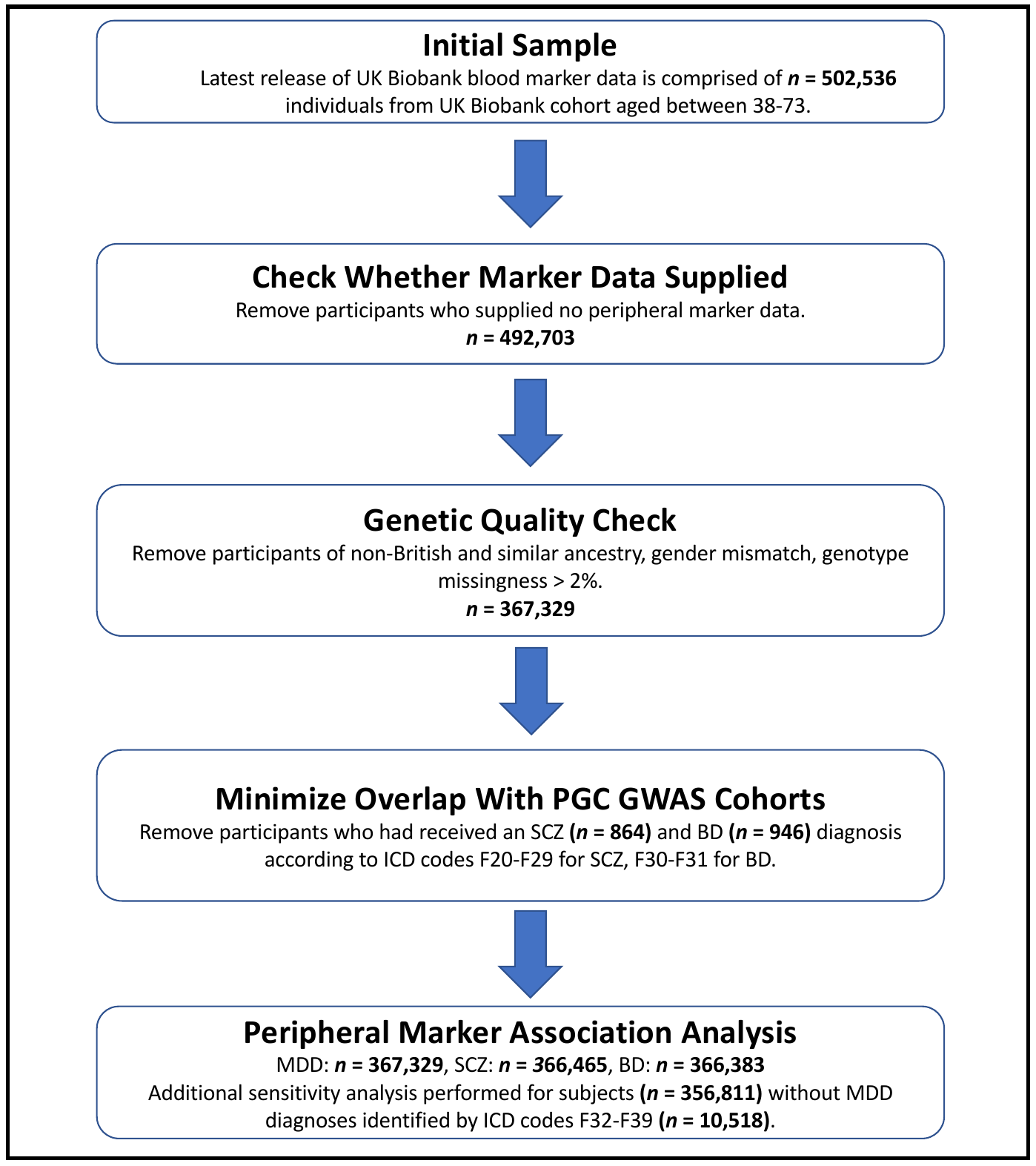

***Covariates***

Genetic principal components computed by UKB using the fastPCA algorithm to account for population stratification (1). Body mass index (BMI) was calculated by dividing weight (kg) (measured to the nearest 0.1kg using a BC-418 MA body composition analyzer; Tanita Corp) by height (measured in cm with a Seca 202 stadiometer) in meters squared. The mean BMI for the sample of *n* = 366,099 participants who had passed genetic quality check and for whom BMI data were available, was 27.36 ± 4.76 kg/m^2^ Alcohol and smoking status data from participant questionnaires, in which respondents were asked to describe both with the following options: ‘never’ (*n* = 11,535 for drinking, *n* = 197,753 for smoking), ‘previous’ (*n* = 12,699 for drinking, *n* = 130,033 for smoking), ‘current’ (*n* = 342,536 for drinking, *n* = 38,051 for smoking) and ‘prefer not to answer’ (*n* = 360 for drinking, *n* = 1,293 for smoking), were collated by UKB at baseline (numbers in brackets are derived from our sample of *n* = 367,130 participants who had passed genetic quality checks and for whom data were available).

***PRS Generation***

**Table S1.** Number of SNPs at all PRS thresholds we examined associations with peripheral markers for, prior to and after linkage disequilibrium clump-based pruning.

| **Disorder** | **Number of Unpruned SNPs** | | | | | **Number of Pruned SNPs** | | | | |
| --- | --- | --- | --- | --- | --- | --- | --- | --- | --- | --- |
|  | **p ≤  .01** | **p ≤  .05** | **p ≤  .1** | **p ≤  .5** | **p ≤  1.0** | **p ≤  .01** | **p ≤  .05** | **p ≤  .1** | **p ≤  .5** | **p ≤  1.0** |
| Bipolar Disorder | 256,789 | 918,318 | 1,661,317 | 7,067,611 | 13,413,245 | 5,573 | 20,936 | 37,258 | 145,975 | 247,595 |
| Major Depressive Disorder | 310,670 | 967,708 | 1,632,632 | 5,972,644 | 10,863,279 | 6,485 | 22,321 | 38,584 | 141,802 | 237,080 |
| Schizophrenia | 428,876 | 1,085,119 | 1,702,616 | 5,423,695 | 9,444,231 | 8,271 | 24,069 | 41,081 | 138,305 | 225,161 |

***Blood Markers***

***Peripheral Marker Classification***

Peripheral markers were classified as blood-brain barrier (BBB)-permeable after conducting an extensive literature search with PubMed (<https://pubmed.ncbi.nlm.nih.gov/>). Our search revealed the following markers are BBB-permeable:, estradiol (2), glucose (3), high-density lipoprotein (HDL) cholesterol (4), insulin-like growth factor 1 (IGF-1) (5), sex hormone binding globulin (SHBG) (6), testosterone (7), triglycerides (8) and vitamin D (9).

“Inflammatory and Hematological” markers are a combination of parameters previously classified as “hematological” by UK Biobank ([https://biobank.ndph.ox.ac.uk/showcase/showcase/docs/hematology.pdf](https://biobank.ndph.ox.ac.uk/showcase/showcase/docs/haematology.pdf)), and markers identified through a PubMed literature search as primary inflammatory indicators: C-reactive protein (10), neutrophil-to-lymphocyte ratio (11). We grouped inflammatory and hematological markers together, as inflammatory responses are known to induce changes in hematological parameters that are also components of the immune system (12, 13).

Liver, renal and cardiovascular marker sorting was achieved using previous classification by UK Biobank (<https://www.ukbiobank.ac.uk/wp-content/uploads/2018/11/BCM023_ukb_biomarker_panel_website_v1.0-Aug-2015-edit-2018.pdf>). However, we classified alkaline phosphatase as an additional parameter of liver function (14). All other markers that were neither found to be BBB-permeable or inflammatory from our literature search, nor classified by UK Biobank as hematological, liver, renal or cardiovascular function parameters, were collated into the “Other” category.

***Estradiol and Rheumatoid Factor Data Processing***

Estradiol and rheumatoid factor (RF) levels were initially recorded as “missing” for a large number of participants due to detected levels deemed lower than the reportable range <https://biobank.ndph.ox.ac.uk/showcase/showcase/docs/biomarker_issues.pdf>. For these participants, who also had data for at least one other biomarker, we recoded estradiol and RF levels as the square root of the minimum stated detectable value, an approach which has previously been employed (15).

***Blood Marker Datasets***

**Table S2.** Baseline data from the latest UK Biobank data release for peripheral markers for which associations were determined with MDD, BD and SCZ PGRS, for *n* = 502,536 participants aged 37-73 years. BBB, blood-brain barrier; CRP, C-reactive protein; HDL, high-density lipoprotein; IGF-1, insulin-like growth factor 1; Min., minimum value; Max., maximum value; LDL, low-density lipoprotein; S.D., standard deviation; SHBG, sex hormone binding globulin.

| **Biomarker** | ***N* =** | **Mean** | **S.D.** | **Min.** | **Max.** |
| --- | --- | --- | --- | --- | --- |
| ***BBB-Permeable*** |  |  |  |  |  |
| Estradiol | 76674 | 461.17 | 431.16 | 175.00 | 14588.00 |
| Glucose | 429590 | 5.13 | 1.24 | 1.00 | 36.81 |
| HDL cholesterol | 429894 | 1.45 | 0.38 | 0.22 | 4.40 |
| IGF-1 | 467066 | 21.40 | 5.70 | 1.45 | 126.77 |
| SHBG | 425869 | 51.63 | 27.78 | 0.39 | 241.92 |
| Testosterone | 425232 | 6.56 | 6.05 | 0.35 | 54.34 |
| Triglycerides | 469240 | 1.75 | 1.03 | 0.23 | 11.28 |
| Vitamin D | 448376 | 48.61 | 21.11 | 10.00 | 340.00 |
| ***Inflammatory and Hematological*** | | | | | |
| Basophil count | 477311 | 0.03 | 0.05 | 0.00 | 3.03 |
| Basophil percentage | 477317 | 0.57 | 0.61 | 0.00 | 33.80 |
| CRP | 468594 | 2.60 | 4.36 | 0.08 | 79.96 |
| Eosinophil count | 477311 | 0.18 | 0.14 | 0.00 | 9.60 |
| Eosinophil percentage | 477317 | 2.57 | 1.88 | 0.00 | 100.00 |
| Hematocrit percentage | 478200 | 41.09 | 3.56 | 0.05 | 72.48 |
| Hemoglobin concentration | 478200 | 14.18 | 1.25 | 0.09 | 22.27 |
| High light scatter reticulocyte count | 469878 | 0.02 | 0.01 | 0.00 | 0.60 |
| High light scatter reticulocyte percentage | 469879 | 0.40 | 0.33 | 0.00 | 80.00 |
| Immature reticulocyte fraction | 469877 | 0.29 | 0.06 | 0.00 | 1.00 |
| Lymphocyte count | 477311 | 1.97 | 1.17 | 0.00 | 196.41 |
| Lymphocyte percentage | 477317 | 28.91 | 7.50 | 0.00 | 98.70 |
| Mean corpuscular hemoglobin | 478197 | 31.45 | 1.92 | 0.00 | 95.67 |
| Mean corpuscular hemoglobin concentration | 478193 | 34.51 | 1.08 | 16.10 | 97.30 |
| Mean corpuscular volume | 478198 | 91.12 | 4.61 | 52.10 | 160.30 |
| Mean platelet (thrombocyte) volume | 478192 | 9.33 | 1.09 | 5.73 | 16.50 |
| Mean reticulocyte volume | 469877 | 105.92 | 7.83 | 46.00 | 249.45 |
| Mean sphered cell volume | 469879 | 82.87 | 5.32 | 43.31 | 205.20 |
| Monocyte count | 477311 | 0.48 | 0.27 | 0.00 | 113.39 |
| Monocyte percentage | 477317 | 7.06 | 2.70 | 0.00 | 96.90 |
| Neutrophil count | 477311 | 4.23 | 1.42 | 0.00 | 76.42 |
| Neutrophil percentage | 477317 | 60.88 | 8.53 | 0.00 | 97.70 |
| Nucleated red blood cell count | 477300 | 0.00 | 0.03 | 0.00 | 6.90 |
| Nucleated red blood cell percentage | 477296 | 0.04 | 0.45 | 0.00 | 81.97 |
| Platelet count | 478197 | 252.99 | 60.05 | 0.30 | 1821.00 |
| Platelet crit | 478193 | 0.23 | 0.05 | 0.00 | 1.45 |
| Platelet distribution width | 478192 | 16.49 | 0.52 | 13.27 | 20.20 |
| Red blood cell (erythrocyte) count | 478200 | 4.52 | 0.42 | 0.01 | 7.91 |
| Red blood cell (erythrocyte) distribution width | 478198 | 13.49 | 0.99 | 2.28 | 38.96 |
| Reticulocyte count | 469878 | 0.06 | 0.04 | 0.00 | 2.43 |
| Reticulocyte percentage | 469878 | 1.35 | 0.89 | 0.00 | 90.91 |
| White blood cell (leukocyte) count | 478195 | 6.89 | 2.12 | 0.00 | 389.70 |
| ***Liver*** | | | | | |
| Albumin | 430098 | 45.21 | 2.63 | 17.38 | 59.80 |
| Alkaline phosphatase | 469628 | 83.67 | 26.46 | 8.00 | 1416.70 |
| Alanine aminotransferase | 469427 | 23.55 | 14.18 | 3.01 | 495.19 |
| Aspartate aminotransferase | 467822 | 26.23 | 10.66 | 3.30 | 947.20 |
| Direct bilirubin | 398634 | 1.83 | 0.85 | 1.00 | 70.06 |
| Gamma glutamyltransferase | 469368 | 37.39 | 42.09 | 5.00 | 1184.90 |
| Total bilirubin | 467586 | 9.13 | 4.43 | 1.08 | 144.52 |
| ***Renal*** | | | | | |
| Creatinine | 469383 | 72.31 | 18.55 | 10.70 | 1499.30 |
| Cystatin C | 469584 | 0.91 | 0.18 | 0.30 | 7.49 |
| Phosphate | 429274 | 1.16 | 0.16 | 0.37 | 4.70 |
| Total protein | 429627 | 72.51 | 4.12 | 36.27 | 117.36 |
| Urate | 469051 | 309.21 | 80.43 | 89.10 | 1067.50 |
| Urea | 469297 | 5.40 | 1.40 | 0.81 | 41.83 |
| ***Cardiovascular*** | | | | | |
| Apolipoprotein A | 427532 | 1.54 | 0.27 | 0.42 | 2.50 |
| Apolipoprotein B | 467232 | 1.03 | 0.24 | 0.40 | 2.00 |
| Cholesterol | 469615 | 5.69 | 1.15 | 0.60 | 15.46 |
| LDL direct | 468732 | 3.56 | 0.87 | 0.27 | 9.80 |
| Lipoprotein A | 375654 | 44.65 | 49.21 | 3.80 | 189.00 |
| ***Other Non-BBB-Permeable-Markers*** | | | | | |
| Calcium | 429955 | 2.38 | 0.09 | 1.05 | 3.61 |
| Glycated hemoglobin | 466530 | 36.13 | 6.78 | 15.00 | 515.20 |
| Rheumatoid factor | 41316 | 24.56 | 19.86 | 10.00 | 120.00 |

**Table S3.** Peripheral marker data with removed outliers, recoded estradiol and rheumatoid factor levels and calculated neutrophil-to-lymphocyte (NLR) ratios for *n* = 367,329 participants who had passed genetic quality checks. BBB, blood-brain barrier; CRP, C-reactive protein; HDL, high-density lipoprotein; IGF-1, insulin-like growth factor 1; LDL, low-density lipoprotein; Min., minimum value; Max., maximum value; NLR, neutrophil-to-lymphocyte ratio; S.D., standard deviation; SHBG, sex hormone binding globulin.

| **Biomarker** | ***N* =** | **Mean** | **S.D.** | **Min.** | **Max.** |
| --- | --- | --- | --- | --- | --- |
| ***BBB-Permeable*** | | | | | |
| Estradiol | 323013 | 73.76 | 177.33 | 8.54 | 1326.80 |
| Glucose | 318519 | 5.04 | 0.82 | 1.01 | 11.16 |
| HDL cholesterol | 321142 | 1.45 | 0.38 | 0.22 | 3.37 |
| IGF-1 | 348798 | 21.41 | 5.57 | 1.91 | 49.81 |
| SHBG | 317455 | 51.59 | 26.72 | 0.39 | 191.06 |
| Testosterone | 317923 | 6.60 | 6.05 | 0.35 | 36.81 |
| Triglycerides | 349486 | 1.73 | 0.96 | 0.23 | 6.86 |
| Vitamin D | 335347 | 49.47 | 20.78 | 10.00 | 154.00 |
| ***Inflammatory and Hematological*** | | | | | |
| Basophil count | 354979 | 0.03 | 0.04 | 0.00 | 0.28 |
| Basophil percentage | 355053 | 0.54 | 0.40 | 0.00 | 3.54 |
| CRP | 347591 | 2.31 | 2.99 | 0.08 | 24.26 |
| Eosinophil count | 355204 | 0.17 | 0.12 | 0.00 | 0.85 |
| Eosinophil percentage | 355307 | 2.51 | 1.65 | 0.00 | 11.77 |
| Hematocrit percentage | 356899 | 41.15 | 3.50 | 23.63 | 58.43 |
| Hemoglobin concentration | 356931 | 14.21 | 1.23 | 8.06 | 20.30 |
| High light scatter reticulocyte count | 350687 | 0.02 | 0.01 | 0.00 | 0.07 |
| High light scatter reticulocyte percentage | 350816 | 0.40 | 0.20 | 0.00 | 1.91 |
| Immature reticulocyte fraction | 350988 | 0.29 | 0.06 | 0.00 | 0.59 |
| Lymphocyte count | 356039 | 1.93 | 0.61 | 0.00 | 7.40 |
| Lymphocyte percentage | 356048 | 28.62 | 7.20 | 0.00 | 65.30 |
| Mean corpuscular hemoglobin | 356084 | 31.54 | 1.66 | 22.37 | 40.73 |
| Mean corpuscular hemoglobin concentration | 356571 | 34.53 | 0.92 | 29.19 | 39.88 |
| Mean corpuscular volume | 356351 | 91.35 | 4.25 | 69.33 | 113.30 |
| Mean platelet thrombocyte volume | 356955 | 9.32 | 1.08 | 5.73 | 14.71 |
| Mean reticulocyte volume | 350516 | 105.85 | 7.58 | 66.99 | 144.71 |
| Mean sphered cell volume | 350747 | 82.88 | 5.18 | 56.68 | 109.15 |
| Monocyte count | 355820 | 0.47 | 0.17 | 0.00 | 1.57 |
| Monocyte percentage | 355759 | 7.03 | 2.16 | 0.00 | 20.58 |
| Neutrophil count | 355911 | 4.24 | 1.37 | 0.00 | 11.30 |
| Neutrophil percentage | 355874 | 61.20 | 8.16 | 19.28 | 97.70 |
| NLR | 355294 | 2.35 | 0.98 | 0.00 | 8.66 |
| Nucleated red blood cell count | 354842 | 0.00 | 0.01 | 0.00 | 0.16 |
| Nucleated red blood cell percentage | 354741 | 0.01 | 0.13 | 0.00 | 2.04 |
| Platelet count | 356646 | 252.58 | 58.15 | 0.30 | 552.00 |
| Platelet crit | 356663 | 0.23 | 0.05 | 0.00 | 0.48 |
| Platelet distribution width | 356822 | 16.49 | 0.52 | 14.13 | 19.08 |
| Red blood cell erythrocyte count | 356880 | 4.51 | 0.41 | 2.47 | 6.55 |
| Red blood cell erythrocyte distribution width | 355391 | 13.44 | 0.82 | 10.47 | 18.23 |
| Reticulocyte count | 350500 | 0.06 | 0.02 | 0.00 | 0.25 |
| Reticulocyte percentage | 350459 | 1.33 | 0.51 | 0.00 | 5.69 |
| White blood cell leukocyte count | 356682 | 6.87 | 1.74 | 0.00 | 16.98 |
| ***Liver*** | | | | | |
| Albumin | 321314 | 45.25 | 2.61 | 32.23 | 58.28 |
| Alkaline phosphatase | 350200 | 82.89 | 23.02 | 8.00 | 215.90 |
| Alanine aminotransferase | 349242 | 23.06 | 11.63 | 3.01 | 94.61 |
| Aspartate aminotransferase | 348166 | 25.81 | 7.58 | 3.30 | 79.20 |
| Direct bilirubin | 297755 | 1.82 | 0.74 | 1.00 | 6.10 |
| Gamma glutamyltransferase | 348627 | 35.12 | 28.70 | 5.00 | 248.50 |
| Total bilirubin | 347935 | 9.04 | 3.96 | 1.08 | 31.35 |
| ***Renal*** | | | | | |
| Creatinine | 350192 | 71.98 | 14.56 | 10.80 | 162.60 |
| Cystatin C | 349950 | 0.90 | 0.15 | 0.30 | 1.77 |
| Phosphate | 320718 | 1.16 | 0.16 | 0.37 | 1.95 |
| Total protein | 320937 | 72.36 | 4.02 | 52.44 | 92.52 |
| Urate | 350405 | 309.05 | 80.20 | 89.20 | 710.00 |
| Urea | 350019 | 5.41 | 1.30 | 0.91 | 12.38 |
| ***Cardiovascular*** | | | | | |
| Apolipoprotein A | 319424 | 1.54 | 0.27 | 0.42 | 2.50 |
| Apolipoprotein B | 349143 | 1.03 | 0.24 | 0.40 | 2.00 |
| Cholesterol | 350856 | 5.71 | 1.14 | 0.60 | 11.42 |
| LDL direct | 350212 | 3.57 | 0.87 | 0.27 | 7.90 |
| Lipoprotein A | 279630 | 43.95 | 49.22 | 3.80 | 189.00 |
| ***Other Non-BBB-Permeable*** | | | | | |
| Calcium | 321026 | 2.38 | 0.09 | 1.92 | 2.85 |
| Glycated hemoglobin | 348370 | 35.62 | 5.12 | 15.30 | 68.50 |
| Rheumatoid factor | 345794 | 4.41 | 4.91 | 3.16 | 47.30 |

**Supplementary Results**

***PRS Associations with blood markers at threshold ≤ 0.5 for each disorder***

**Table S4.** Peripheral marker minimally adjusted associations with polygenic risk scores (PRS) at threshold ≤ 0.5 for major depressive disorder. BBB, blood-brain barrier; CRP, C-reactive protein; HDL, high-density lipoprotein; IGF-1, insulin-like growth factor 1; LDL, low-density lipoprotein; NLR, neutrophil-to-lymphocyte ratio; p-uncorr., p-uncorrected value; p-corr., Bonferroni p-corrected value; S.E., standard error; SHBG, sex hormone binding globulin. ***p corr. ≤ 0.001, **p corr. ≤ 0.01, *p corr. ≤ 0.05

| **Biomarker** | **Effect Size (β)** | **S.E.** | **t statistic** | **p-uncorr.** | **p-corr.** |
| --- | --- | --- | --- | --- | --- |
| ***BBB-Permeable*** | | | | | |
| Estradiol | -7.82e-4 | 1.59e-3 | -0.493 | 0.622 | 1.000 |
| Glucose | 0.009 | 1.76e-3 | 5.016 | 5.27e-7 | 3.27e-5*** |
| HDL cholesterol | -0.017 | 1.61e-3 | -10.332 | 5.11e-25 | 3.17e-23*** |
| IGF-1 | -0.016 | 1.63e-3 | -10.035 | 1.08e-23 | 6.70e-22*** |
| SHBG | -0.009 | 1.62e-3 | -5.716 | 1.09e-8 | 6.78e-7*** |
| Testosterone | -3.00e-3 | 7.79e-4 | -3.854 | 1.16e-4 | 0.007** |
| Triglycerides | 0.023 | 1.66e-3 | 14.185 | 1.17e-45 | 7.28e-44*** |
| Vitamin D | -0.019 | 1.72e-3 | -10.852 | 1.97e-27 | 1.22e-25*** |
| ***Inflammatory and Hematological*** | | | | | |
| Basophil count | 0.008 | 1.66e-3 | 4.647 | 3.36e-6 | 2.09e-4*** |
| Basophil percentage | 2.48e-4 | 1.68e-3 | 0.147 | 0.883 | 1.000 |
| CRP | 0.022 | 1.70e-3 | 13.006 | 1.16e-38 | 7.19e-37*** |
| Eosinophil count | 0.007 | 1.68e-3 | 4.168 | 3.08e-5 | 1.91e-3** |
| Eosinophil percentage | -2.85e-3 | 1.68e-3 | -1.702 | 0.089 | 1.000 |
| Hematocrit percentage | 9.21e-4 | 1.37e-3 | 0.672 | 0.502 | 1.000 |
| Hemoglobin concentration | 1.40e-3 | 1.34e-3 | 1.049 | 0.294 | 1.000 |
| High light scatter reticulocyte count | 0.021 | 1.68e-3 | 12.552 | 3.93e-36 | 2.43e-34*** |
| High light scatter reticulocyte percentage | 0.021 | 1.69e-3 | 12.573 | 3.04e-36 | 1.88e-34*** |
| Immature reticulocyte fraction | 0.018 | 1.68e-3 | 10.858 | 1.85e-27 | 1.15e-25*** |
| Lymphocyte count | 0.019 | 1.67e-3 | 11.204 | 3.95e-29 | 2.45e-27*** |
| Lymphocyte percentage | -0.006 | 1.66e-3 | -3.461 | 5.38e-4 | 0.033* |
| Mean corpuscular hemoglobin | 1.69e-3 | 1.66e-3 | 1.017 | 0.309 | 1.000 |
| Mean corpuscular hemoglobin concentration | 1.25e-3 | 1.66e-3 | 0.749 | 0.454 | 1.000 |
| Mean corpuscular volume | 1.52e-3 | 1.67e-3 | 0.911 | 0.362 | 1.000 |
| Mean platelet thrombocyte volume | 3.34e-3 | 1.68e-3 | 1.987 | 0.047 | 1.000 |
| Mean reticulocyte volume | 4.16e-3 | 1.68e-3 | 2.473 | 0.013 | 0.830 |
| Mean sphered cell volume | 1.33e-3 | 1.69e-3 | 0.789 | 0.430 | 1.000 |
| Monocyte count | 0.013 | 1.63e-3 | 7.976 | 1.52e-15 | 9.40e-14*** |
| Monocyte percentage | -0.007 | 1.63e-3 | -4.582 | 4.62e-6 | 2.86e-4*** |
| Neutrophil count | 0.025 | 1.68e-3 | 14.760 | 2.75e-49 | 1.70e-47*** |
| Neutrophil percentage | 0.007 | 1.68e-3 | 4.309 | 1.64e-5 | 1.02e-3** |
| NLR | 0.006 | 1.67e-3 | 3.724 | 1.96e-4 | 0.012* |
| Nucleated red blood cell count | 1.02e-3 | 1.69e-3 | 0.606 | 0.544 | 1.000 |
| Nucleated red blood cell percentage | 1.51e-3 | 1.69e-3 | 0.897 | 0.370 | 1.000 |
| Platelet count | 0.006 | 1.62e-3 | 3.569 | 3.58e-4 | 0.022* |
| Platelet crit | 0.008 | 1.59e-3 | 5.174 | 2.29e-7 | 1.42e-5*** |
| Platelet distribution width | 4.70e-3 | 1.66e-3 | 2.824 | 4.75e-3 | 0.294 |
| Red blood cell erythrocyte count | 9.19e-5 | 1.44e-3 | 0.064 | 0.949 | 1.000 |
| Red blood cell erythrocyte distribution width | 0.006 | 1.68e-3 | 3.872 | 1.08e-4 | 0.007** |
| Reticulocyte count | 0.018 | 1.67e-3 | 10.767 | 4.96e-27 | 3.07e-25*** |
| Reticulocyte percentage | 0.019 | 1.69e-3 | 11.136 | 8.46e-29 | 5.25e-27*** |
| White blood cell leukocyte count | 0.028 | 1.68e-3 | 16.751 | 5.92e-63 | 3.67e-61*** |
| ***Liver*** | | | | | |
| Albumin | -0.007 | 1.74e-3 | -3.906 | 9.39e-5 | 0.006** |
| Alkaline phosphatase | 0.011 | 1.66e-3 | 6.569 | 5.07e-11 | 3.14e-9*** |
| Alanine aminotransferase | 0.011 | 1.62e-3 | 6.904 | 5.08e-12 | 3.15e-10*** |
| Aspartate aminotransferase | 0.008 | 1.65e-3 | 4.874 | 1.09e-6 | 6.79e-5*** |
| Direct bilirubin | -0.006 | 1.79e-3 | -3.544 | 3.94e-4 | 0.024* |
| Gamma glutamyltransferase | 0.016 | 1.65e-3 | 9.926 | 3.23e-23 | 2.00e-21*** |
| Total bilirubin | -0.014 | 1.65e-3 | -8.324 | 8.52e-17 | 5.28e-15*** |
| ***Renal*** | | | | | |
| Creatinine | -0.008 | 1.37e-3 | -5.557 | 2.75e-8 | 1.71e-6*** |
| Cystatin C | 0.016 | 1.55e-3 | 10.231 | 1.46e-24 | 9.03e-23*** |
| Phosphate | -4.15e-4 | 1.71e-3 | -0.243 | 0.808 | 1.000 |
| Total protein | -8.79e-4 | 1.77e-3 | -0.498 | 0.619 | 1.000 |
| Urate | 0.008 | 1.44e-3 | 5.480 | 4.25e-8 | 2.64e-6*** |
| Urea | -0.005 | 1.62e-3 | -3.241 | 1.19e-3 | 0.074 |
| ***Cardiovascular*** | | | | | |
| Apolipoprotein A | -0.013 | 1.63e-3 | -7.945 | 1.95e-15 | 1.21e-13*** |
| Apolipoprotein B | 1.36e-4 | 1.69e-3 | 0.081 | 0.936 | 1.000 |
| Cholesterol | -0.006 | 1.66e-3 | -3.507 | 4.54e-4 | 0.028* |
| LDL direct | -4.48e-3 | 1.68e-3 | -2.669 | 0.008 | 0.472 |
| Lipoprotein A | -6.77e-5 | 1.90e-3 | -0.036 | 0.972 | 1.000 |
| ***Other Non-BBB-Permeable*** | | | | | |
| Calcium | 3.81e-3 | 1.76e-3 | 2.164 | 0.030 | 1.000 |
| Glycated hemoglobin | 0.015 | 1.64e-3 | 9.411 | 4.94e-21 | 3.06e-19*** |
| Rheumatoid factor | -3.50e-3 | 1.71e-3 | -2.049 | 0.040 | 1.000 |

**Table S5.** Peripheral marker minimally adjusted associations with polygenic risk scores (PRS) at threshold ≤ 0.5 for schizophrenia. BBB, blood-brain barrier; CRP, C-reactive protein; HDL, high-density lipoprotein; IGF-1, insulin-like growth factor 1; LDL, low-density lipoprotein; NLR, neutrophil-to-lymphocyte ratio; p-uncorr., p-uncorrected value; p-corr., Bonferroni p-corrected value; S.E., standard error; SHBG, sex hormone binding globulin. ***p corr. ≤ 0.001, **p corr. ≤ 0.01, *p corr. ≤ 0.05

| **Biomarker** | **Effect Size (β)** | **S.E.** | **t statistic** | **p-uncorr.** | **p-corr.** |
| --- | --- | --- | --- | --- | --- |
| ***BBB-Permeable*** | | | | | |
| Estradiol | 4.32e-3 | 1.65e-3 | 2.614 | 0.009 | 0.555 |
| Glucose | -0.008 | 1.83e-3 | -4.171 | 3.03e-5 | 1.88e-3** |
| HDL cholesterol | 0.011 | 1.68e-3 | 6.817 | 9.34e-12 | 5.79e-10*** |
| IGF-1 | 4.53e-3 | 1.71e-3 | 2.657 | 0.008 | 0.488 |
| SHBG | 0.014 | 1.69e-3 | 8.194 | 2.53e-16 | 1.57e-14*** |
| Testosterone | 4.74e-3 | 8.13e-4 | 5.823 | 5.77e-9 | 3.58e-7*** |
| Triglycerides | -0.006 | 1.73e-3 | -3.510 | 4.47e-4 | 0.028* |
| Vitamin D | -0.014 | 1.80e-3 | -7.660 | 1.86e-14 | 1.16e-12*** |
| ***Inflammatory and Hematological*** | | | | | |
| Basophil count | 0.005 | 1.73e-3 | 3.114 | 1.85e-3 | 0.114 |
| Basophil percentage | 0.006 | 1.75e-3 | 3.259 | 1.12e-3 | 0.069 |
| CRP | -0.007 | 1.77e-3 | -3.957 | 7.59e-5 | 4.71e-3** |
| Eosinophil count | 0.008 | 1.75e-3 | 4.787 | 1.69e-6 | 1.05e-4*** |
| Eosinophil percentage | 0.005 | 1.75e-3 | 2.993 | 2.76e-3 | 0.171 |
| Hematocrit percentage | 5.38e-4 | 1.43e-3 | 0.376 | 0.707 | 1.000 |
| Hemoglobin concentration | 1.35e-3 | 1.39e-3 | 0.967 | 0.334 | 1.000 |
| High light scatter reticulocyte count | -0.010 | 1.75e-3 | -5.867 | 4.44e-9 | 2.75e-7*** |
| High light scatter reticulocyte percentage | -0.010 | 1.76e-3 | -5.555 | 2.78e-8 | 1.72e-6*** |
| Immature reticulocyte fraction | -0.008 | 1.75e-3 | -4.317 | 1.59e-5 | 9.83e-4*** |
| Lymphocyte count | 0.015 | 1.74e-3 | 8.614 | 7.05e-18 | 4.37e-16*** |
| Lymphocyte percentage | 0.009 | 1.73e-3 | 5.061 | 4.16e-7 | 2.58e-5*** |
| Mean corpuscular hemoglobin | 0.009 | 1.73e-3 | 4.988 | 6.11e-7 | 3.79e-5*** |
| Mean corpuscular hemoglobin concentration | 1.78e-3 | 1.73e-3 | 1.026 | 0.305 | 1.000 |
| Mean corpuscular volume | 0.009 | 1.74e-3 | 4.989 | 6.08e-7 | 3.77e-5*** |
| Mean platelet thrombocyte volume | -7.90e-4 | 1.75e-3 | -0.451 | 0.652 | 1.000 |
| Mean reticulocyte volume | 0.006 | 1.75e-3 | 3.515 | 4.40e-4 | 0.027* |
| Mean sphered cell volume | 0.011 | 1.76e-3 | 6.360 | 2.02e-10 | 1.26e-8*** |
| Monocyte count | 4.44e-3 | 1.70e-3 | 2.614 | 0.009 | 0.555 |
| Monocyte percentage | -3.05e-3 | 1.70e-3 | -1.796 | 0.073 | 1.000 |
| Neutrophil count | 4.36e-3 | 1.75e-3 | 2.492 | 0.013 | 0.788 |
| Neutrophil percentage | -0.008 | 1.75e-3 | -4.777 | 1.78e-6 | 1.10e-4*** |
| NLR | -0.007 | 1.74e-3 | -4.258 | 2.06e-5 | 1.28e-3** |
| Nucleated red blood cell count | -2.43e-3 | 1.76e-3 | -1.381 | 0.167 | 1.000 |
| Nucleated red blood cell percentage | -1.66e-3 | 1.76e-3 | -0.945 | 0.345 | 1.000 |
| Platelet count | 2.25e-3 | 1.69e-3 | 1.332 | 0.183 | 1.000 |
| Platelet crit | 2.49e-3 | 1.66e-3 | 1.504 | 0.133 | 1.000 |
| Platelet distribution width | 2.15e-3 | 1.73e-3 | 1.238 | 0.216 | 1.000 |
| Red blood cell erythrocyte count | -4.22e-3 | 1.50e-3 | -2.815 | 4.87e-3 | 0.302 |
| Red blood cell erythrocyte distribution width | -0.006 | 1.75e-3 | -3.165 | 1.55e-3 | 0.096 |
| Reticulocyte count | -0.009 | 1.74e-3 | -5.335 | 9.55e-8 | 5.92e-6*** |
| Reticulocyte percentage | -0.009 | 1.77e-3 | -4.823 | 1.41e-6 | 8.75e-5*** |
| White blood cell leukocyte count | 0.010 | 1.75e-3 | 5.584 | 2.36e-8 | 1.46e-6*** |
| ***Liver*** | | | | | |
| Albumin | 3.66e-3 | 1.81e-3 | 2.019 | 0.043 | 1.000 |
| Alkaline phosphatase | 1.37e-3 | 1.73e-3 | 0.791 | 0.429 | 1.000 |
| Alanine aminotransferase | -3.67e-3 | 1.69e-3 | -2.170 | 0.030 | 1.000 |
| Aspartate aminotransferase | 0.010 | 1.72e-3 | 5.699 | 1.21e-8 | 7.48e-7*** |
| Direct bilirubin | 2.09e-3 | 1.87e-3 | 1.121 | 0.262 | 1.000 |
| Gamma glutamyltransferase | -2.31e-3 | 1.72e-3 | -1.343 | 0.179 | 1.000 |
| Total bilirubin | 1.64e-3 | 1.72e-3 | 0.955 | 0.339 | 1.000 |
| ***Renal*** | | | | | |
| Creatinine | -0.014 | 1.43e-3 | -9.844 | 7.36e-23 | 4.56e-21*** |
| Cystatin C | -0.010 | 1.62e-3 | -6.353 | 2.11e-10 | 1.31e-8*** |
| Phosphate | 0.008 | 1.78e-3 | 4.643 | 3.44e-6 | 2.13e-4*** |
| Total protein | 0.011 | 1.84e-3 | 5.949 | 2.70e-9 | 1.68e-7*** |
| Urate | -0.013 | 1.50e-3 | -8.939 | 3.96e-19 | 2.45e-17*** |
| Urea | -0.007 | 1.69e-3 | -4.198 | 2.69e-5 | 1.67e-3** |
| ***Cardiovascular*** | | | | | |
| Apolipoprotein A | 0.007 | 1.70e-3 | 3.969 | 7.21e-5 | 4.47e-3** |
| Apolipoprotein B | 1.82e-3 | 1.76e-3 | 1.036 | 0.300 | 1.000 |
| Cholesterol | 4.99e-3 | 1.73e-3 | 2.887 | 3.89e-3 | 0.241 |
| LDL direct | 2.26e-3 | 1.75e-3 | 1.287 | 0.198 | 1.000 |
| Lipoprotein A | 0.006 | 1.98e-3 | 3.024 | 2.50e-3 | 0.155 |
| ***Other Non-BBB-Permeable*** | | | | | |
| Calcium | -3.93e-3 | 1.84e-3 | -2.140 | 0.032 | 1.000 |
| Glycated hemoglobin | -0.007 | 1.71e-3 | -4.239 | 2.24e-5 | 1.39e-3** |
| Rheumatoid factor | -0.005 | 1.78e-3 | -2.998 | 2.71e-3 | 0.168 |

**Table S6.** Peripheral marker minimally adjusted associations with polygenic risk scores (PRS) at threshold ≤ 0.5 for bipolar disorder. BBB, blood-brain barrier; CRP, C-reactive protein; HDL, high-density lipoprotein; IGF-1, insulin-like growth factor 1; LDL, low-density lipoprotein; NLR, neutrophil-to-lymphocyte ratio; p-uncorr., p-uncorrected value; p-corr., Bonferroni p-corrected value; S.E., standard error; SHBG, sex hormone binding globulin. ***p corr. ≤ 0.001, **p corr. ≤ 0.01, *p corr. ≤ 0.05

| **Biomarker** | **Effect Size (β)** | **S.E.** | **t statistic** | **p-uncorr.** | **p-corr.** |
| --- | --- | --- | --- | --- | --- |
| ***BBB-Permeable*** | | | | | |
| Estradiol | 2.52e-3 | 1.65e-3 | 1.532 | 0.126 | 1.000 |
| Glucose | -3.58e-3 | 1.82e-3 | -1.967 | 0.049 | 1.000 |
| HDL cholesterol | 0.010 | 1.67e-3 | 5.789 | 7.07e-9 | 4.38e-7*** |
| IGF-1 | -8.87e-4 | 1.70e-3 | -0.523 | 0.601 | 1.000 |
| SHBG | 3.76e-3 | 1.68e-3 | 2.231 | 0.026 | 1.000 |
| Testosterone | -5.93e-4 | 8.09e-4 | -0.733 | 0.464 | 1.000 |
| Triglycerides | -1.97e-3 | 1.72e-3 | -1.146 | 0.252 | 1.000 |
| Vitamin D | -0.006 | 1.79e-3 | -3.571 | 3.55e-4 | 0.022* |
| ***Inflammatory and Hematological*** | | | | | |
| Basophil count | -2.05e-3 | 1.72e-3 | -1.190 | 0.234 | 1.000 |
| Basophil percentage | -1.05e-3 | 1.75e-3 | -0.600 | 0.548 | 1.000 |
| CRP | -2.93e-3 | 1.76e-3 | -1.665 | 0.096 | 1.000 |
| Eosinophil count | 3.44e-3 | 1.74e-3 | 1.976 | 0.048 | 1.000 |
| Eosinophil percentage | 2.18e-3 | 1.74e-3 | 1.253 | 0.210 | 1.000 |
| Hematocrit percentage | -1.73e-3 | 1.42e-3 | -1.220 | 0.223 | 1.000 |
| Hemoglobin concentration | -9.24e-4 | 1.39e-3 | -0.666 | 0.505 | 1.000 |
| High light scatter reticulocyte count | -0.010 | 1.74e-3 | -5.783 | 7.33e-9 | 4.54e-7*** |
| High light scatter reticulocyte percentage | -0.009 | 1.76e-3 | -5.300 | 1.16e-7 | 7.19e-6*** |
| Immature reticulocyte fraction | -0.006 | 1.75e-3 | -3.626 | 2.88e-4 | 0.018* |
| Lymphocyte count | 0.006 | 1.73e-3 | 3.321 | 8.96e-4 | 0.056 |
| Lymphocyte percentage | 2.77e-3 | 1.73e-3 | 1.606 | 0.108 | 1.000 |
| Mean corpuscular hemoglobin | 0.007 | 1.73e-3 | 4.137 | 3.52e-5 | 2.18e-3** |
| Mean corpuscular hemoglobin concentration | 1.85e-3 | 1.73e-3 | 1.073 | 0.283 | 1.000 |
| Mean corpuscular volume | 0.007 | 1.73e-3 | 4.163 | 3.15e-5 | 1.95e-3** |
| Mean platelet thrombocyte volume | -1.94e-3 | 1.74e-3 | -1.115 | 0.265 | 1.000 |
| Mean reticulocyte volume | 2.81e-3 | 1.75e-3 | 1.611 | 0.107 | 1.000 |
| Mean sphered cell volume | 0.007 | 1.75e-3 | 4.047 | 5.20e-5 | 3.22e-3** |
| Monocyte count | 1.45e-3 | 1.69e-3 | 0.856 | 0.392 | 1.000 |
| Monocyte percentage | -1.62e-3 | 1.69e-3 | -0.960 | 0.337 | 1.000 |
| Neutrophil count | 1.54e-3 | 1.74e-3 | 0.887 | 0.375 | 1.000 |
| Neutrophil percentage | -2.16e-3 | 1.74e-3 | -1.239 | 0.215 | 1.000 |
| NLR | -2.41e-3 | 1.74e-3 | -1.391 | 0.164 | 1.000 |
| Nucleated red blood cell count | -5.47e-4 | 1.75e-3 | -0.312 | 0.755 | 1.000 |
| Nucleated red blood cell percentage | -3.57e-4 | 1.75e-3 | -0.204 | 0.838 | 1.000 |
| Platelet count | -2.09e-3 | 1.68e-3 | -1.246 | 0.213 | 1.000 |
| Platelet crit | -3.39e-3 | 1.65e-3 | -2.053 | 0.040 | 1.000 |
| Platelet distribution width | -7.27e-4 | 1.73e-3 | -0.421 | 0.674 | 1.000 |
| Red blood cell erythrocyte count | -0.005 | 1.49e-3 | -3.423 | 6.20e-4 | 0.038* |
| Red blood cell erythrocyte distribution width | -4.67e-3 | 1.74e-3 | -2.684 | 0.007 | 0.451 |
| Reticulocyte count | -0.009 | 1.74e-3 | -5.389 | 7.07e-8 | 4.39e-6*** |
| Reticulocyte percentage | -0.008 | 1.76e-3 | -4.777 | 1.78e-6 | 1.10e-4*** |
| White blood cell leukocyte count | 3.28e-3 | 1.74e-3 | 1.888 | 0.059 | 1.000 |
| ***Liver*** | | | | | |
| Albumin | -6.18e-4 | 1.80e-3 | -0.343 | 0.732 | 1.000 |
| Alkaline phosphatase | -0.007 | 1.72e-3 | -4.344 | 1.40e-5 | 8.67e-4*** |
| Alanine aminotransferase | -2.01e-3 | 1.68e-3 | -1.194 | 0.233 | 1.000 |
| Aspartate aminotransferase | 0.006 | 1.71e-3 | 3.520 | 4.32e-4 | 0.027* |
| Direct bilirubin | -1.64e-3 | 1.85e-3 | -0.884 | 0.377 | 1.000 |
| Gamma glutamyltransferase | -1.44e-3 | 1.71e-3 | -0.844 | 0.399 | 1.000 |
| Total bilirubin | -9.93e-4 | 1.71e-3 | -0.581 | 0.561 | 1.000 |
| ***Renal*** | | | | | |
| Creatinine | -0.012 | 1.43e-3 | -8.674 | 4.18e-18 | 2.59e-16*** |
| Cystatin C | -0.014 | 1.61e-3 | -8.533 | 1.43e-17 | 8.89e-16*** |
| Phosphate | 0.009 | 1.77e-3 | 4.931 | 8.18e-7 | 5.07e-5*** |
| Total protein | -2.43e-3 | 1.83e-3 | -1.325 | 0.185 | 1.000 |
| Urate | -0.006 | 1.49e-3 | -4.328 | 1.51e-5 | 9.33e-4*** |
| Urea | -0.005 | 1.69e-3 | -3.182 | 1.46e-3 | 0.091 |
| ***Cardiovascular*** | | | | | |
| Apolipoprotein A | 0.006 | 1.69e-3 | 3.399 | 6.77e-4 | 0.042* |
| Apolipoprotein B | -9.65e-4 | 1.75e-3 | -0.551 | 0.582 | 1.000 |
| Cholesterol | 3.08e-3 | 1.72e-3 | 1.793 | 0.073 | 1.000 |
| LDL direct | 5.54e-4 | 1.74e-3 | 0.318 | 0.751 | 1.000 |
| Lipoprotein A | 8.48e-4 | 1.97e-3 | 0.430 | 0.667 | 1.000 |
| ***Other Non-BBB-Permeable*** | | | | | |
| Calcium | -0.007 | 1.83e-3 | -3.686 | 2.28e-4 | 0.014* |
| Glycated hemoglobin | 4.01e-4 | 1.70e-3 | 0.235 | 0.814 | 1.000 |
| Rheumatoid factor | -1.75e-3 | 1.77e-3 | -0.988 | 0.323 | 1.000 |

**Table S7.** Peripheral marker fully adjusted associations with polygenic risk scores (PRS) at threshold ≤ 0.5 for major depressive disorder. BBB, blood-brain barrier; CRP, C-reactive protein; HDL, high-density lipoprotein; IGF-1, insulin-like growth factor 1; LDL, low-density lipoprotein; NLR, neutrophil-to-lymphocyte ratio; p-uncorr., p-uncorrected value; p-corr., Bonferroni p-corrected value; S.E., standard error; SHBG, sex hormone binding globulin. ***p corr. ≤ 0.001, **p corr. ≤ 0.01, *p corr. ≤ 0.05

| **Biomarker** | **Effect Size (β)** | **S.E.** | **t statistic** | **p-uncorr.** | **p-corr.** |
| --- | --- | --- | --- | --- | --- |
| ***BBB-Permeable*** | | | | | |
| Estradiol | -3.28e-4 | 1.59e-3 | -0.206 | 0.837 | 1.000 |
| Glucose | 2.86e-3 | 1.74e-3 | 1.646 | 0.100 | 1.000 |
| HDL cholesterol | -2.18e-3 | 1.49e-3 | -1.456 | 0.145 | 1.000 |
| IGF-1 | -0.010 | 1.62e-3 | -5.854 | 4.80e-9 | 2.98e-7*** |
| SHBG | 2.20e-3 | 1.51e-3 | 1.457 | 0.145 | 1.000 |
| Testosterone | -1.48e-4 | 7.68e-4 | -0.193 | 0.847 | 1.000 |
| Triglycerides | 0.010 | 1.58e-3 | 6.476 | 9.40e-11 | 5.83e-9*** |
| Vitamin D | -0.010 | 1.70e-3 | -5.948 | 2.71e-9 | 1.68e-7*** |
| ***Inflammatory and Hematological*** | | | | | |
| Basophil count | 2.72e-3 | 1.65e-3 | 1.645 | 0.100 | 1.000 |
| Basophil percentage | -6.73e-4 | 1.69e-3 | -0.399 | 0.690 | 1.000 |
| CRP | 0.006 | 1.59e-3 | 3.727 | 1.94e-4 | 0.012* |
| Eosinophil count | 4.34e-4 | 1.67e-3 | 0.260 | 0.795 | 1.000 |
| Eosinophil percentage | -4.18e-3 | 1.68e-3 | -2.486 | 0.013 | 0.801 |
| Hematocrit percentage | -2.80e-3 | 1.36e-3 | -2.053 | 0.040 | 1.000 |
| Hemoglobin concentration | -2.18e-3 | 1.33e-3 | -1.639 | 0.101 | 1.000 |
| High light scatter reticulocyte count | 4.20e-3 | 1.54e-3 | 2.730 | 0.006 | 0.393 |
| High light scatter reticulocyte percentage | 4.71e-3 | 1.56e-3 | 3.015 | 2.57e-3 | 0.159 |
| Immature reticulocyte fraction | 4.33e-3 | 1.60e-3 | 2.700 | 0.007 | 0.430 |
| Lymphocyte count | 0.006 | 1.62e-3 | 3.844 | 1.21e-4 | 0.008** |
| Lymphocyte percentage | -0.006 | 1.67e-3 | -3.327 | 8.77e-4 | 0.054 |
| Mean corpuscular hemoglobin | 2.22e-3 | 1.64e-3 | 1.355 | 0.175 | 1.000 |
| Mean corpuscular hemoglobin concentration | 1.54e-3 | 1.67e-3 | 0.925 | 0.355 | 1.000 |
| Mean corpuscular volume | 2.03e-3 | 1.64e-3 | 1.241 | 0.215 | 1.000 |
| Mean platelet thrombocyte volume | 1.87e-3 | 1.68e-3 | 1.111 | 0.267 | 1.000 |
| Mean reticulocyte volume | 3.05e-3 | 1.68e-3 | 1.816 | 0.069 | 1.000 |
| Mean sphered cell volume | 2.03e-3 | 1.65e-3 | 1.227 | 0.220 | 1.000 |
| Monocyte count | 4.59e-3 | 1.61e-3 | 2.848 | 4.41e-3 | 0.273 |
| Monocyte percentage | -4.62e-3 | 1.63e-3 | -2.836 | 4.57e-3 | 0.283 |
| Neutrophil count | 0.012 | 1.63e-3 | 7.663 | 1.82e-14 | 1.13e-12*** |
| Neutrophil percentage | 0.007 | 1.68e-3 | 3.906 | 9.40e-5 | 0.006** |
| NLR | 0.006 | 1.68e-3 | 3.620 | 2.95e-4 | 0.018* |
| Nucleated red blood cell count | 1.72e-3 | 1.69e-3 | 1.017 | 0.309 | 1.000 |
| Nucleated red blood cell percentage | 1.85e-3 | 1.69e-3 | 1.097 | 0.273 | 1.000 |
| Platelet count | 2.33e-3 | 1.62e-3 | 1.437 | 0.151 | 1.000 |
| Platelet crit | 3.56e-3 | 1.59e-3 | 2.242 | 0.025 | 1.000 |
| Platelet distribution width | 3.79e-3 | 1.67e-3 | 2.272 | 0.023 | 1.000 |
| Red blood cell erythrocyte count | -3.67e-3 | 1.42e-3 | -2.586 | 0.010 | 0.601 |
| Red blood cell erythrocyte distribution width | 6.57e-4 | 1.67e-3 | 0.394 | 0.694 | 1.000 |
| Reticulocyte count | 3.48e-3 | 1.56e-3 | 2.233 | 0.026 | 1.000 |
| Reticulocyte percentage | 4.61e-3 | 1.59e-3 | 2.895 | 3.79e-3 | 0.235 |
| White blood cell leukocyte count | 0.013 | 1.59e-3 | 7.888 | 3.09e-15 | 1.91e-13*** |
| ***Liver*** | | | | | |
| Albumin | 4.82e-4 | 1.72e-3 | 0.280 | 0.779 | 1.000 |
| Alkaline phosphatase | 2.22e-3 | 1.64e-3 | 1.359 | 0.174 | 1.000 |
| Alanine aminotransferase | 1.95e-3 | 1.56e-3 | 1.249 | 0.212 | 1.000 |
| Aspartate aminotransferase | 0.005 | 1.64e-3 | 3.199 | 1.38e-3 | 0.085 |
| Direct bilirubin | -2.12e-3 | 1.78e-3 | -1.188 | 0.235 | 1.000 |
| Gamma glutamyltransferase | 0.007 | 1.61e-3 | 4.272 | 1.94e-5 | 1.20e-3** |
| Total bilirubin | -0.007 | 1.64e-3 | -4.111 | 3.93e-5 | 2.44e-3** |
| ***Renal*** | | | | | |
| Creatinine | -0.008 | 1.37e-3 | -5.897 | 3.71e-9 | 2.30e-7*** |
| Cystatin C | 8.68e-4 | 1.46e-3 | 0.595 | 0.552 | 1.000 |
| Phosphate | -1.22e-4 | 1.71e-3 | -0.072 | 0.943 | 1.000 |
| Total protein | 8.27e-4 | 1.77e-3 | 0.468 | 0.640 | 1.000 |
| Urate | -3.94e-3 | 1.33e-3 | -2.954 | 3.14e-3 | 0.195 |
| Urea | -0.006 | 1.62e-3 | -3.522 | 4.29e-4 | 0.027* |
| ***Cardiovascular*** | | | | | |
| Apolipoprotein A | -1.55e-3 | 1.56e-3 | -0.995 | 0.320 | 1.000 |
| Apolipoprotein B | -3.05e-3 | 1.69e-3 | -1.811 | 0.070 | 1.000 |
| Cholesterol | -3.44e-3 | 1.66e-3 | -2.074 | 0.038 | 1.000 |
| LDL direct | -4.50e-3 | 1.68e-3 | -2.672 | 0.008 | 0.468 |
| Lipoprotein A | -4.60e-4 | 1.90e-3 | -0.242 | 0.809 | 1.000 |
| ***Other Non-BBB-Permeable*** | | | | | |
| Calcium | 4.96e-3 | 1.76e-3 | 2.812 | 4.93e-3 | 0.305 |
| Glycated hemoglobin | 2.90e-3 | 1.59e-3 | 1.827 | 0.068 | 1.000 |
| Rheumatoid factor | -3.47e-3 | 1.71e-3 | -2.027 | 0.043 | 1.000 |

**Table S8.** Peripheral marker fully adjusted associations with polygenic risk scores (PRS) at threshold ≤ 0.5 for schizophrenia. BBB, blood-brain barrier; CRP, C-reactive protein; HDL, high-density lipoprotein; IGF-1, insulin-like growth factor 1; LDL, low-density lipoprotein; NLR, neutrophil-to-lymphocyte ratio; p-uncorr., p-uncorrected value; p-corr., Bonferroni p-corrected value; S.E., standard error; SHBG, sex hormone binding globulin. ***p corr. ≤ 0.001, **p corr. ≤ 0.01, *p corr. ≤ 0.05

| **Biomarker** | **Effect Size (β)** | **S.E.** | **t statistic** | **p-uncorr.** | **p-corr.** |
| --- | --- | --- | --- | --- | --- |
| ***BBB-Permeable*** | | | | | |
| Estradiol | 3.89e-3 | 1.66e-3 | 2.345 | 0.019 | 1.000 |
| Glucose | -3.84e-3 | 1.81e-3 | -2.121 | 0.034 | 1.000 |
| HDL cholesterol | 4.82e-3 | 1.56e-3 | 3.093 | 1.98e-3 | 0.123 |
| IGF-1 | 2.91e-3 | 1.69e-3 | 1.722 | 0.085 | 1.000 |
| SHBG | 0.005 | 1.57e-3 | 3.266 | 1.09e-3 | 0.068 |
| Testosterone | 2.81e-3 | 8.01e-4 | 3.513 | 4.43e-4 | 0.027* |
| Triglycerides | -9.09e-4 | 1.65e-3 | -0.550 | 0.582 | 1.000 |
| Vitamin D | -0.017 | 1.77e-3 | -9.504 | 2.03e-21 | 1.26e-19*** |
| ***Inflammatory and Hematological*** | | | | | |
| Basophil count | 4.45e-3 | 1.72e-3 | 2.584 | 0.010 | 0.606 |
| Basophil percentage | 4.71e-3 | 1.76e-3 | 2.683 | 0.007 | 0.452 |
| CRP | -1.32e-3 | 1.66e-3 | -0.796 | 0.426 | 1.000 |
| Eosinophil count | 0.009 | 1.74e-3 | 5.057 | 4.26e-7 | 2.64e-5*** |
| Eosinophil percentage | 0.006 | 1.75e-3 | 3.269 | 1.08e-3 | 0.067 |
| Hematocrit percentage | 2.14e-3 | 1.42e-3 | 1.512 | 0.131 | 1.000 |
| Hemoglobin concentration | 3.01e-3 | 1.38e-3 | 2.174 | 0.030 | 1.000 |
| High light scatter reticulocyte count | -1.55e-3 | 1.60e-3 | -0.970 | 0.332 | 1.000 |
| High light scatter reticulocyte percentage | -1.81e-3 | 1.63e-3 | -1.113 | 0.266 | 1.000 |
| Immature reticulocyte fraction | -1.98e-3 | 1.67e-3 | -1.186 | 0.236 | 1.000 |
| Lymphocyte count | 0.015 | 1.69e-3 | 8.880 | 6.69e-19 | 4.15e-17*** |
| Lymphocyte percentage | 0.009 | 1.74e-3 | 5.214 | 1.85e-7 | 1.15e-5*** |
| Mean corpuscular hemoglobin | 3.04e-3 | 1.71e-3 | 1.784 | 0.074 | 1.000 |
| Mean corpuscular hemoglobin concentration | 1.94e-3 | 1.74e-3 | 1.114 | 0.265 | 1.000 |
| Mean corpuscular volume | 2.22e-3 | 1.70e-3 | 1.303 | 0.193 | 1.000 |
| Mean platelet thrombocyte volume | -7.23e-4 | 1.75e-3 | -0.412 | 0.680 | 1.000 |
| Mean reticulocyte volume | 2.75e-3 | 1.75e-3 | 1.574 | 0.115 | 1.000 |
| Mean sphered cell volume | 4.41e-3 | 1.72e-3 | 2.562 | 0.010 | 0.645 |
| Monocyte count | 0.005 | 1.68e-3 | 3.020 | 2.53e-3 | 0.157 |
| Monocyte percentage | -2.32e-3 | 1.70e-3 | -1.367 | 0.172 | 1.000 |
| Neutrophil count | 3.72e-3 | 1.70e-3 | 2.196 | 0.028 | 1.000 |
| Neutrophil percentage | -0.009 | 1.75e-3 | -5.049 | 4.45e-7 | 2.76e-5*** |
| NLR | -0.008 | 1.75e-3 | -4.626 | 3.74e-6 | 2.32e-4*** |
| Nucleated red blood cell count | -2.57e-3 | 1.76e-3 | -1.460 | 0.144 | 1.000 |
| Nucleated red blood cell percentage | -1.76e-3 | 1.76e-3 | -0.999 | 0.318 | 1.000 |
| Platelet count | 2.50e-3 | 1.69e-3 | 1.480 | 0.139 | 1.000 |
| Platelet crit | 2.90e-3 | 1.65e-3 | 1.755 | 0.079 | 1.000 |
| Platelet distribution width | 3.37e-3 | 1.74e-3 | 1.941 | 0.052 | 1.000 |
| Red blood cell erythrocyte count | 6.24e-4 | 1.48e-3 | 0.422 | 0.673 | 1.000 |
| Red blood cell erythrocyte distribution width | -4.57e-3 | 1.74e-3 | -2.627 | 0.009 | 0.533 |
| Reticulocyte count | -1.06e-3 | 1.62e-3 | -0.653 | 0.514 | 1.000 |
| Reticulocyte percentage | -1.06e-3 | 1.66e-3 | -0.640 | 0.522 | 1.000 |
| White blood cell leukocyte count | 0.009 | 1.66e-3 | 5.629 | 1.82e-8 | 1.13e-6*** |
| ***Liver*** | | | | | |
| Albumin | 1.17e-3 | 1.79e-3 | 0.651 | 0.515 | 1.000 |
| Alkaline phosphatase | 3.25e-3 | 1.71e-3 | 1.904 | 0.057 | 1.000 |
| Alanine aminotransferase | 2.97e-3 | 1.63e-3 | 1.828 | 0.068 | 1.000 |
| Aspartate aminotransferase | 0.013 | 1.71e-3 | 7.565 | 3.89e-14 | 2.41e-12*** |
| Direct bilirubin | 2.05e-3 | 1.86e-3 | 1.103 | 0.270 | 1.000 |
| Gamma glutamyltransferase | 7.78e-4 | 1.68e-3 | 0.462 | 0.644 | 1.000 |
| Total bilirubin | 1.95e-3 | 1.70e-3 | 1.145 | 0.252 | 1.000 |
| ***Renal*** | | | | | |
| Creatinine | -0.011 | 1.43e-3 | -7.808 | 5.85e-15 | 3.63e-13*** |
| Cystatin C | -0.006 | 1.52e-3 | -3.993 | 6.54e-5 | 4.05e-3** |
| Phosphate | 4.96e-3 | 1.78e-3 | 2.787 | 0.005 | 0.330 |
| Total protein | 0.013 | 1.84e-3 | 7.207 | 5.74e-13 | 3.56e-11*** |
| Urate | -0.006 | 1.39e-3 | -4.388 | 1.14e-5 | 7.08e-4*** |
| Urea | -3.71e-3 | 1.69e-3 | -2.198 | 0.028 | 1.000 |
| ***Cardiovascular*** | | | | | |
| Apolipoprotein A | 1.72e-3 | 1.63e-3 | 1.055 | 0.291 | 1.000 |
| Apolipoprotein B | 3.89e-3 | 1.76e-3 | 2.213 | 0.027 | 1.000 |
| Cholesterol | 4.94e-3 | 1.73e-3 | 2.861 | 4.22e-3 | 0.262 |
| LDL direct | 3.65e-3 | 1.75e-3 | 2.083 | 0.037 | 1.000 |
| Lipoprotein A | 0.006 | 1.99e-3 | 2.962 | 3.06e-3 | 0.190 |
| ***Other Non-BBB-Permeable*** | | | | | |
| Calcium | -4.74e-3 | 1.84e-3 | -2.575 | 0.010 | 0.622 |
| Glycated hemoglobin | -3.90e-3 | 1.65e-3 | -2.359 | 0.018 | 1.000 |
| Rheumatoid factor | -0.006 | 1.79e-3 | -3.098 | 1.95e-3 | 0.121 |

**Table S9.** Peripheral marker fully adjusted associations with polygenic risk scores (PRS) at threshold ≤ 0.5 for bipolar disorder. BBB, blood-brain barrier; CRP, C-reactive protein; HDL, high-density lipoprotein; IGF-1, insulin-like growth factor 1; LDL, low-density lipoprotein; NLR, neutrophil-to-lymphocyte ratio; p-uncorr., p-uncorrected value; p-corr., Bonferroni p-corrected value; S.E., standard error; SHBG, sex hormone binding globulin. ***p corr. ≤ 0.001, **p corr. ≤ 0.01, *p corr. ≤ 0.05

| **Biomarker** | **Effect Size (β)** | **S.E.** | **t statistic** | **p-uncorr.** | **p-corr.** |
| --- | --- | --- | --- | --- | --- |
| ***BBB-Permeable*** | | | | | |
| Estradiol | 2.36e-3 | 1.65e-3 | 1.431 | 0.152 | 1.000 |
| Glucose | -1.90e-3 | 1.80e-3 | -1.056 | 0.291 | 1.000 |
| HDL cholesterol | 0.007 | 1.55e-3 | 4.230 | 2.33e-5 | 1.45e-3** |
| IGF-1 | -1.63e-3 | 1.68e-3 | -0.970 | 0.332 | 1.000 |
| SHBG | -4.30e-4 | 1.57e-3 | -0.275 | 0.784 | 1.000 |
| Testosterone | -1.44e-3 | 7.96e-4 | -1.805 | 0.071 | 1.000 |
| Triglycerides | 6.60e-4 | 1.64e-3 | 0.402 | 0.688 | 1.000 |
| Vitamin D | -0.008 | 1.76e-3 | -4.432 | 9.35e-6 | 5.80e-4*** |
| ***Inflammatory and Hematological*** | | | | | |
| Basophil count | -2.36e-3 | 1.71e-3 | -1.379 | 0.168 | 1.000 |
| Basophil percentage | -1.44e-3 | 1.75e-3 | -0.822 | 0.411 | 1.000 |
| CRP | -2.05e-4 | 1.65e-3 | -0.125 | 0.901 | 1.000 |
| Eosinophil count | 3.70e-3 | 1.73e-3 | 2.137 | 0.033 | 1.000 |
| Eosinophil percentage | 2.39e-3 | 1.74e-3 | 1.373 | 0.170 | 1.000 |
| Hematocrit percentage | -5.11e-4 | 1.41e-3 | -0.362 | 0.717 | 1.000 |
| Hemoglobin concentration | 3.28e-4 | 1.38e-3 | 0.238 | 0.812 | 1.000 |
| High light scatter reticulocyte count | -0.006 | 1.59e-3 | -3.518 | 4.34e-4 | 0.027* |
| High light scatter reticulocyte percentage | -0.005 | 1.62e-3 | -3.254 | 1.14e-3 | 0.071 |
| Immature reticulocyte fraction | -3.56e-3 | 1.66e-3 | -2.142 | 0.032 | 1.000 |
| Lymphocyte count | 0.006 | 1.68e-3 | 3.587 | 3.34e-4 | 0.021* |
| Lymphocyte percentage | 2.96e-3 | 1.73e-3 | 1.710 | 0.087 | 1.000 |
| Mean corpuscular hemoglobin | 4.43e-3 | 1.70e-3 | 2.612 | 0.009 | 0.558 |
| Mean corpuscular hemoglobin concentration | 1.94e-3 | 1.73e-3 | 1.120 | 0.263 | 1.000 |
| Mean corpuscular volume | 4.14e-3 | 1.69e-3 | 2.444 | 0.015 | 0.900 |
| Mean platelet thrombocyte volume | -1.80e-3 | 1.75e-3 | -1.029 | 0.304 | 1.000 |
| Mean reticulocyte volume | 1.03e-3 | 1.74e-3 | 0.592 | 0.554 | 1.000 |
| Mean sphered cell volume | 3.61e-3 | 1.71e-3 | 2.110 | 0.035 | 1.000 |
| Monocyte count | 1.83e-3 | 1.67e-3 | 1.096 | 0.273 | 1.000 |
| Monocyte percentage | -1.35e-3 | 1.69e-3 | -0.801 | 0.423 | 1.000 |
| Neutrophil count | 1.47e-3 | 1.69e-3 | 0.871 | 0.384 | 1.000 |
| Neutrophil percentage | -2.43e-3 | 1.75e-3 | -1.393 | 0.164 | 1.000 |
| NLR | -2.84e-3 | 1.74e-3 | -1.635 | 0.102 | 1.000 |
| Nucleated red blood cell count | -3.18e-4 | 1.75e-3 | -0.181 | 0.856 | 1.000 |
| Nucleated red blood cell percentage | -4.64e-5 | 1.75e-3 | -0.026 | 0.979 | 1.000 |
| Platelet count | -2.02e-3 | 1.68e-3 | -1.205 | 0.228 | 1.000 |
| Platelet crit | -3.21e-3 | 1.65e-3 | -1.951 | 0.051 | 1.000 |
| Platelet distribution width | -1.10e-5 | 1.73e-3 | -0.006 | 0.995 | 1.000 |
| Red blood cell erythrocyte count | -2.37e-3 | 1.47e-3 | -1.613 | 0.107 | 1.000 |
| Red blood cell erythrocyte distribution width | -4.24e-3 | 1.73e-3 | -2.450 | 0.014 | 0.887 |
| Reticulocyte count | -0.005 | 1.62e-3 | -3.157 | 1.59e-3 | 0.099 |
| Reticulocyte percentage | -4.58e-3 | 1.65e-3 | -2.778 | 0.005 | 0.339 |
| White blood cell leukocyte count | 3.34e-3 | 1.65e-3 | 2.022 | 0.043 | 1.000 |
| ***Liver*** | | | | | |
| Albumin | -1.74e-3 | 1.78e-3 | -0.974 | 0.330 | 1.000 |
| Alkaline phosphatase | -0.006 | 1.70e-3 | -3.797 | 1.47e-4 | 0.009** |
| Alanine aminotransferase | 1.31e-3 | 1.62e-3 | 0.812 | 0.417 | 1.000 |
| Aspartate aminotransferase | 0.008 | 1.70e-3 | 4.433 | 9.32e-6 | 5.78e-4*** |
| Direct bilirubin | -1.48e-3 | 1.85e-3 | -0.803 | 0.422 | 1.000 |
| Gamma glutamyltransferase | 1.26e-4 | 1.67e-3 | 0.075 | 0.940 | 1.000 |
| Total bilirubin | -7.29e-4 | 1.69e-3 | -0.430 | 0.667 | 1.000 |
| ***Renal*** | | | | | |
| Creatinine | -0.011 | 1.42e-3 | -7.571 | 3.70e-14 | 2.29e-12*** |
| Cystatin C | -0.012 | 1.51e-3 | -7.830 | 4.91e-15 | 3.04e-13*** |
| Phosphate | 0.007 | 1.77e-3 | 3.942 | 8.08e-5 | 0.005** |
| Total protein | -1.26e-3 | 1.83e-3 | -0.689 | 0.491 | 1.000 |
| Urate | -2.86e-3 | 1.38e-3 | -2.068 | 0.039 | 1.000 |
| Urea | -3.71e-3 | 1.68e-3 | -2.210 | 0.027 | 1.000 |
| ***Cardiovascular*** | | | | | |
| Apolipoprotein A | 3.59e-3 | 1.62e-3 | 2.218 | 0.027 | 1.000 |
| Apolipoprotein B | 3.81e-4 | 1.75e-3 | 0.218 | 0.828 | 1.000 |
| Cholesterol | 3.44e-3 | 1.72e-3 | 2.000 | 0.046 | 1.000 |
| LDL direct | 1.56e-3 | 1.74e-3 | 0.896 | 0.371 | 1.000 |
| Lipoprotein A | 8.54e-4 | 1.97e-3 | 0.433 | 0.665 | 1.000 |
| ***Other Non-BBB-Permeable*** | | | | | |
| Calcium | -0.007 | 1.83e-3 | -3.919 | 8.89e-5 | 0.006** |
| Glycated hemoglobin | 1.93e-3 | 1.64e-3 | 1.173 | 0.241 | 1.000 |
| Rheumatoid factor | -2.07e-3 | 1.78e-3 | -1.168 | 0.243 | 1.000 |

***Sensitivity analysis of major depressive disorder polygenic risk score associations***

**Table S10.** Results from sensitivity analysis of major depressive disorder (MDD) peripheral marker fully adjusted associations with MDD polygenic risk scores (PRS) at threshold ≤ 0.5, excluding patients. BBB, blood-brain barrier; CRP, C-reactive protein; HDL, high-density lipoprotein; IGF-1, insulin-like growth factor 1; LDL, low-density lipoprotein; NLR, neutrophil-to-lymphocyte ratio; p-uncorr., p-uncorrected value; p-corr., Bonferroni p-corrected value; S.E., standard error; SHBG, sex hormone binding globulin. ***p corr. ≤ 0.001, **p corr. ≤ 0.01, *p corr. ≤ 0.05

| **Biomarker** | **Effect Size (β)** | **S.E.** | **t statistic** | **p-uncorr.** | **p-corr.** |
| --- | --- | --- | --- | --- | --- |
| ***BBB-Permeable*** | | | | | |
| Estradiol | -4.74e-4 | 1.61e-3 | -0.294 | 0.769 | 1.000 |
| Glucose | 2.52e-3 | 1.76e-3 | 1.433 | 0.152 | 1.000 |
| HDL cholesterol | -1.86e-3 | 1.51e-3 | -1.226 | 0.220 | 1.000 |
| IGF-1 | -0.009 | 1.65e-3 | -5.330 | 9.85e-8 | 6.10e-6*** |
| SHBG | 1.78e-3 | 1.53e-3 | 1.164 | 0.244 | 1.000 |
| Testosterone | -1.86e-4 | 7.78e-4 | -0.240 | 0.811 | 1.000 |
| Triglycerides | 0.010 | 1.61e-3 | 5.933 | 2.98e-9 | 1.85e-7*** |
| Vitamin D | -0.010 | 1.72e-3 | -5.618 | 1.93e-8 | 1.20e-6*** |
| ***Inflammatory and Hematological*** | | | | | |
| Basophil count | 1.77e-3 | 1.68e-3 | 1.056 | 0.291 | 1.000 |
| Basophil percentage | -1.81e-3 | 1.71e-3 | -1.061 | 0.289 | 1.000 |
| CRP | 4.74e-3 | 1.62e-3 | 2.932 | 3.37e-3 | 0.209 |
| Eosinophil count | 2.21e-4 | 1.69e-3 | 0.131 | 0.896 | 1.000 |
| Eosinophil percentage | -4.25e-3 | 1.71e-3 | -2.490 | 0.013 | 0.791 |
| Hematocrit percentage | -2.25e-3 | 1.38e-3 | -1.634 | 0.102 | 1.000 |
| Hemoglobin concentration | -1.51e-3 | 1.34e-3 | -1.126 | 0.260 | 1.000 |
| High light scatter reticulocyte count | 3.61e-3 | 1.56e-3 | 2.315 | 0.021 | 1.000 |
| High light scatter reticulocyte percentage | 3.89e-3 | 1.59e-3 | 2.452 | 0.014 | 0.881 |
| Immature reticulocyte fraction | 3.39e-3 | 1.63e-3 | 2.085 | 0.037 | 1.000 |
| Lymphocyte count | 0.007 | 1.65e-3 | 4.041 | 5.33e-5 | 3.31e-3** |
| Lymphocyte percentage | -4.90e-3 | 1.69e-3 | -2.894 | 3.81e-3 | 0.236 |
| Mean corpuscular hemoglobin | 1.86e-3 | 1.66e-3 | 1.120 | 0.263 | 1.000 |
| Mean corpuscular hemoglobin concentration | 1.93e-3 | 1.69e-3 | 1.138 | 0.255 | 1.000 |
| Mean corpuscular volume | 1.36e-3 | 1.66e-3 | 0.820 | 0.412 | 1.000 |
| Mean platelet thrombocyte volume | 2.69e-3 | 1.71e-3 | 1.574 | 0.115 | 1.000 |
| Mean reticulocyte volume | 1.96e-3 | 1.70e-3 | 1.153 | 0.249 | 1.000 |
| Mean sphered cell volume | 1.25e-3 | 1.68e-3 | 0.747 | 0.455 | 1.000 |
| Monocyte count | 4.63e-3 | 1.63e-3 | 2.836 | 4.56e-3 | 0.283 |
| Monocyte percentage | -4.38e-3 | 1.65e-3 | -2.654 | 0.008 | 0.493 |
| Neutrophil count | 0.012 | 1.65e-3 | 7.324 | 2.41e-13 | 1.50e-11** |
| Neutrophil percentage | 0.006 | 1.71e-3 | 3.586 | 3.36e-4 | 0.021* |
| NLR | 0.005 | 1.70e-3 | 3.173 | 1.51e-3 | 0.094 |
| Nucleated red blood cell count | 1.73e-3 | 1.72e-3 | 1.005 | 0.315 | 1.000 |
| Nucleated red blood cell percentage | 1.96e-3 | 1.72e-3 | 1.140 | 0.254 | 1.000 |
| Platelet count | 1.33e-3 | 1.64e-3 | 0.810 | 0.418 | 1.000 |
| Platelet crit | 2.89e-3 | 1.61e-3 | 1.795 | 0.073 | 1.000 |
| Platelet distribution width | 3.98e-3 | 1.69e-3 | 2.354 | 0.019 | 1.000 |
| Red blood cell erythrocyte count | -2.88e-3 | 1.44e-3 | -2.005 | 0.045 | 1.000 |
| Red blood cell erythrocyte distribution width | -3.79e-5 | 1.69e-3 | -0.022 | 0.982 | 1.000 |
| Reticulocyte count | 3.23e-3 | 1.58e-3 | 2.040 | 0.041 | 1.000 |
| Reticulocyte percentage | 4.13e-3 | 1.62e-3 | 2.556 | 0.011 | 0.657 |
| White blood cell leukocyte count | 0.012 | 1.62e-3 | 7.625 | 2.45e-14 | 1.52e-12*** |
| ***Liver*** |  |  |  |  |  |
| Albumin | 1.86e-3 | 1.74e-3 | 1.068 | 0.285 | 1.000 |
| Alkaline phosphatase | 1.41e-3 | 1.66e-3 | 0.849 | 0.396 | 1.000 |
| Alanine aminotransferase | 1.89e-3 | 1.58e-3 | 1.195 | 0.232 | 1.000 |
| Aspartate aminotransferase | 0.005 | 1.67e-3 | 3.022 | 2.51e-3 | 0.156 |
| Direct bilirubin | -2.04e-3 | 1.81e-3 | -1.130 | 0.258 | 1.000 |
| Gamma glutamyltransferase | 0.006 | 1.64e-3 | 3.787 | 1.52e-4 | 0.009** |
| Total bilirubin | -0.006 | 1.66e-3 | -3.775 | 1.60e-4 | 0.010* |
| ***Renal*** | | | | | |
| Creatinine | -0.008 | 1.38e-3 | -5.818 | 5.96e-9 | 3.70e-7*** |
| Cystatin C | 2.85e-4 | 1.48e-3 | 0.193 | 0.847 | 1.000 |
| Phosphate | 2.80e-4 | 1.73e-3 | 0.162 | 0.872 | 1.000 |
| Total protein | 1.75e-3 | 1.79e-3 | 0.978 | 0.328 | 1.000 |
| Urate | -3.94e-3 | 1.35e-3 | -2.919 | 3.51e-3 | 0.218 |
| Urea | -0.005 | 1.64e-3 | -3.306 | 9.46e-4 | 0.059 |
| ***Cardiovascular*** | | | | | |
| Apolipoprotein A | -1.33e-3 | 1.58e-3 | -0.842 | 0.400 | 1.000 |
| Apolipoprotein B | -3.53e-3 | 1.71e-3 | -2.065 | 0.039 | 1.000 |
| Cholesterol | -3.92e-3 | 1.68e-3 | -2.332 | 0.020 | 1.000 |
| LDL direct | -4.94e-3 | 1.71e-3 | -2.894 | 3.80e-3 | 0.236 |
| Lipoprotein A | -4.58e-4 | 1.93e-3 | -0.237 | 0.812 | 1.000 |
| ***Other Non-BBB-Permeable*** | | | | | |
| Calcium | 0.006 | 1.79e-3 | 3.285 | 1.02e-3 | 0.063 |
| Glycated hemoglobin | 2.63e-3 | 1.61e-3 | 1.636 | 0.102 | 1.000 |
| Rheumatoid factor | -3.51e-3 | 1.74e-3 | -2.021 | 0.043 | 1.000 |

***Disorder PRS Associations with blood markers at all other PRS thresholds***

**Table S11.** Peripheral marker minimally adjusted associations with polygenic risk scores (PRS) at threshold ≤ 0.01 for major depressive disorder. BBB, blood-brain barrier; CRP, C-reactive protein; HDL, high-density lipoprotein; IGF-1, insulin-like growth factor 1; LDL, low-density lipoprotein; NLR, neutrophil-to-lymphocyte ratio; p-uncorr., p-uncorrected value; p-corr., Bonferroni p-corrected value; S.E., standard error; SHBG, sex hormone binding globulin. ***p corr. ≤ 0.001, **p corr. ≤ 0.01, *p corr. ≤ 0.05

| **Biomarker** | **Effect Size (β)** | **S.E.** | **t statistic** | **p-uncorr.** | **p-corr.** |
| --- | --- | --- | --- | --- | --- |
| ***BBB-Permeable*** | | | | | |
| Estradiol | -2.13e-4 | 1.58e-3 | -0.135 | 0.893 | 1.000 |
| Glucose | 0.008 | 1.75e-3 | 4.362 | 1.29e-5 | 8.00e-4*** |
| HDL cholesterol | -0.012 | 1.60e-3 | -7.506 | 6.11e-14 | 3.79e-12*** |
| IGF-1 | -0.011 | 1.63e-3 | -6.568 | 5.12e-11 | 3.18e-9*** |
| SHBG | -0.007 | 1.62e-3 | -4.493 | 7.04e-6 | 4.36e-4*** |
| Testosterone | -2.97e-3 | 7.77e-4 | -3.826 | 1.30e-4 | 0.008** |
| Triglycerides | 0.020 | 1.65e-3 | 12.391 | 2.97e-35 | 1.84e-33*** |
| Vitamin D | -0.014 | 1.72e-3 | -8.244 | 1.66e-16 | 1.03e-14*** |
| ***Inflammatory and Hematological*** | | | | | |
| Basophil count | 0.005 | 1.65e-3 | 3.179 | 1.48e-3 | 0.092 |
| Basophil percentage | -1.31e-3 | 1.68e-3 | -0.784 | 0.433 | 1.000 |
| CRP | 0.017 | 1.69e-3 | 10.236 | 1.38e-24 | 8.53e-23*** |
| Eosinophil count | 0.010 | 1.67e-3 | 5.908 | 3.47e-9 | 2.15e-7*** |
| Eosinophil percentage | 1.04e-3 | 1.67e-3 | 0.623 | 0.533 | 1.000 |
| Hematocrit percentage | 1.07e-4 | 1.37e-3 | 0.079 | 0.937 | 1.000 |
| Hemoglobin concentration | 1.61e-3 | 1.33e-3 | 1.213 | 0.225 | 1.000 |
| High light scatter reticulocyte count | 0.022 | 1.67e-3 | 12.932 | 3.05e-38 | 1.89e-36*** |
| High light scatter reticulocyte percentage | 0.021 | 1.69e-3 | 12.608 | 1.94e-36 | 1.20e-34*** |
| Immature reticulocyte fraction | 0.018 | 1.68e-3 | 10.611 | 2.67e-26 | 1.66e-24*** |
| Lymphocyte count | 0.021 | 1.66e-3 | 12.759 | 2.83e-37 | 1.76e-35*** |
| Lymphocyte percentage | -1.07e-3 | 1.66e-3 | -0.647 | 0.517 | 1.000 |
| Mean corpuscular hemoglobin | 1.13e-3 | 1.66e-3 | 0.684 | 0.494 | 1.000 |
| Mean corpuscular hemoglobin concentration | 4.45e-3 | 1.66e-3 | 2.689 | 0.007 | 0.445 |
| Mean corpuscular volume | -8.65e-4 | 1.66e-3 | -0.521 | 0.603 | 1.000 |
| Mean platelet thrombocyte volume | 2.63e-3 | 1.67e-3 | 1.572 | 0.116 | 1.000 |
| Mean reticulocyte volume | -3.16e-3 | 1.67e-3 | -1.888 | 0.059 | 1.000 |
| Mean sphered cell volume | -4.77e-3 | 1.68e-3 | -2.842 | 4.48e-3 | 0.278 |
| Monocyte count | 0.014 | 1.62e-3 | 8.696 | 3.44e-18 | 2.13e-16*** |
| Monocyte percentage | -0.005 | 1.62e-3 | -3.349 | 8.10e-4 | 0.050* |
| Neutrophil count | 0.022 | 1.67e-3 | 13.014 | 1.03e-38 | 6.41e-37*** |
| Neutrophil percentage | 2.23e-3 | 1.67e-3 | 1.333 | 0.183 | 1.000 |
| NLR | 9.19e-4 | 1.67e-3 | 0.552 | 0.581 | 1.000 |
| Nucleated red blood cell count | -1.54e-3 | 1.68e-3 | -0.914 | 0.361 | 1.000 |
| Nucleated red blood cell percentage | -4.05e-4 | 1.68e-3 | -0.241 | 0.809 | 1.000 |
| Platelet count | 4.38e-3 | 1.61e-3 | 2.717 | 0.007 | 0.408 |
| Platelet crit | 0.006 | 1.58e-3 | 4.020 | 5.81e-5 | 3.61e-3** |
| Platelet distribution width | 0.008 | 1.66e-3 | 4.761 | 1.93e-6 | 1.20e-4*** |
| Red blood cell erythrocyte count | 6.70e-4 | 1.43e-3 | 0.468 | 0.640 | 1.000 |
| Red blood cell erythrocyte distribution width | 1.69e-4 | 1.67e-3 | 0.101 | 0.919 | 1.000 |
| Reticulocyte count | 0.019 | 1.67e-3 | 11.469 | 1.92e-30 | 1.19e-28*** |
| Reticulocyte percentage | 0.020 | 1.69e-3 | 11.737 | 8.34e-32 | 5.17e-30*** |
| White blood cell leukocyte count | 0.027 | 1.67e-3 | 15.994 | 1.48e-57 | 9.20e-56*** |
| ***Liver*** | | | | | |
| Albumin | -0.006 | 1.73e-3 | -3.646 | 2.66e-4 | 0.017* |
| Alkaline phosphatase | 0.011 | 1.65e-3 | 6.346 | 2.21e-10 | 1.37e-8*** |
| Alanine aminotransferase | 0.011 | 1.61e-3 | 6.619 | 3.62e-11 | 2.25e-9*** |
| Aspartate aminotransferase | 0.009 | 1.64e-3 | 5.464 | 4.67e-8 | 2.89e-6*** |
| Direct bilirubin | -0.006 | 1.78e-3 | -3.599 | 3.19e-4 | 0.020* |
| Gamma glutamyltransferase | 0.014 | 1.64e-3 | 8.411 | 4.09e-17 | 2.54e-15*** |
| Total bilirubin | -0.012 | 1.64e-3 | -7.226 | 4.99e-13 | 3.09e-11*** |
| ***Renal*** | | | | | |
| Creatinine | -0.008 | 1.37e-3 | -5.747 | 9.09e-9 | 5.64e-7*** |
| Cystatin C | 0.010 | 1.55e-3 | 6.281 | 3.38e-10 | 2.09e-8*** |
| Phosphate | 7.46e-4 | 1.70e-3 | 0.438 | 0.661 | 1.000 |
| Total protein | 1.24e-3 | 1.76e-3 | 0.705 | 0.481 | 1.000 |
| Urate | 2.35e-4 | 1.43e-3 | 0.164 | 0.870 | 1.000 |
| Urea | -4.04e-3 | 1.62e-3 | -2.496 | 0.013 | 0.778 |
| ***Cardiovascular*** | | | | | |
| Apolipoprotein A | -0.009 | 1.62e-3 | -5.370 | 7.89e-8 | 4.89e-6*** |
| Apolipoprotein B | -7.36e-4 | 1.68e-3 | -0.438 | 0.662 | 1.000 |
| Cholesterol | -4.10e-3 | 1.65e-3 | -2.487 | 0.013 | 0.799 |
| LDL direct | -4.10e-3 | 1.67e-3 | -2.448 | 0.014 | 0.890 |
| Lipoprotein A | 1.40e-3 | 1.89e-3 | 0.743 | 0.458 | 1.000 |
| ***Other Non-BBB-Permeable*** | | | | | |
| Calcium | 8.18e-4 | 1.75e-3 | 0.467 | 0.641 | 1.000 |
| Glycated hemoglobin | 0.011 | 1.64e-3 | 6.772 | 1.27e-11 | 7.89e-10*** |
| Rheumatoid factor | -0.007 | 1.70e-3 | -3.899 | 9.67e-5 | 0.006** |

**Table S12.** Peripheral marker minimally adjusted associations with polygenic risk scores (PRS) at threshold ≤ 0.05 for major depressive disorder. BBB, blood-brain barrier; CRP, C-reactive protein; HDL, high-density lipoprotein; IGF-1, insulin-like growth factor 1; LDL, low-density lipoprotein; NLR, neutrophil-to-lymphocyte ratio; p-uncorr., p-uncorrected value; p-corr., Bonferroni p-corrected value; S.E., standard error; SHBG, sex hormone binding globulin. ***p corr. ≤ 0.001, **p corr. ≤ 0.01, *p corr. ≤ 0.05

| **Biomarker** | **Effect Size (β)** | **S.E.** | **t statistic** | **p-uncorr.** | **p-corr.** |
| --- | --- | --- | --- | --- | --- |
| ***BBB-Permeable*** | | | | | |
| Estradiol | 4.95e-5 | 1.58e-3 | 0.031 | 0.975 | 1.000 |
| Glucose | 0.009 | 1.75e-3 | 5.047 | 4.50e-7 | 2.79e-5*** |
| HDL cholesterol | -0.013 | 1.60e-3 | -8.432 | 3.40e-17 | 2.11e-15*** |
| IGF-1 | -0.014 | 1.63e-3 | -8.624 | 6.50e-18 | 4.03e-16*** |
| SHBG | -0.009 | 1.62e-3 | -5.374 | 7.69e-8 | 4.77e-6*** |
| Testosterone | -3.03e-3 | 7.77e-4 | -3.899 | 9.65e-5 | 0.006** |
| Triglycerides | 0.022 | 1.65e-3 | 13.068 | 5.11e-39 | 3.17e-37*** |
| Vitamin D | -0.018 | 1.72e-3 | -10.558 | 4.68e-26 | 2.90e-24*** |
| ***Inflammatory and Hematological*** | | | | | |
| Basophil count | 4.88e-3 | 1.65e-3 | 2.957 | 3.11e-3 | 0.193 |
| Basophil percentage | -2.46e-3 | 1.68e-3 | -1.466 | 0.143 | 1.000 |
| CRP | 0.020 | 1.69e-3 | 11.770 | 5.68e-32 | 3.52e-30*** |
| Eosinophil count | 0.008 | 1.67e-3 | 4.696 | 2.65e-6 | 1.64e-4*** |
| Eosinophil percentage | -1.50e-3 | 1.67e-3 | -0.898 | 0.369 | 1.000 |
| Hematocrit percentage | 7.14e-4 | 1.37e-3 | 0.523 | 0.601 | 1.000 |
| Hemoglobin concentration | 1.53e-3 | 1.33e-3 | 1.148 | 0.251 | 1.000 |
| High light scatter reticulocyte count | 0.022 | 1.68e-3 | 12.995 | 1.34e-38 | 8.29e-37*** |
| High light scatter reticulocyte percentage | 0.021 | 1.69e-3 | 12.683 | 7.51e-37 | 4.66e-35*** |
| Immature reticulocyte fraction | 0.018 | 1.68e-3 | 10.973 | 5.19e-28 | 3.22e-26*** |
| Lymphocyte count | 0.019 | 1.66e-3 | 11.244 | 2.50e-29 | 1.55e-27*** |
| Lymphocyte percentage | -3.77e-3 | 1.66e-3 | -2.273 | 0.023 | 1.000 |
| Mean corpuscular hemoglobin | -1.04e-3 | 1.66e-3 | -0.628 | 0.530 | 1.000 |
| Mean corpuscular hemoglobin concentration | 2.02e-3 | 1.66e-3 | 1.220 | 0.222 | 1.000 |
| Mean corpuscular volume | -1.97e-3 | 1.66e-3 | -1.187 | 0.235 | 1.000 |
| Mean platelet thrombocyte volume | 0.005 | 1.67e-3 | 3.222 | 1.27e-3 | 0.079 |
| Mean reticulocyte volume | 3.51e-4 | 1.68e-3 | 0.209 | 0.834 | 1.000 |
| Mean sphered cell volume | -2.27e-3 | 1.68e-3 | -1.352 | 0.176 | 1.000 |
| Monocyte count | 0.013 | 1.62e-3 | 8.127 | 4.41e-16 | 2.73e-14*** |
| Monocyte percentage | -0.006 | 1.62e-3 | -3.670 | 2.42e-4 | 0.015* |
| Neutrophil count | 0.022 | 1.67e-3 | 13.380 | 8.07e-41 | 5.01e-39*** |
| Neutrophil percentage | 4.97e-3 | 1.67e-3 | 2.967 | 3.01e-3 | 0.187 |
| NLR | 3.78e-3 | 1.67e-3 | 2.269 | 0.023 | 1.000 |
| Nucleated red blood cell count | -1.01e-3 | 1.68e-3 | -0.599 | 0.549 | 1.000 |
| Nucleated red blood cell percentage | 1.58e-4 | 1.68e-3 | 0.094 | 0.925 | 1.000 |
| Platelet count | 3.42e-3 | 1.61e-3 | 2.119 | 0.034 | 1.000 |
| Platelet crit | 0.007 | 1.58e-3 | 4.167 | 3.08e-5 | 1.91e-3** |
| Platelet distribution width | 0.006 | 1.66e-3 | 3.685 | 2.28e-4 | 0.014* |
| Red blood cell erythrocyte count | 1.84e-3 | 1.43e-3 | 1.282 | 0.200 | 1.000 |
| Red blood cell erythrocyte distribution width | 0.005 | 1.67e-3 | 3.199 | 1.38e-3 | 0.085 |
| Reticulocyte count | 0.019 | 1.67e-3 | 11.130 | 9.09e-29 | 5.63e-27*** |
| Reticulocyte percentage | 0.019 | 1.69e-3 | 11.232 | 2.89e-29 | 1.79e-27*** |
| White blood cell leukocyte count | 0.026 | 1.67e-3 | 15.727 | 1.04e-55 | 6.44e-54*** |
| ***Liver*** | | | | | |
| Albumin | -4.65e-3 | 1.73e-3 | -2.689 | 0.007 | 0.444 |
| Alkaline phosphatase | 0.010 | 1.66e-3 | 6.034 | 1.60e-9 | 9.93e-8*** |
| Alanine aminotransferase | 0.010 | 1.61e-3 | 6.323 | 2.56e-10 | 1.59e-8*** |
| Aspartate aminotransferase | 0.009 | 1.64e-3 | 5.292 | 1.21e-7 | 7.49e-6*** |
| Direct bilirubin | -0.005 | 1.78e-3 | -2.973 | 2.95e-3 | 0.183 |
| Gamma glutamyltransferase | 0.013 | 1.64e-3 | 7.994 | 1.31e-15 | 8.11e-14*** |
| Total bilirubin | -0.012 | 1.64e-3 | -7.148 | 8.85e-13 | 5.49e-11*** |
| ***Renal*** | | | | | |
| Creatinine | -0.007 | 1.37e-3 | -5.252 | 1.50e-7 | 9.31e-6*** |
| Cystatin C | 0.013 | 1.55e-3 | 8.724 | 2.70e-18 | 1.67e-16*** |
| Phosphate | 1.82e-3 | 1.70e-3 | 1.071 | 0.284 | 1.000 |
| Total protein | 2.11e-3 | 1.76e-3 | 1.201 | 0.230 | 1.000 |
| Urate | 2.52e-3 | 1.43e-3 | 1.763 | 0.078 | 1.000 |
| Urea | -4.23e-3 | 1.62e-3 | -2.611 | 0.009 | 0.560 |
| ***Cardiovascular*** | | | | | |
| Apolipoprotein A | -0.010 | 1.62e-3 | -6.313 | 2.74e-10 | 1.70e-8*** |
| Apolipoprotein B | -1.77e-3 | 1.68e-3 | -1.050 | 0.294 | 1.000 |
| Cholesterol | -0.006 | 1.65e-3 | -3.569 | 3.58e-4 | 0.022* |
| LDL direct | -0.006 | 1.67e-3 | -3.332 | 8.62e-4 | 0.053 |
| Lipoprotein A | 1.06e-3 | 1.89e-3 | 0.563 | 0.574 | 1.000 |
| ***Other Non-BBB-Permeable*** | | | | | |
| Calcium | 2.03e-3 | 1.75e-3 | 1.158 | 0.247 | 1.000 |
| Glycated hemoglobin | 0.015 | 1.64e-3 | 8.965 | 3.12e-19 | 1.93e-17*** |
| Rheumatoid factor | -4.00e-3 | 1.70e-3 | -2.355 | 0.019 | 1.000 |

**Table S13.** Peripheral marker minimally adjusted associations with polygenic risk scores (PRS) at threshold ≤ 0.1 for major depressive disorder. BBB, blood-brain barrier; CRP, C-reactive protein; HDL, high-density lipoprotein; IGF-1, insulin-like growth factor 1; LDL, low-density lipoprotein; NLR, neutrophil-to-lymphocyte ratio; p-uncorr., p-uncorrected value; p-corr., Bonferroni p-corrected value; S.E., standard error; SHBG, sex hormone binding globulin. ***p corr. ≤ 0.001, **p corr. ≤ 0.01, *p corr. ≤ 0.05

| **Biomarker** | **Effect Size (β)** | **S.E.** | **t statistic** | **p-uncorr.** | **p-corr.** |
| --- | --- | --- | --- | --- | --- |
| ***BBB-Permeable*** | | | | | |
| Estradiol | 1.98e-4 | 1.58e-3 | 0.125 | 0.900 | 1.000 |
| Glucose | 0.008 | 1.75e-3 | 4.629 | 3.68e-6 | 2.28e-4*** |
| HDL cholesterol | -0.017 | 1.60e-3 | -10.321 | 5.71e-25 | 3.54e-23*** |
| IGF-1 | -0.015 | 1.63e-3 | -9.368 | 7.40e-21 | 4.59e-19*** |
| SHBG | -0.010 | 1.62e-3 | -5.928 | 3.07e-9 | 1.90e-7*** |
| Testosterone | -2.76e-3 | 7.77e-4 | -3.557 | 3.75e-4 | 0.023* |
| Triglycerides | 0.023 | 1.65e-3 | 14.171 | 1.43e-45 | 8.88e-44*** |
| Vitamin D | -0.019 | 1.72e-3 | -10.910 | 1.05e-27 | 6.50e-26*** |
| ***Inflammatory and Hematological*** | | | | | |
| Basophil count | 0.006 | 1.65e-3 | 3.815 | 1.36e-4 | 0.008** |
| Basophil percentage | -1.28e-3 | 1.68e-3 | -0.764 | 0.445 | 1.000 |
| CRP | 0.021 | 1.69e-3 | 12.597 | 2.24e-36 | 1.39e-34*** |
| Eosinophil count | 0.008 | 1.67e-3 | 4.994 | 5.93e-7 | 3.68e-5*** |
| Eosinophil percentage | -1.33e-3 | 1.67e-3 | -0.793 | 0.428 | 1.000 |
| Hematocrit percentage | 7.70e-4 | 1.37e-3 | 0.564 | 0.573 | 1.000 |
| Hemoglobin concentration | 1.54e-3 | 1.33e-3 | 1.155 | 0.248 | 1.000 |
| High light scatter reticulocyte count | 0.021 | 1.68e-3 | 12.789 | 1.92e-37 | 1.19e-35*** |
| High light scatter reticulocyte percentage | 0.021 | 1.69e-3 | 12.645 | 1.21e-36 | 7.51e-35*** |
| Immature reticulocyte fraction | 0.018 | 1.68e-3 | 10.608 | 2.75e-26 | 1.70e-24*** |
| Lymphocyte count | 0.020 | 1.67e-3 | 11.878 | 1.56e-32 | 9.70e-31*** |
| Lymphocyte percentage | -3.67e-3 | 1.66e-3 | -2.214 | 0.027 | 1.000 |
| Mean corpuscular hemoglobin | -2.01e-4 | 1.66e-3 | -0.121 | 0.903 | 1.000 |
| Mean corpuscular hemoglobin concentration | 1.62e-3 | 1.66e-3 | 0.974 | 0.330 | 1.000 |
| Mean corpuscular volume | -9.38e-4 | 1.66e-3 | -0.564 | 0.573 | 1.000 |
| Mean platelet thrombocyte volume | 4.30e-3 | 1.67e-3 | 2.568 | 0.010 | 0.634 |
| Mean reticulocyte volume | 1.26e-3 | 1.68e-3 | 0.753 | 0.452 | 1.000 |
| Mean sphered cell volume | -9.64e-4 | 1.68e-3 | -0.574 | 0.566 | 1.000 |
| Monocyte count | 0.014 | 1.62e-3 | 8.469 | 2.49e-17 | 1.54e-15*** |
| Monocyte percentage | -0.006 | 1.62e-3 | -3.805 | 1.42e-4 | 0.009** |
| Neutrophil count | 0.023 | 1.67e-3 | 13.930 | 4.30e-44 | 2.67e-42*** |
| Neutrophil percentage | 4.91e-3 | 1.68e-3 | 2.930 | 3.39e-3 | 0.210 |
| NLR | 3.77e-3 | 1.67e-3 | 2.263 | 0.024 | 1.000 |
| Nucleated red blood cell count | 1.23e-4 | 1.68e-3 | 0.073 | 0.942 | 1.000 |
| Nucleated red blood cell percentage | 8.56e-4 | 1.68e-3 | 0.509 | 0.611 | 1.000 |
| Platelet count | 0.005 | 1.62e-3 | 3.223 | 1.27e-3 | 0.079 |
| Platelet crit | 0.008 | 1.59e-3 | 5.033 | 4.83e-7 | 2.99e-5*** |
| Platelet distribution width | 0.006 | 1.66e-3 | 3.559 | 3.72e-4 | 0.023* |
| Red blood cell erythrocyte count | 1.46e-3 | 1.44e-3 | 1.019 | 0.308 | 1.000 |
| Red blood cell erythrocyte distribution width | 0.006 | 1.67e-3 | 3.645 | 2.67e-4 | 0.017* |
| Reticulocyte count | 0.019 | 1.67e-3 | 11.116 | 1.06e-28 | 6.54e-27*** |
| Reticulocyte percentage | 0.019 | 1.69e-3 | 11.287 | 1.55e-29 | 9.58e-28*** |
| White blood cell leukocyte count | 0.028 | 1.67e-3 | 16.470 | 6.37e-61 | 3.95e-59*** |
| ***Liver*** | | | | | |
| Albumin | -0.007 | 1.73e-3 | -3.765 | 1.67e-4 | 0.010* |
| Alkaline phosphatase | 0.012 | 1.66e-3 | 7.337 | 2.20e-13 | 1.36e-11*** |
| Alanine aminotransferase | 0.012 | 1.61e-3 | 7.153 | 8.50e-13 | 5.27e-11*** |
| Aspartate aminotransferase | 0.009 | 1.64e-3 | 5.528 | 3.25e-8 | 2.01e-6*** |
| Direct bilirubin | -0.007 | 1.78e-3 | -3.917 | 8.98e-5 | 0.006** |
| Gamma glutamyltransferase | 0.016 | 1.64e-3 | 9.550 | 1.30e-21 | 8.08e-20*** |
| Total bilirubin | -0.014 | 1.64e-3 | -8.419 | 3.82e-17 | 2.37e-15*** |
| ***Renal*** | | | | | |
| Creatinine | -0.007 | 1.37e-3 | -5.360 | 8.34e-8 | 5.17e-6*** |
| Cystatin C | 0.014 | 1.55e-3 | 9.123 | 7.33e-20 | 4.55e-18*** |
| Phosphate | 1.02e-3 | 1.70e-3 | 0.597 | 0.550 | 1.000 |
| Total protein | 8.87e-4 | 1.76e-3 | 0.504 | 0.614 | 1.000 |
| Urate | 4.78e-3 | 1.43e-3 | 3.335 | 8.54e-4 | 0.053 |
| Urea | -0.006 | 1.62e-3 | -3.503 | 4.60e-4 | 0.029* |
| ***Cardiovascular*** | | | | | |
| Apolipoprotein A | -0.013 | 1.62e-3 | -8.022 | 1.04e-15 | 6.48e-14*** |
| Apolipoprotein B | 1.81e-4 | 1.68e-3 | 0.107 | 0.914 | 1.000 |
| Cholesterol | -0.005 | 1.65e-3 | -3.237 | 1.21e-3 | 0.075 |
| LDL direct | -4.27e-3 | 1.67e-3 | -2.549 | 0.011 | 0.670 |
| Lipoprotein A | 1.30e-3 | 1.89e-3 | 0.686 | 0.493 | 1.000 |
| ***Other Non-BBB-Permeable*** | | | | | |
| Calcium | 3.16e-3 | 1.75e-3 | 1.801 | 0.072 | 1.000 |
| Glycated hemoglobin | 0.016 | 1.64e-3 | 9.653 | 4.81e-22 | 2.98e-20*** |
| Rheumatoid factor | -3.17e-3 | 1.70e-3 | -1.861 | 0.063 | 1.000 |

**Table S14.** Peripheral marker minimally adjusted associations with polygenic risk scores (PRS) at threshold ≤ 1.0 for major depressive disorder. BBB, blood-brain barrier; CRP, C-reactive protein; HDL, high-density lipoprotein; IGF-1, insulin-like growth factor 1; LDL, low-density lipoprotein; NLR, neutrophil-to-lymphocyte ratio; p-uncorr., p-uncorrected value; p-corr., Bonferroni p-corrected value; S.E., standard error; SHBG, sex hormone binding globulin. ***p corr. ≤ 0.001, **p corr. ≤ 0.01, *p corr. ≤ 0.05

| **Biomarker** | **Effect Size (β)** | **S.E.** | **t statistic** | **p-uncorr.** | **p-corr.** |
| --- | --- | --- | --- | --- | --- |
| ***BBB-Permeable*** | | | | | |
| Estradiol | -5.19e-4 | 1.59e-3 | -0.327 | 0.743 | 1.000 |
| Glucose | 0.008 | 1.76e-3 | 4.839 | 1.31e-6 | 8.11e-5*** |
| HDL cholesterol | -0.016 | 1.61e-3 | -10.188 | 2.26e-24 | 1.40e-22*** |
| IGF-1 | -0.016 | 1.64e-3 | -9.981 | 1.86e-23 | 1.16e-21*** |
| SHBG | -0.009 | 1.62e-3 | -5.475 | 4.39e-8 | 2.72e-6*** |
| Testosterone | -2.94e-3 | 7.80e-4 | -3.775 | 1.60e-4 | 0.010* |
| Triglycerides | 0.023 | 1.66e-3 | 14.107 | 3.53e-45 | 2.19e-43*** |
| Vitamin D | -0.018 | 1.72e-3 | -10.732 | 7.27e-27 | 4.51e-25*** |
| ***Inflammatory and Hematological*** | | | | | |
| Basophil count | 0.007 | 1.66e-3 | 4.447 | 8.71e-6 | 5.40e-4*** |
| Basophil percentage | -6.24e-5 | 1.68e-3 | -0.037 | 0.970 | 1.000 |
| CRP | 0.022 | 1.70e-3 | 12.898 | 4.73e-38 | 2.93e-36*** |
| Eosinophil count | 0.007 | 1.68e-3 | 3.882 | 1.03e-4 | 0.006** |
| Eosinophil percentage | -3.29e-3 | 1.68e-3 | -1.964 | 0.050 | 1.000 |
| Hematocrit percentage | 8.69e-4 | 1.37e-3 | 0.634 | 0.526 | 1.000 |
| Hemoglobin concentration | 1.23e-3 | 1.34e-3 | 0.920 | 0.357 | 1.000 |
| High light scatter reticulocyte count | 0.020 | 1.68e-3 | 12.075 | 1.45e-33 | 9.00e-32*** |
| High light scatter reticulocyte percentage | 0.020 | 1.69e-3 | 12.065 | 1.64e-33 | 1.02e-31*** |
| Immature reticulocyte fraction | 0.018 | 1.68e-3 | 10.402 | 2.45e-25 | 1.52e-23*** |
| Lymphocyte count | 0.018 | 1.67e-3 | 11.017 | 3.19e-28 | 1.98e-26*** |
| Lymphocyte percentage | -0.006 | 1.66e-3 | -3.398 | 6.78e-4 | 0.042* |
| Mean corpuscular hemoglobin | 1.23e-3 | 1.66e-3 | 0.741 | 0.459 | 1.000 |
| Mean corpuscular hemoglobin concentration | 8.97e-4 | 1.66e-3 | 0.539 | 0.590 | 1.000 |
| Mean corpuscular volume | 1.17e-3 | 1.67e-3 | 0.700 | 0.484 | 1.000 |
| Mean platelet thrombocyte volume | 3.27e-3 | 1.68e-3 | 1.946 | 0.052 | 1.000 |
| Mean reticulocyte volume | 3.93e-3 | 1.68e-3 | 2.335 | 0.020 | 1.000 |
| Mean sphered cell volume | 1.22e-3 | 1.69e-3 | 0.726 | 0.468 | 1.000 |
| Monocyte count | 0.013 | 1.63e-3 | 7.755 | 8.89e-15 | 5.51e-13*** |
| Monocyte percentage | -0.008 | 1.63e-3 | -4.659 | 3.18e-6 | 1.97e-4*** |
| Neutrophil count | 0.024 | 1.68e-3 | 14.488 | 1.49e-47 | 9.22e-46*** |
| Neutrophil percentage | 0.007 | 1.68e-3 | 4.299 | 1.71e-5 | 1.06e-3** |
| NLR | 0.006 | 1.67e-3 | 3.677 | 2.36e-4 | 0.015* |
| Nucleated red blood cell count | 1.27e-3 | 1.69e-3 | 0.753 | 0.451 | 1.000 |
| Nucleated red blood cell percentage | 1.65e-3 | 1.69e-3 | 0.978 | 0.328 | 1.000 |
| Platelet count | 0.006 | 1.62e-3 | 3.446 | 5.70e-4 | 0.035* |
| Platelet crit | 0.008 | 1.59e-3 | 5.033 | 4.84e-7 | 3.00e-5*** |
| Platelet distribution width | 4.57e-3 | 1.66e-3 | 2.744 | 0.006 | 0.376 |
| Red blood cell erythrocyte count | 1.66e-4 | 1.44e-3 | 0.115 | 0.908 | 1.000 |
| Red blood cell erythrocyte distribution width | 0.006 | 1.68e-3 | 3.848 | 1.19e-4 | 0.007** |
| Reticulocyte count | 0.017 | 1.67e-3 | 10.371 | 3.40e-25 | 2.11e-23*** |
| Reticulocyte percentage | 0.018 | 1.69e-3 | 10.710 | 9.25e-27 | 5.74e-25*** |
| White blood cell leukocyte count | 0.028 | 1.68e-3 | 16.430 | 1.23e-60 | 7.64e-59*** |
| ***Liver*** | | | | | |
| Albumin | -0.006 | 1.74e-3 | -3.709 | 2.08e-4 | 0.013* |
| Alkaline phosphatase | 0.010 | 1.66e-3 | 6.301 | 2.97e-10 | 1.84e-8*** |
| Alanine aminotransferase | 0.011 | 1.62e-3 | 7.004 | 2.50e-12 | 1.55e-10*** |
| Aspartate aminotransferase | 0.008 | 1.65e-3 | 5.089 | 3.59e-7 | 2.23e-5*** |
| Direct bilirubin | -0.006 | 1.79e-3 | -3.589 | 3.32e-4 | 0.021* |
| Gamma glutamyltransferase | 0.016 | 1.65e-3 | 9.790 | 1.25e-22 | 7.72e-21*** |
| Total bilirubin | -0.014 | 1.65e-3 | -8.227 | 1.92e-16 | 1.19e-14*** |
| ***Renal*** | | | | | |
| Creatinine | -0.007 | 1.37e-3 | -5.447 | 5.12e-8 | 3.18e-6*** |
| Cystatin C | 0.015 | 1.55e-3 | 9.954 | 2.43e-23 | 1.51e-21*** |
| Phosphate | -4.81e-4 | 1.71e-3 | -0.282 | 0.778 | 1.000 |
| Total protein | -2.90e-4 | 1.77e-3 | -0.164 | 0.869 | 1.000 |
| Urate | 0.008 | 1.44e-3 | 5.615 | 1.97e-8 | 1.22e-6*** |
| Urea | -0.005 | 1.62e-3 | -3.353 | 8.00e-4 | 0.050* |
| ***Cardiovascular*** | | | | | |
| Apolipoprotein A | -0.013 | 1.63e-3 | -7.764 | 8.27e-15 | 5.13e-13*** |
| Apolipoprotein B | 2.96e-4 | 1.69e-3 | 0.175 | 0.861 | 1.000 |
| Cholesterol | -0.006 | 1.66e-3 | -3.394 | 6.88e-4 | 0.043* |
| LDL direct | -4.34e-3 | 1.68e-3 | -2.581 | 0.010 | 0.611 |
| Lipoprotein A | 8.61e-5 | 1.90e-3 | 0.045 | 0.964 | 1.000 |
| ***Other Non-BBB-Permeable*** | | | | | |
| Calcium | 3.79e-3 | 1.76e-3 | 2.151 | 0.031 | 1.000 |
| Glycated hemoglobin | 0.015 | 1.64e-3 | 9.442 | 3.67e-21 | 2.27e-19*** |
| Rheumatoid factor | -3.40e-3 | 1.71e-3 | -1.991 | 0.046 | 1.000 |

**Table S15.** Peripheral marker fully adjusted associations with polygenic risk scores (PRS) at threshold ≤ 0.01 for major depressive disorder. BBB, blood-brain barrier; CRP, C-reactive protein; HDL, high-density lipoprotein; IGF-1, insulin-like growth factor 1; LDL, low-density lipoprotein; NLR, neutrophil-to-lymphocyte ratio; p-uncorr., p-uncorrected value; p-corr., Bonferroni p-corrected value; S.E., standard error; SHBG, sex hormone binding globulin. ***p corr. ≤ 0.001, **p corr. ≤ 0.01, *p corr. ≤ 0.05

| **Biomarker** | **Effect Size (β)** | **S.E.** | **t statistic** | **p-uncorr.** | **p-corr.** |
| --- | --- | --- | --- | --- | --- |
| ***BBB-Permeable*** | | | | | |
| Estradiol | 7.80e-5 | 1.59e-3 | 0.049 | 0.961 | 1.000 |
| Glucose | 2.64e-3 | 1.73e-3 | 1.526 | 0.127 | 1.000 |
| HDL cholesterol | -6.36e-5 | 1.49e-3 | -0.043 | 0.966 | 1.000 |
| IGF-1 | -4.84e-3 | 1.62e-3 | -2.997 | 2.73e-3 | 0.169 |
| SHBG | 1.98e-3 | 1.50e-3 | 1.319 | 0.187 | 1.000 |
| Testosterone | -6.11e-4 | 7.65e-4 | -0.799 | 0.424 | 1.000 |
| Triglycerides | 0.010 | 1.58e-3 | 6.095 | 1.10e-9 | 6.80e-8*** |
| Vitamin D | -0.007 | 1.69e-3 | -4.212 | 2.54e-5 | 1.57e-3** |
| ***Inflammatory and Hematological*** | | | | | |
| Basophil count | 1.17e-3 | 1.64e-3 | 0.710 | 0.478 | 1.000 |
| Basophil percentage | -2.13e-3 | 1.68e-3 | -1.269 | 0.205 | 1.000 |
| CRP | 3.93e-3 | 1.58e-3 | 2.483 | 0.013 | 0.808 |
| Eosinophil count | 4.67e-3 | 1.66e-3 | 2.809 | 4.97e-3 | 0.308 |
| Eosinophil percentage | 8.11e-5 | 1.67e-3 | 0.048 | 0.961 | 1.000 |
| Hematocrit percentage | -2.69e-3 | 1.36e-3 | -1.982 | 0.048 | 1.000 |
| Hemoglobin concentration | -1.06e-3 | 1.32e-3 | -0.806 | 0.421 | 1.000 |
| High light scatter reticulocyte count | 0.008 | 1.53e-3 | 5.132 | 2.86e-7 | 1.78e-5*** |
| High light scatter reticulocyte percentage | 0.008 | 1.55e-3 | 4.951 | 7.39e-7 | 4.58e-5*** |
| Immature reticulocyte fraction | 0.006 | 1.60e-3 | 4.002 | 6.28e-5 | 3.90e-3** |
| Lymphocyte count | 0.011 | 1.61e-3 | 6.907 | 4.97e-12 | 3.08e-10*** |
| Lymphocyte percentage | -8.22e-4 | 1.66e-3 | -0.495 | 0.620 | 1.000 |
| Mean corpuscular hemoglobin | 1.50e-3 | 1.63e-3 | 0.920 | 0.358 | 1.000 |
| Mean corpuscular hemoglobin concentration | 4.70e-3 | 1.66e-3 | 2.827 | 4.69e-3 | 0.291 |
| Mean corpuscular volume | -5.39e-4 | 1.63e-3 | -0.331 | 0.741 | 1.000 |
| Mean platelet thrombocyte volume | 1.48e-3 | 1.68e-3 | 0.881 | 0.378 | 1.000 |
| Mean reticulocyte volume | -4.08e-3 | 1.67e-3 | -2.443 | 0.015 | 0.902 |
| Mean sphered cell volume | -4.24e-3 | 1.64e-3 | -2.580 | 0.010 | 0.612 |
| Monocyte count | 0.007 | 1.60e-3 | 4.548 | 5.42e-6 | 3.36e-4*** |
| Monocyte percentage | -3.08e-3 | 1.62e-3 | -1.902 | 0.057 | 1.000 |
| Neutrophil count | 0.012 | 1.62e-3 | 7.207 | 5.71e-13 | 3.54e-11*** |
| Neutrophil percentage | 1.60e-3 | 1.68e-3 | 0.953 | 0.340 | 1.000 |
| NLR | 7.03e-4 | 1.67e-3 | 0.421 | 0.674 | 1.000 |
| Nucleated red blood cell count | -9.77e-4 | 1.68e-3 | -0.580 | 0.562 | 1.000 |
| Nucleated red blood cell percentage | -1.45e-4 | 1.68e-3 | -0.086 | 0.931 | 1.000 |
| Platelet count | 1.56e-3 | 1.61e-3 | 0.965 | 0.335 | 1.000 |
| Platelet crit | 2.55e-3 | 1.58e-3 | 1.616 | 0.106 | 1.000 |
| Platelet distribution width | 0.007 | 1.66e-3 | 4.258 | 2.07e-5 | 1.28e-3** |
| Red blood cell erythrocyte count | -2.12e-3 | 1.41e-3 | -1.502 | 0.133 | 1.000 |
| Red blood cell erythrocyte distribution width | -4.63e-3 | 1.66e-3 | -2.788 | 0.005 | 0.329 |
| Reticulocyte count | 0.007 | 1.55e-3 | 4.680 | 2.87e-6 | 1.78e-4*** |
| Reticulocyte percentage | 0.008 | 1.59e-3 | 5.112 | 3.19e-7 | 1.98e-5*** |
| White blood cell leukocyte count | 0.014 | 1.59e-3 | 8.862 | 7.91e-19 | 4.91e-17*** |
| ***Liver*** | | | | | |
| Albumin | -2.23e-4 | 1.71e-3 | -0.130 | 0.897 | 1.000 |
| Alkaline phosphatase | 3.42e-3 | 1.63e-3 | 2.102 | 0.036 | 1.000 |
| Alanine aminotransferase | 3.24e-3 | 1.55e-3 | 2.084 | 0.037 | 1.000 |
| Aspartate aminotransferase | 0.007 | 1.64e-3 | 4.158 | 3.21e-5 | 1.99e-3** |
| Direct bilirubin | -2.76e-3 | 1.78e-3 | -1.554 | 0.120 | 1.000 |
| Gamma glutamyltransferase | 0.006 | 1.61e-3 | 3.827 | 1.30e-4 | 0.008** |
| Total bilirubin | -0.006 | 1.63e-3 | -3.619 | 2.96e-4 | 0.018* |
| ***Renal*** | | | | | |
| Creatinine | -0.008 | 1.36e-3 | -5.919 | 3.25e-9 | 2.01e-7*** |
| Cystatin C | -2.70e-3 | 1.45e-3 | -1.861 | 0.063 | 1.000 |
| Phosphate | 8.95e-4 | 1.70e-3 | 0.527 | 0.598 | 1.000 |
| Total protein | 2.76e-3 | 1.76e-3 | 1.571 | 0.116 | 1.000 |
| Urate | -0.010 | 1.33e-3 | -7.171 | 7.48e-13 | 4.63e-11*** |
| Urea | -4.34e-3 | 1.61e-3 | -2.692 | 0.007 | 0.441 |
| ***Cardiovascular*** | | | | | |
| Apolipoprotein A | 7.67e-4 | 1.55e-3 | 0.493 | 0.622 | 1.000 |
| Apolipoprotein B | -3.07e-3 | 1.68e-3 | -1.827 | 0.068 | 1.000 |
| Cholesterol | -1.85e-3 | 1.65e-3 | -1.118 | 0.263 | 1.000 |
| LDL direct | -3.78e-3 | 1.68e-3 | -2.255 | 0.024 | 1.000 |
| Lipoprotein A | 9.44e-4 | 1.90e-3 | 0.498 | 0.619 | 1.000 |
| ***Other Non-BBB-Permeable*** | | | | | |
| Calcium | 1.91e-3 | 1.76e-3 | 1.088 | 0.276 | 1.000 |
| Glycated hemoglobin | 9.01e-4 | 1.58e-3 | 0.570 | 0.568 | 1.000 |
| Rheumatoid factor | -0.006 | 1.71e-3 | -3.810 | 1.39e-4 | 0.009** |

**Table S16.** Peripheral marker fully adjusted associations with polygenic risk scores (PRS) at threshold ≤ 0.05 for major depressive disorder. BBB, blood-brain barrier; CRP, C-reactive protein; HDL, high-density lipoprotein; IGF-1, insulin-like growth factor 1; LDL, low-density lipoprotein; NLR, neutrophil-to-lymphocyte ratio; p-uncorr., p-uncorrected value; p-corr., Bonferroni p-corrected value; S.E., standard error; SHBG, sex hormone binding globulin. ***p corr. ≤ 0.001, **p corr. ≤ 0.01, *p corr. ≤ 0.05

| **Biomarker** | **Effect Size (β)** | **S.E.** | **t statistic** | **p-uncorr.** | **p-corr.** |
| --- | --- | --- | --- | --- | --- |
| ***BBB-Permeable*** | | | | | |
| Estradiol | 4.96e-4 | 1.59e-3 | 0.313 | 0.755 | 1.000 |
| Glucose | 3.26e-3 | 1.73e-3 | 1.882 | 0.060 | 1.000 |
| HDL cholesterol | 2.43e-5 | 1.49e-3 | 0.016 | 0.987 | 1.000 |
| IGF-1 | -0.008 | 1.62e-3 | -4.660 | 3.16e-6 | 1.96e-4*** |
| SHBG | 1.93e-3 | 1.50e-3 | 1.285 | 0.199 | 1.000 |
| Testosterone | -4.33e-4 | 7.66e-4 | -0.566 | 0.572 | 1.000 |
| Triglycerides | 0.009 | 1.58e-3 | 5.814 | 6.12e-9 | 3.79e-7*** |
| Vitamin D | -0.010 | 1.69e-3 | -5.952 | 2.65e-9 | 1.64e-7*** |
| ***Inflammatory and Hematological*** | | | | | |
| Basophil count | 2.16e-4 | 1.65e-3 | 0.131 | 0.896 | 1.000 |
| Basophil percentage | -3.33e-3 | 1.68e-3 | -1.981 | 0.048 | 1.000 |
| CRP | 4.88e-3 | 1.59e-3 | 3.080 | 2.07e-3 | 0.128 |
| Eosinophil count | 1.92e-3 | 1.66e-3 | 1.157 | 0.247 | 1.000 |
| Eosinophil percentage | -2.51e-3 | 1.68e-3 | -1.497 | 0.134 | 1.000 |
| Hematocrit percentage | -2.72e-3 | 1.36e-3 | -2.006 | 0.045 | 1.000 |
| Hemoglobin concentration | -1.79e-3 | 1.32e-3 | -1.353 | 0.176 | 1.000 |
| High light scatter reticulocyte count | 0.006 | 1.53e-3 | 3.896 | 9.77e-5 | 0.006** |
| High light scatter reticulocyte percentage | 0.006 | 1.56e-3 | 3.795 | 1.48e-4 | 0.009** |
| Immature reticulocyte fraction | 0.005 | 1.60e-3 | 3.368 | 7.56e-4 | 0.047* |
| Lymphocyte count | 0.007 | 1.62e-3 | 4.376 | 1.21e-5 | 7.49e-4*** |
| Lymphocyte percentage | -3.49e-3 | 1.66e-3 | -2.099 | 0.036 | 1.000 |
| Mean corpuscular hemoglobin | -6.18e-4 | 1.63e-3 | -0.379 | 0.705 | 1.000 |
| Mean corpuscular hemoglobin concentration | 2.26e-3 | 1.66e-3 | 1.356 | 0.175 | 1.000 |
| Mean corpuscular volume | -1.54e-3 | 1.63e-3 | -0.945 | 0.345 | 1.000 |
| Mean platelet thrombocyte volume | 4.06e-3 | 1.68e-3 | 2.418 | 0.016 | 0.967 |
| Mean reticulocyte volume | -6.65e-4 | 1.67e-3 | -0.398 | 0.691 | 1.000 |
| Mean sphered cell volume | -1.65e-3 | 1.64e-3 | -1.002 | 0.316 | 1.000 |
| Monocyte count | 0.005 | 1.60e-3 | 3.350 | 8.07e-4 | 0.050* |
| Monocyte percentage | -3.21e-3 | 1.62e-3 | -1.977 | 0.048 | 1.000 |
| Neutrophil count | 0.011 | 1.62e-3 | 6.622 | 3.54e-11 | 2.20e-9*** |
| Neutrophil percentage | 4.21e-3 | 1.68e-3 | 2.511 | 0.012 | 0.747 |
| NLR | 3.54e-3 | 1.67e-3 | 2.118 | 0.034 | 1.000 |
| Nucleated red blood cell count | -4.32e-4 | 1.69e-3 | -0.256 | 0.798 | 1.000 |
| Nucleated red blood cell percentage | 3.96e-4 | 1.68e-3 | 0.235 | 0.814 | 1.000 |
| Platelet count | 1.71e-4 | 1.62e-3 | 0.106 | 0.916 | 1.000 |
| Platelet crit | 2.22e-3 | 1.58e-3 | 1.402 | 0.161 | 1.000 |
| Platelet distribution width | 0.005 | 1.66e-3 | 3.163 | 1.56e-3 | 0.097 |
| Red blood cell erythrocyte count | -1.62e-3 | 1.41e-3 | -1.149 | 0.251 | 1.000 |
| Red blood cell erythrocyte distribution width | -1.46e-4 | 1.66e-3 | -0.088 | 0.930 | 1.000 |
| Reticulocyte count | 4.97e-3 | 1.55e-3 | 3.203 | 1.36e-3 | 0.084 |
| Reticulocyte percentage | 0.006 | 1.59e-3 | 3.545 | 3.92e-4 | 0.024* |
| White blood cell leukocyte count | 0.012 | 1.59e-3 | 7.343 | 2.09e-13 | 1.30e-11*** |
| ***Liver*** | | | | | |
| Albumin | 2.28e-3 | 1.71e-3 | 1.331 | 0.183 | 1.000 |
| Alkaline phosphatase | 1.91e-3 | 1.63e-3 | 1.174 | 0.240 | 1.000 |
| Alanine aminotransferase | 1.69e-3 | 1.56e-3 | 1.090 | 0.276 | 1.000 |
| Aspartate aminotransferase | 0.006 | 1.64e-3 | 3.770 | 1.63e-4 | 0.010* |
| Direct bilirubin | -1.28e-3 | 1.78e-3 | -0.718 | 0.473 | 1.000 |
| Gamma glutamyltransferase | 4.37e-3 | 1.61e-3 | 2.717 | 0.007 | 0.409 |
| Total bilirubin | -0.005 | 1.63e-3 | -3.154 | 1.61e-3 | 0.100 |
| ***Renal*** | | | | | |
| Creatinine | -0.008 | 1.36e-3 | -5.514 | 3.51e-8 | 2.18e-6*** |
| Cystatin C | -6.01e-4 | 1.45e-3 | -0.414 | 0.679 | 1.000 |
| Phosphate | 2.09e-3 | 1.70e-3 | 1.228 | 0.220 | 1.000 |
| Total protein | 3.79e-3 | 1.76e-3 | 2.153 | 0.031 | 1.000 |
| Urate | -0.009 | 1.33e-3 | -6.425 | 1.32e-10 | 8.16e-9*** |
| Urea | -4.55e-3 | 1.61e-3 | -2.823 | 4.76e-3 | 0.295 |
| ***Cardiovascular*** | | | | | |
| Apolipoprotein A | 4.41e-4 | 1.56e-3 | 0.284 | 0.777 | 1.000 |
| Apolipoprotein B | -4.69e-3 | 1.68e-3 | -2.790 | 0.005 | 0.327 |
| Cholesterol | -3.59e-3 | 1.65e-3 | -2.177 | 0.030 | 1.000 |
| LDL direct | -0.006 | 1.68e-3 | -3.282 | 1.03e-3 | 0.064 |
| Lipoprotein A | 5.74e-4 | 1.90e-3 | 0.303 | 0.762 | 1.000 |
| ***Other Non-BBB-Permeable*** | | | | | |
| Calcium | 3.28e-3 | 1.76e-3 | 1.864 | 0.062 | 1.000 |
| Glycated hemoglobin | 2.77e-3 | 1.58e-3 | 1.754 | 0.079 | 1.000 |
| Rheumatoid factor | -3.93e-3 | 1.71e-3 | -2.305 | 0.021 | 1.000 |

**Table S17.** Peripheral marker fully adjusted associations with polygenic risk scores (PRS) at threshold ≤ 0.1 for major depressive disorder. BBB, blood-brain barrier; CRP, C-reactive protein; HDL, high-density lipoprotein; IGF-1, insulin-like growth factor 1; LDL, low-density lipoprotein; NLR, neutrophil-to-lymphocyte ratio; p-uncorr., p-uncorrected value; p-corr., Bonferroni p-corrected value; S.E., standard error; SHBG, sex hormone binding globulin. ***p corr. ≤ 0.001, **p corr. ≤ 0.01, *p corr. ≤ 0.05

| **Biomarker** | **Effect Size (β)** | **S.E.** | **t statistic** | **p-uncorr.** | **p-corr.** |
| --- | --- | --- | --- | --- | --- |
| ***BBB-Permeable*** | | | | | |
| Estradiol | 6.45e-4 | 1.59e-3 | 0.407 | 0.684 | 1.000 |
| Glucose | 2.35e-3 | 1.73e-3 | 1.356 | 0.175 | 1.000 |
| HDL cholesterol | -2.50e-3 | 1.49e-3 | -1.677 | 0.093 | 1.000 |
| IGF-1 | -0.009 | 1.62e-3 | -5.313 | 1.08e-7 | 6.70e-6*** |
| SHBG | 1.35e-3 | 1.51e-3 | 0.898 | 0.369 | 1.000 |
| Testosterone | -8.05e-5 | 7.66e-4 | -0.105 | 0.916 | 1.000 |
| Triglycerides | 0.011 | 1.58e-3 | 6.694 | 2.18e-11 | 1.35e-9*** |
| Vitamin D | -0.010 | 1.69e-3 | -6.086 | 1.16e-9 | 7.18e-8*** |
| ***Inflammatory and Hematological*** | | | | | |
| Basophil count | 1.36e-3 | 1.65e-3 | 0.827 | 0.408 | 1.000 |
| Basophil percentage | -2.25e-3 | 1.68e-3 | -1.340 | 0.180 | 1.000 |
| CRP | 0.006 | 1.59e-3 | 3.550 | 3.85e-4 | 0.024* |
| Eosinophil count | 2.04e-3 | 1.67e-3 | 1.226 | 0.220 | 1.000 |
| Eosinophil percentage | -2.50e-3 | 1.68e-3 | -1.492 | 0.136 | 1.000 |
| Hematocrit percentage | -2.86e-3 | 1.36e-3 | -2.105 | 0.035 | 1.000 |
| Hemoglobin concentration | -1.96e-3 | 1.32e-3 | -1.480 | 0.139 | 1.000 |
| High light scatter reticulocyte count | 0.005 | 1.53e-3 | 3.391 | 6.97e-4 | 0.043* |
| High light scatter reticulocyte percentage | 0.005 | 1.56e-3 | 3.478 | 5.06e-4 | 0.031* |
| Immature reticulocyte fraction | 4.38e-3 | 1.60e-3 | 2.737 | 0.006 | 0.384 |
| Lymphocyte count | 0.008 | 1.62e-3 | 4.648 | 3.36e-6 | 2.08e-4*** |
| Lymphocyte percentage | -3.43e-3 | 1.66e-3 | -2.061 | 0.039 | 1.000 |
| Mean corpuscular hemoglobin | 8.56e-5 | 1.63e-3 | 0.052 | 0.958 | 1.000 |
| Mean corpuscular hemoglobin concentration | 1.87e-3 | 1.66e-3 | 1.122 | 0.262 | 1.000 |
| Mean corpuscular volume | -6.70e-4 | 1.63e-3 | -0.411 | 0.681 | 1.000 |
| Mean platelet thrombocyte volume | 2.89e-3 | 1.68e-3 | 1.721 | 0.085 | 1.000 |
| Mean reticulocyte volume | 1.15e-4 | 1.67e-3 | 0.069 | 0.945 | 1.000 |
| Mean sphered cell volume | -5.30e-4 | 1.65e-3 | -0.322 | 0.748 | 1.000 |
| Monocyte count | 0.006 | 1.61e-3 | 3.463 | 5.35e-4 | 0.033* |
| Monocyte percentage | -3.30e-3 | 1.62e-3 | -2.033 | 0.042 | 1.000 |
| Neutrophil count | 0.011 | 1.62e-3 | 6.877 | 6.11e-12 | 3.79e-10*** |
| Neutrophil percentage | 4.18e-3 | 1.68e-3 | 2.488 | 0.013 | 0.796 |
| NLR | 3.54e-3 | 1.67e-3 | 2.119 | 0.034 | 1.000 |
| Nucleated red blood cell count | 7.58e-4 | 1.69e-3 | 0.449 | 0.653 | 1.000 |
| Nucleated red blood cell percentage | 1.14e-3 | 1.69e-3 | 0.676 | 0.499 | 1.000 |
| Platelet count | 1.78e-3 | 1.62e-3 | 1.103 | 0.270 | 1.000 |
| Platelet crit | 3.37e-3 | 1.58e-3 | 2.129 | 0.033 | 1.000 |
| Platelet distribution width | 0.005 | 1.66e-3 | 3.082 | 2.06e-3 | 0.128 |
| Red blood cell erythrocyte count | -2.09e-3 | 1.42e-3 | -1.479 | 0.139 | 1.000 |
| Red blood cell erythrocyte distribution width | 3.99e-4 | 1.66e-3 | 0.240 | 0.811 | 1.000 |
| Reticulocyte count | 4.59e-3 | 1.55e-3 | 2.953 | 3.14e-3 | 0.195 |
| Reticulocyte percentage | 0.005 | 1.59e-3 | 3.383 | 7.16e-4 | 0.044* |
| White blood cell leukocyte count | 0.012 | 1.59e-3 | 7.699 | 1.37e-14 | 8.52e-13*** |
| ***Liver*** | | | | | |
| Albumin | 6.44e-4 | 1.71e-3 | 0.375 | 0.707 | 1.000 |
| Alkaline phosphatase | 3.70e-3 | 1.63e-3 | 2.269 | 0.023 | 1.000 |
| Alanine aminotransferase | 2.74e-3 | 1.56e-3 | 1.761 | 0.078 | 1.000 |
| Aspartate aminotransferase | 0.007 | 1.64e-3 | 3.985 | 6.76e-5 | 4.19e-3** |
| Direct bilirubin | -2.76e-3 | 1.78e-3 | -1.552 | 0.121 | 1.000 |
| Gamma glutamyltransferase | 0.007 | 1.61e-3 | 4.086 | 4.38e-5 | 2.72e-3** |
| Total bilirubin | -0.007 | 1.63e-3 | -4.213 | 2.52e-5 | 1.56e-3** |
| ***Renal*** | | | | | |
| Creatinine | -0.008 | 1.36e-3 | -5.559 | 2.71e-8 | 1.68e-6*** |
| Cystatin C | -5.41e-4 | 1.45e-3 | -0.372 | 0.710 | 1.000 |
| Phosphate | 1.23e-3 | 1.70e-3 | 0.725 | 0.469 | 1.000 |
| Total protein | 2.65e-3 | 1.76e-3 | 1.505 | 0.132 | 1.000 |
| Urate | -0.007 | 1.33e-3 | -4.921 | 8.63e-7 | 5.35e-5*** |
| Urea | -0.006 | 1.61e-3 | -3.717 | 2.02e-4 | 0.013* |
| ***Cardiovascular*** | | | | | |
| Apolipoprotein A | -1.94e-3 | 1.56e-3 | -1.245 | 0.213 | 1.000 |
| Apolipoprotein B | -2.86e-3 | 1.68e-3 | -1.700 | 0.089 | 1.000 |
| Cholesterol | -2.96e-3 | 1.65e-3 | -1.794 | 0.073 | 1.000 |
| LDL direct | -4.21e-3 | 1.68e-3 | -2.506 | 0.012 | 0.756 |
| Lipoprotein A | 9.05e-4 | 1.90e-3 | 0.476 | 0.634 | 1.000 |
| ***Other Non-BBB-Permeable*** | | | | | |
| Calcium | 4.36e-3 | 1.76e-3 | 2.477 | 0.013 | 0.820 |
| Glycated hemoglobin | 3.53e-3 | 1.58e-3 | 2.231 | 0.026 | 1.000 |
| Rheumatoid factor | -3.10e-3 | 1.71e-3 | -1.817 | 0.069 | 1.000 |

**Table S18.** Peripheral marker fully adjusted associations with polygenic risk scores (PRS) at threshold ≤ 1.0 for major depressive disorder. BBB, blood-brain barrier; CRP, C-reactive protein; HDL, high-density lipoprotein; IGF-1, insulin-like growth factor 1; LDL, low-density lipoprotein; NLR, neutrophil-to-lymphocyte ratio; p-uncorr., p-uncorrected value; p-corr., Bonferroni p-corrected value; S.E., standard error; SHBG, sex hormone binding globulin. ***p corr. ≤ 0.001, **p corr. ≤ 0.01, *p corr. ≤ 0.05

| **Biomarker** | **Effect Size (β)** | **S.E.** | **t statistic** | **p-uncorr.** | **p-corr.** |
| --- | --- | --- | --- | --- | --- |
| ***BBB-Permeable*** | | | | | |
| Estradiol | -6.78e-5 | 1.59e-3 | -0.043 | 0.966 | 1.000 |
| Glucose | 2.68e-3 | 1.74e-3 | 1.544 | 0.123 | 1.000 |
| HDL cholesterol | -2.18e-3 | 1.49e-3 | -1.457 | 0.145 | 1.000 |
| IGF-1 | -0.010 | 1.62e-3 | -5.879 | 4.14e-9 | 2.57e-7*** |
| SHBG | 2.45e-3 | 1.51e-3 | 1.621 | 0.105 | 1.000 |
| Testosterone | -1.44e-4 | 7.69e-4 | -0.187 | 0.851 | 1.000 |
| Triglycerides | 0.010 | 1.58e-3 | 6.530 | 6.57e-11 | 4.07e-9*** |
| Vitamin D | -0.010 | 1.70e-3 | -5.939 | 2.87e-9 | 1.78e-7*** |
| ***Inflammatory and Hematological*** | | | | | |
| Basophil count | 2.47e-3 | 1.65e-3 | 1.496 | 0.135 | 1.000 |
| Basophil percentage | -9.69e-4 | 1.69e-3 | -0.575 | 0.566 | 1.000 |
| CRP | 0.006 | 1.59e-3 | 3.783 | 1.55e-4 | 0.010* |
| Eosinophil count | 7.17e-5 | 1.67e-3 | 0.043 | 0.966 | 1.000 |
| Eosinophil percentage | -4.61e-3 | 1.68e-3 | -2.743 | 0.006 | 0.378 |
| Hematocrit percentage | -2.78e-3 | 1.36e-3 | -2.042 | 0.041 | 1.000 |
| Hemoglobin concentration | -2.28e-3 | 1.33e-3 | -1.715 | 0.086 | 1.000 |
| High light scatter reticulocyte count | 3.66e-3 | 1.54e-3 | 2.377 | 0.017 | 1.000 |
| High light scatter reticulocyte percentage | 4.10e-3 | 1.56e-3 | 2.627 | 0.009 | 0.535 |
| Immature reticulocyte fraction | 3.78e-3 | 1.60e-3 | 2.355 | 0.019 | 1.000 |
| Lymphocyte count | 0.006 | 1.62e-3 | 3.803 | 1.43e-4 | 0.009** |
| Lymphocyte percentage | -0.005 | 1.67e-3 | -3.278 | 1.05e-3 | 0.065 |
| Mean corpuscular hemoglobin | 1.76e-3 | 1.64e-3 | 1.074 | 0.283 | 1.000 |
| Mean corpuscular hemoglobin concentration | 1.21e-3 | 1.67e-3 | 0.725 | 0.469 | 1.000 |
| Mean corpuscular volume | 1.66e-3 | 1.64e-3 | 1.016 | 0.310 | 1.000 |
| Mean platelet thrombocyte volume | 1.84e-3 | 1.68e-3 | 1.090 | 0.276 | 1.000 |
| Mean reticulocyte volume | 2.82e-3 | 1.68e-3 | 1.678 | 0.093 | 1.000 |
| Mean sphered cell volume | 1.91e-3 | 1.65e-3 | 1.155 | 0.248 | 1.000 |
| Monocyte count | 4.39e-3 | 1.61e-3 | 2.726 | 0.006 | 0.398 |
| Monocyte percentage | -4.80e-3 | 1.63e-3 | -2.948 | 3.20e-3 | 0.198 |
| Neutrophil count | 0.012 | 1.63e-3 | 7.539 | 4.75e-14 | 2.94e-12*** |
| Neutrophil percentage | 0.007 | 1.68e-3 | 3.915 | 9.03e-5 | 0.006** |
| NLR | 0.006 | 1.68e-3 | 3.584 | 3.39e-4 | 0.021* |
| Nucleated red blood cell count | 1.94e-3 | 1.69e-3 | 1.148 | 0.251 | 1.000 |
| Nucleated red blood cell percentage | 1.97e-3 | 1.69e-3 | 1.163 | 0.245 | 1.000 |
| Platelet count | 2.18e-3 | 1.62e-3 | 1.346 | 0.178 | 1.000 |
| Platelet crit | 3.41e-3 | 1.59e-3 | 2.149 | 0.032 | 1.000 |
| Platelet distribution width | 3.68e-3 | 1.67e-3 | 2.208 | 0.027 | 1.000 |
| Red blood cell erythrocyte count | -3.53e-3 | 1.42e-3 | -2.484 | 0.013 | 0.806 |
| Red blood cell erythrocyte distribution width | 7.66e-4 | 1.67e-3 | 0.459 | 0.646 | 1.000 |
| Reticulocyte count | 3.05e-3 | 1.56e-3 | 1.957 | 0.050 | 1.000 |
| Reticulocyte percentage | 4.11e-3 | 1.59e-3 | 2.581 | 0.010 | 0.611 |
| White blood cell leukocyte count | 0.012 | 1.60e-3 | 7.751 | 9.13e-15 | 5.66e-13*** |
| ***Liver*** | | | | | |
| Albumin | 6.84e-4 | 1.72e-3 | 0.397 | 0.691 | 1.000 |
| Alkaline phosphatase | 1.93e-3 | 1.64e-3 | 1.182 | 0.237 | 1.000 |
| Alanine aminotransferase | 2.27e-3 | 1.56e-3 | 1.452 | 0.147 | 1.000 |
| Aspartate aminotransferase | 0.006 | 1.64e-3 | 3.455 | 5.51e-4 | 0.034* |
| Direct bilirubin | -2.24e-3 | 1.79e-3 | -1.257 | 0.209 | 1.000 |
| Gamma glutamyltransferase | 0.007 | 1.61e-3 | 4.205 | 2.61e-5 | 1.62e-3** |
| Total bilirubin | -0.007 | 1.64e-3 | -4.068 | 4.74e-5 | 2.94e-3** |
| ***Renal*** | | | | | |
| Creatinine | -0.008 | 1.37e-3 | -5.770 | 7.94e-9 | 4.92e-7*** |
| Cystatin C | 6.99e-4 | 1.46e-3 | 0.479 | 0.632 | 1.000 |
| Phosphate | -1.90e-4 | 1.71e-3 | -0.111 | 0.911 | 1.000 |
| Total protein | 1.37e-3 | 1.77e-3 | 0.776 | 0.438 | 1.000 |
| Urate | -3.57e-3 | 1.33e-3 | -2.674 | 0.007 | 0.465 |
| Urea | -0.006 | 1.62e-3 | -3.647 | 2.65e-4 | 0.016* |
| ***Cardiovascular*** | | | | | |
| Apolipoprotein A | -1.45e-3 | 1.56e-3 | -0.926 | 0.354 | 1.000 |
| Apolipoprotein B | -2.85e-3 | 1.69e-3 | -1.692 | 0.091 | 1.000 |
| Cholesterol | -3.30e-3 | 1.66e-3 | -1.989 | 0.047 | 1.000 |
| LDL direct | -4.35e-3 | 1.68e-3 | -2.583 | 0.010 | 0.608 |
| Lipoprotein A | -2.94e-4 | 1.91e-3 | -0.154 | 0.877 | 1.000 |
| ***Other Non-BBB-Permeable*** | | | | | |
| Calcium | 4.90e-3 | 1.76e-3 | 2.776 | 0.005 | 0.341 |
| Glycated hemoglobin | 3.17e-3 | 1.59e-3 | 1.998 | 0.046 | 1.000 |
| Rheumatoid factor | -3.37e-3 | 1.71e-3 | -1.966 | 0.049 | 1.000 |

**Table S19.** Peripheral marker minimally adjusted associations with polygenic risk scores (PRS) at threshold ≤ 0.01 for schizophrenia. BBB, blood-brain barrier; CRP, C-reactive protein; HDL, high-density lipoprotein; IGF-1, insulin-like growth factor 1; LDL, low-density lipoprotein; NLR, neutrophil-to-lymphocyte ratio; p-uncorr., p-uncorrected value; p-corr., Bonferroni p-corrected value; S.E., standard error; SHBG, sex hormone binding globulin. ***p corr. ≤ 0.001, **p corr. ≤ 0.01, *p corr. ≤ 0.05

| **Biomarker** | **Effect Size (β)** | **S.E.** | **t statistic** | **p-uncorr.** | **p-corr.** |
| --- | --- | --- | --- | --- | --- |
| ***BBB-Permeable*** | | | | | |
| Estradiol | 3.86e-3 | 1.60e-3 | 2.409 | 0.016 | 0.992 |
| Glucose | -0.005 | 1.77e-3 | -2.959 | 3.09e-3 | 0.191 |
| HDL cholesterol | 0.014 | 1.62e-3 | 8.436 | 3.31e-17 | 2.05e-15*** |
| IGF-1 | 0.008 | 1.65e-3 | 4.959 | 7.08e-7 | 4.39e-5*** |
| SHBG | 0.013 | 1.64e-3 | 7.990 | 1.36e-15 | 8.40e-14*** |
| Testosterone | 4.35e-3 | 7.87e-4 | 5.532 | 3.16e-8 | 1.96e-6*** |
| Triglycerides | -3.44e-3 | 1.67e-3 | -2.056 | 0.040 | 1.000 |
| Vitamin D | -0.010 | 1.74e-3 | -5.994 | 2.04e-9 | 1.27e-7*** |
| ***Inflammatory and Hematological*** | | | | | |
| Basophil count | 0.006 | 1.67e-3 | 3.403 | 6.67e-4 | 0.041* |
| Basophil percentage | 3.81e-3 | 1.70e-3 | 2.244 | 0.025 | 1.000 |
| CRP | -0.007 | 1.71e-3 | -4.059 | 4.93e-5 | 3.05e-3** |
| Eosinophil count | 0.015 | 1.69e-3 | 8.884 | 6.46e-19 | 4.01e-17*** |
| Eosinophil percentage | 0.010 | 1.69e-3 | 6.202 | 5.57e-10 | 3.45e-8*** |
| Hematocrit percentage | 4.09e-3 | 1.38e-3 | 2.960 | 3.08e-3 | 0.191 |
| Hemoglobin concentration | 0.005 | 1.35e-3 | 4.046 | 5.22e-5 | 3.24e-3** |
| High light scatter reticulocyte count | -0.007 | 1.70e-3 | -4.102 | 4.10e-5 | 2.54e-3** |
| High light scatter reticulocyte percentage | -0.008 | 1.71e-3 | -4.767 | 1.87e-6 | 1.16e-4*** |
| Immature reticulocyte fraction | -0.008 | 1.70e-3 | -4.953 | 7.31e-7 | 4.53e-5*** |
| Lymphocyte count | 0.022 | 1.69e-3 | 13.067 | 5.21e-39 | 3.23e-37*** |
| Lymphocyte percentage | 0.011 | 1.68e-3 | 6.781 | 1.19e-11 | 7.39e-10*** |
| Mean corpuscular hemoglobin | 3.23e-3 | 1.68e-3 | 1.926 | 0.054 | 1.000 |
| Mean corpuscular hemoglobin concentration | 4.26e-3 | 1.68e-3 | 2.538 | 0.011 | 0.692 |
| Mean corpuscular volume | 1.06e-3 | 1.68e-3 | 0.629 | 0.529 | 1.000 |
| Mean platelet thrombocyte volume | 1.42e-3 | 1.69e-3 | 0.841 | 0.400 | 1.000 |
| Mean reticulocyte volume | -4.59e-3 | 1.70e-3 | -2.706 | 0.007 | 0.422 |
| Mean sphered cell volume | 2.53e-3 | 1.70e-3 | 1.486 | 0.137 | 1.000 |
| Monocyte count | 0.010 | 1.64e-3 | 5.910 | 3.42e-9 | 2.12e-7*** |
| Monocyte percentage | -2.02e-3 | 1.64e-3 | -1.227 | 0.220 | 1.000 |
| Neutrophil count | 0.007 | 1.69e-3 | 4.167 | 3.09e-5 | 1.92e-3** |
| Neutrophil percentage | -0.012 | 1.69e-3 | -7.072 | 1.53e-12 | 9.51e-11*** |
| NLR | -0.011 | 1.69e-3 | -6.571 | 4.99e-11 | 3.10e-9*** |
| Nucleated red blood cell count | -3.61e-3 | 1.70e-3 | -2.121 | 0.034 | 1.000 |
| Nucleated red blood cell percentage | -3.55e-3 | 1.70e-3 | -2.086 | 0.037 | 1.000 |
| Platelet count | 3.37e-3 | 1.63e-3 | 2.063 | 0.039 | 1.000 |
| Platelet crit | 4.91e-3 | 1.60e-3 | 3.063 | 2.20e-3 | 0.136 |
| Platelet distribution width | 1.99e-3 | 1.68e-3 | 1.183 | 0.237 | 1.000 |
| Red blood cell erythrocyte count | 3.39e-3 | 1.45e-3 | 2.332 | 0.020 | 1.000 |
| Red blood cell erythrocyte distribution width | -0.006 | 1.69e-3 | -3.768 | 1.65e-4 | 0.010* |
| Reticulocyte count | -4.61e-3 | 1.69e-3 | -2.728 | 0.006 | 0.395 |
| Reticulocyte percentage | -0.006 | 1.71e-3 | -3.347 | 8.17e-4 | 0.051 |
| White blood cell leukocyte count | 0.016 | 1.69e-3 | 9.386 | 6.26e-21 | 3.88e-19*** |
| ***Liver*** | | | | | |
| Albumin | 2.00e-3 | 1.75e-3 | 1.141 | 0.254 | 1.000 |
| Alkaline phosphatase | -10.00e-4 | 1.68e-3 | -0.596 | 0.551 | 1.000 |
| Alanine aminotransferase | -3.91e-3 | 1.63e-3 | -2.390 | 0.017 | 1.000 |
| Aspartate aminotransferase | 0.012 | 1.67e-3 | 7.008 | 2.43e-12 | 1.51e-10*** |
| Direct bilirubin | -3.67e-4 | 1.81e-3 | -0.203 | 0.839 | 1.000 |
| Gamma glutamyltransferase | -1.39e-3 | 1.66e-3 | -0.838 | 0.402 | 1.000 |
| Total bilirubin | 6.59e-4 | 1.66e-3 | 0.396 | 0.692 | 1.000 |
| ***Renal*** | | | | | |
| Creatinine | -0.012 | 1.39e-3 | -8.998 | 2.31e-19 | 1.43e-17*** |
| Cystatin C | -0.017 | 1.57e-3 | -10.752 | 5.89e-27 | 3.65e-25*** |
| Phosphate | 0.009 | 1.72e-3 | 5.218 | 1.81e-7 | 1.12e-5*** |
| Total protein | 0.015 | 1.78e-3 | 8.206 | 2.29e-16 | 1.42e-14*** |
| Urate | -0.015 | 1.45e-3 | -10.660 | 1.57e-26 | 9.76e-25*** |
| Urea | -0.006 | 1.64e-3 | -3.378 | 7.29e-4 | 0.045* |
| ***Cardiovascular*** | | | | | |
| Apolipoprotein A | 0.009 | 1.65e-3 | 5.708 | 1.15e-8 | 7.10e-7*** |
| Apolipoprotein B | 2.43e-3 | 1.70e-3 | 1.428 | 0.153 | 1.000 |
| Cholesterol | 0.007 | 1.67e-3 | 4.248 | 2.16e-5 | 1.34e-3** |
| LDL direct | 3.11e-3 | 1.70e-3 | 1.832 | 0.067 | 1.000 |
| Lipoprotein A | 3.50e-3 | 1.92e-3 | 1.829 | 0.067 | 1.000 |
| ***Other Non-BBB-Permeable*** | | | | | |
| Calcium | -0.006 | 1.78e-3 | -3.362 | 7.74e-4 | 0.048* |
| Glycated hemoglobin | -0.007 | 1.66e-3 | -4.427 | 9.56e-6 | 5.93e-4*** |
| Rheumatoid factor | -0.014 | 1.72e-3 | -8.030 | 9.78e-16 | 6.06e-14*** |

**Table S20.** Peripheral marker minimally adjusted associations with polygenic risk scores (PRS) at threshold ≤ 0.05 for schizophrenia. BBB, blood-brain barrier; CRP, C-reactive protein; HDL, high-density lipoprotein; IGF-1, insulin-like growth factor 1; LDL, low-density lipoprotein; NLR, neutrophil-to-lymphocyte ratio; p-uncorr., p-uncorrected value; p-corr., Bonferroni p-corrected value; S.E., standard error; SHBG, sex hormone binding globulin. ***p corr. ≤ 0.001, **p corr. ≤ 0.01, *p corr. ≤ 0.05

| **Biomarker** | **Effect Size (β)** | **S.E.** | **t statistic** | **p-uncorr.** | **p-corr.** |
| --- | --- | --- | --- | --- | --- |
| ***BBB-Permeable*** | | | | | |
| Estradiol | 4.00e-3 | 1.62e-3 | 2.465 | 0.014 | 0.851 |
| Glucose | -0.008 | 1.79e-3 | -4.456 | 8.34e-6 | 5.17e-4*** |
| HDL cholesterol | 0.013 | 1.64e-3 | 7.951 | 1.86e-15 | 1.15e-13*** |
| IGF-1 | 0.006 | 1.67e-3 | 3.667 | 2.46e-4 | 0.015* |
| SHBG | 0.014 | 1.66e-3 | 8.590 | 8.72e-18 | 5.40e-16*** |
| Testosterone | 4.40e-3 | 7.97e-4 | 5.521 | 3.37e-8 | 2.09e-6*** |
| Triglycerides | -4.37e-3 | 1.69e-3 | -2.583 | 0.010 | 0.607 |
| Vitamin D | -0.013 | 1.76e-3 | -7.558 | 4.11e-14 | 2.55e-12*** |
| ***Inflammatory and Hematological*** | | | | | |
| Basophil count | 0.007 | 1.69e-3 | 3.844 | 1.21e-4 | 0.008** |
| Basophil percentage | 0.005 | 1.72e-3 | 2.915 | 3.56e-3 | 0.221 |
| CRP | -0.008 | 1.73e-3 | -4.485 | 7.29e-6 | 4.52e-4*** |
| Eosinophil count | 0.012 | 1.71e-3 | 7.110 | 1.16e-12 | 7.22e-11*** |
| Eosinophil percentage | 0.007 | 1.71e-3 | 4.138 | 3.51e-5 | 2.18e-3** |
| Hematocrit percentage | 2.69e-3 | 1.40e-3 | 1.918 | 0.055 | 1.000 |
| Hemoglobin concentration | 3.94e-3 | 1.36e-3 | 2.883 | 3.94e-3 | 0.244 |
| High light scatter reticulocyte count | -0.009 | 1.72e-3 | -5.279 | 1.30e-7 | 8.07e-6*** |
| High light scatter reticulocyte percentage | -0.010 | 1.73e-3 | -5.595 | 2.21e-8 | 1.37e-6*** |
| Immature reticulocyte fraction | -0.009 | 1.72e-3 | -4.957 | 7.17e-7 | 4.45e-5*** |
| Lymphocyte count | 0.019 | 1.71e-3 | 11.234 | 2.80e-29 | 1.73e-27*** |
| Lymphocyte percentage | 0.009 | 1.70e-3 | 5.374 | 7.72e-8 | 4.79e-6*** |
| Mean corpuscular hemoglobin | 0.005 | 1.70e-3 | 3.107 | 1.89e-3 | 0.117 |
| Mean corpuscular hemoglobin concentration | 3.26e-3 | 1.70e-3 | 1.920 | 0.055 | 1.000 |
| Mean corpuscular volume | 3.84e-3 | 1.70e-3 | 2.251 | 0.024 | 1.000 |
| Mean platelet thrombocyte volume | 3.54e-5 | 1.72e-3 | 0.021 | 0.984 | 1.000 |
| Mean reticulocyte volume | 6.54e-5 | 1.72e-3 | 0.038 | 0.970 | 1.000 |
| Mean sphered cell volume | 0.006 | 1.72e-3 | 3.462 | 5.36e-4 | 0.033* |
| Monocyte count | 0.008 | 1.66e-3 | 5.013 | 5.37e-7 | 3.33e-5*** |
| Monocyte percentage | -2.25e-3 | 1.66e-3 | -1.351 | 0.177 | 1.000 |
| Neutrophil count | 0.007 | 1.71e-3 | 4.330 | 1.49e-5 | 9.23e-4*** |
| Neutrophil percentage | -0.009 | 1.72e-3 | -5.342 | 9.22e-8 | 5.72e-6*** |
| NLR | -0.009 | 1.71e-3 | -5.033 | 4.84e-7 | 3.00e-5*** |
| Nucleated red blood cell count | -1.86e-3 | 1.72e-3 | -1.081 | 0.280 | 1.000 |
| Nucleated red blood cell percentage | -7.83e-4 | 1.72e-3 | -0.454 | 0.650 | 1.000 |
| Platelet count | 3.84e-3 | 1.66e-3 | 2.323 | 0.020 | 1.000 |
| Platelet crit | 4.84e-3 | 1.62e-3 | 2.980 | 2.88e-3 | 0.179 |
| Platelet distribution width | 1.12e-3 | 1.70e-3 | 0.660 | 0.509 | 1.000 |
| Red blood cell erythrocyte count | 5.27e-4 | 1.47e-3 | 0.358 | 0.720 | 1.000 |
| Red blood cell erythrocyte distribution width | -0.006 | 1.71e-3 | -3.275 | 1.06e-3 | 0.065 |
| Reticulocyte count | -0.007 | 1.71e-3 | -4.351 | 1.36e-5 | 8.42e-4*** |
| Reticulocyte percentage | -0.008 | 1.73e-3 | -4.526 | 6.01e-6 | 3.73e-4*** |
| White blood cell leukocyte count | 0.015 | 1.71e-3 | 8.544 | 1.31e-17 | 8.10e-16*** |
| ***Liver*** | | | | | |
| Albumin | 2.10e-3 | 1.78e-3 | 1.185 | 0.236 | 1.000 |
| Alkaline phosphatase | -2.50e-4 | 1.70e-3 | -0.147 | 0.883 | 1.000 |
| Alanine aminotransferase | -4.05e-3 | 1.66e-3 | -2.447 | 0.014 | 0.894 |
| Aspartate aminotransferase | 0.011 | 1.69e-3 | 6.263 | 3.79e-10 | 2.35e-8*** |
| Direct bilirubin | 1.91e-3 | 1.83e-3 | 1.044 | 0.296 | 1.000 |
| Gamma glutamyltransferase | -2.56e-3 | 1.68e-3 | -1.519 | 0.129 | 1.000 |
| Total bilirubin | 2.41e-3 | 1.68e-3 | 1.433 | 0.152 | 1.000 |
| ***Renal*** | | | | | |
| Creatinine | -0.014 | 1.41e-3 | -9.873 | 5.51e-23 | 3.42e-21*** |
| Cystatin C | -0.013 | 1.59e-3 | -8.507 | 1.78e-17 | 1.11e-15*** |
| Phosphate | 0.010 | 1.75e-3 | 5.766 | 8.14e-9 | 5.05e-7*** |
| Total protein | 0.012 | 1.80e-3 | 6.691 | 2.21e-11 | 1.37e-9*** |
| Urate | -0.014 | 1.47e-3 | -9.848 | 7.02e-23 | 4.35e-21*** |
| Urea | -0.008 | 1.66e-3 | -4.572 | 4.83e-6 | 2.99e-4*** |
| ***Cardiovascular*** | | | | | |
| Apolipoprotein A | 0.009 | 1.67e-3 | 5.176 | 2.27e-7 | 1.41e-5*** |
| Apolipoprotein B | 2.77e-3 | 1.73e-3 | 1.605 | 0.109 | 1.000 |
| Cholesterol | 0.006 | 1.69e-3 | 3.767 | 1.65e-4 | 0.010* |
| LDL direct | 2.92e-3 | 1.72e-3 | 1.700 | 0.089 | 1.000 |
| Lipoprotein A | 0.006 | 1.94e-3 | 2.900 | 3.73e-3 | 0.231 |
| ***Other Non-BBB-Permeable*** | | | | | |
| Calcium | -0.005 | 1.80e-3 | -2.995 | 2.75e-3 | 0.170 |
| Glycated hemoglobin | -0.008 | 1.68e-3 | -4.883 | 1.04e-6 | 6.47e-5*** |
| Rheumatoid factor | -0.010 | 1.74e-3 | -5.636 | 1.74e-8 | 1.08e-6*** |

**Table S21.** Peripheral marker minimally adjusted associations with polygenic risk scores (PRS) at threshold ≤ 0.1 for schizophrenia. BBB, blood-brain barrier; CRP, C-reactive protein; HDL, high-density lipoprotein; IGF-1, insulin-like growth factor 1; LDL, low-density lipoprotein; NLR, neutrophil-to-lymphocyte ratio; p-uncorr., p-uncorrected value; p-corr., Bonferroni p-corrected value; S.E., standard error; SHBG, sex hormone binding globulin. ***p corr. ≤ 0.001, **p corr. ≤ 0.01, *p corr. ≤ 0.05

| **Biomarker** | **Effect Size (β)** | **S.E.** | **t statistic** | **p-uncorr.** | **p-corr.** |
| --- | --- | --- | --- | --- | --- |
| ***BBB-Permeable*** | | | | | |
| Estradiol | 4.57e-3 | 1.63e-3 | 2.800 | 0.005 | 0.317 |
| Glucose | -0.007 | 1.80e-3 | -3.990 | 6.62e-5 | 4.10e-3** |
| HDL cholesterol | 0.013 | 1.65e-3 | 7.864 | 3.73e-15 | 2.31e-13*** |
| IGF-1 | 0.006 | 1.68e-3 | 3.832 | 1.27e-4 | 0.008** |
| SHBG | 0.015 | 1.67e-3 | 9.094 | 9.63e-20 | 5.97e-18*** |
| Testosterone | 4.72e-3 | 8.02e-4 | 5.894 | 3.78e-9 | 2.35e-7*** |
| Triglycerides | -0.005 | 1.70e-3 | -3.070 | 2.14e-3 | 0.133 |
| Vitamin D | -0.013 | 1.77e-3 | -7.406 | 1.31e-13 | 8.10e-12*** |
| ***Inflammatory and Hematological*** | | | | | |
| Basophil count | 0.007 | 1.70e-3 | 3.858 | 1.14e-4 | 0.007** |
| Basophil percentage | 0.005 | 1.73e-3 | 3.016 | 2.56e-3 | 0.159 |
| CRP | -0.007 | 1.74e-3 | -4.248 | 2.16e-5 | 1.34e-3** |
| Eosinophil count | 0.010 | 1.72e-3 | 5.877 | 4.19e-9 | 2.60e-7*** |
| Eosinophil percentage | 0.005 | 1.72e-3 | 3.069 | 2.14e-3 | 0.133 |
| Hematocrit percentage | 2.54e-3 | 1.41e-3 | 1.804 | 0.071 | 1.000 |
| Hemoglobin concentration | 3.62e-3 | 1.37e-3 | 2.638 | 0.008 | 0.517 |
| High light scatter reticulocyte count | -0.010 | 1.73e-3 | -5.878 | 4.16e-9 | 2.58e-7*** |
| High light scatter reticulocyte percentage | -0.011 | 1.74e-3 | -6.076 | 1.24e-9 | 7.66e-8*** |
| Immature reticulocyte fraction | -0.009 | 1.73e-3 | -5.243 | 1.58e-7 | 9.82e-6*** |
| Lymphocyte count | 0.017 | 1.72e-3 | 10.048 | 9.48e-24 | 5.88e-22*** |
| Lymphocyte percentage | 0.007 | 1.71e-3 | 4.281 | 1.86e-5 | 1.15e-3** |
| Mean corpuscular hemoglobin | 0.007 | 1.71e-3 | 3.805 | 1.42e-4 | 0.009** |
| Mean corpuscular hemoglobin concentration | 2.95e-3 | 1.71e-3 | 1.728 | 0.084 | 1.000 |
| Mean corpuscular volume | 0.006 | 1.71e-3 | 3.225 | 1.26e-3 | 0.078 |
| Mean platelet thrombocyte volume | 6.79e-4 | 1.73e-3 | 0.394 | 0.694 | 1.000 |
| Mean reticulocyte volume | 2.44e-3 | 1.73e-3 | 1.412 | 0.158 | 1.000 |
| Mean sphered cell volume | 0.008 | 1.73e-3 | 4.614 | 3.96e-6 | 2.45e-4*** |
| Monocyte count | 0.008 | 1.67e-3 | 4.663 | 3.12e-6 | 1.94e-4*** |
| Monocyte percentage | -2.66e-3 | 1.67e-3 | -1.589 | 0.112 | 1.000 |
| Neutrophil count | 0.008 | 1.72e-3 | 4.644 | 3.42e-6 | 2.12e-4*** |
| Neutrophil percentage | -0.007 | 1.73e-3 | -4.169 | 3.05e-5 | 1.89e-3** |
| NLR | -0.007 | 1.72e-3 | -3.822 | 1.32e-4 | 0.008** |
| Nucleated red blood cell count | -2.52e-3 | 1.73e-3 | -1.455 | 0.146 | 1.000 |
| Nucleated red blood cell percentage | -1.49e-3 | 1.73e-3 | -0.860 | 0.390 | 1.000 |
| Platelet count | 3.02e-3 | 1.66e-3 | 1.813 | 0.070 | 1.000 |
| Platelet crit | 4.26e-3 | 1.63e-3 | 2.607 | 0.009 | 0.566 |
| Platelet distribution width | 1.28e-3 | 1.71e-3 | 0.748 | 0.454 | 1.000 |
| Red blood cell erythrocyte count | -3.11e-4 | 1.48e-3 | -0.210 | 0.834 | 1.000 |
| Red blood cell erythrocyte distribution width | -0.006 | 1.72e-3 | -3.420 | 6.27e-4 | 0.039* |
| Reticulocyte count | -0.008 | 1.72e-3 | -4.877 | 1.08e-6 | 6.68e-5*** |
| Reticulocyte percentage | -0.009 | 1.74e-3 | -4.954 | 7.26e-7 | 4.50e-5*** |
| White blood cell leukocyte count | 0.014 | 1.72e-3 | 8.215 | 2.13e-16 | 1.32e-14*** |
| ***Liver*** | | | | | |
| Albumin | 2.46e-3 | 1.79e-3 | 1.375 | 0.169 | 1.000 |
| Alkaline phosphatase | 6.68e-4 | 1.71e-3 | 0.391 | 0.696 | 1.000 |
| Alanine aminotransferase | -4.04e-3 | 1.67e-3 | -2.424 | 0.015 | 0.951 |
| Aspartate aminotransferase | 0.010 | 1.70e-3 | 5.660 | 1.51e-8 | 9.38e-7*** |
| Direct bilirubin | 2.32e-3 | 1.84e-3 | 1.262 | 0.207 | 1.000 |
| Gamma glutamyltransferase | -2.39e-3 | 1.69e-3 | -1.411 | 0.158 | 1.000 |
| Total bilirubin | 2.89e-3 | 1.69e-3 | 1.707 | 0.088 | 1.000 |
| ***Renal*** | | | | | |
| Creatinine | -0.014 | 1.41e-3 | -9.682 | 3.61e-22 | 2.24e-20*** |
| Cystatin C | -0.012 | 1.60e-3 | -7.236 | 4.61e-13 | 2.86e-11*** |
| Phosphate | 0.009 | 1.76e-3 | 5.392 | 6.97e-8 | 4.32e-6*** |
| Total protein | 0.011 | 1.82e-3 | 5.811 | 6.22e-9 | 3.86e-7*** |
| Urate | -0.014 | 1.48e-3 | -9.353 | 8.55e-21 | 5.30e-19*** |
| Urea | -0.007 | 1.67e-3 | -4.066 | 4.78e-5 | 2.97e-3** |
| ***Cardiovascular*** | | | | | |
| Apolipoprotein A | 0.008 | 1.68e-3 | 4.740 | 2.13e-6 | 1.32e-4*** |
| Apolipoprotein B | 1.51e-3 | 1.74e-3 | 0.871 | 0.383 | 1.000 |
| Cholesterol | 0.005 | 1.70e-3 | 2.937 | 3.31e-3 | 0.206 |
| LDL direct | 1.67e-3 | 1.73e-3 | 0.969 | 0.333 | 1.000 |
| Lipoprotein A | 0.006 | 1.95e-3 | 3.153 | 1.62e-3 | 0.100 |
| ***Other Non-BBB-Permeable*** | | | | | |
| Calcium | -0.005 | 1.81e-3 | -2.764 | 0.006 | 0.355 |
| Glycated hemoglobin | -0.007 | 1.69e-3 | -4.109 | 3.98e-5 | 2.47e-3** |
| Rheumatoid factor | -0.007 | 1.76e-3 | -3.995 | 6.48e-5 | 4.02e-3** |

**Table S22.** Peripheral marker minimally adjusted associations with polygenic risk scores (PRS) at threshold ≤ 1.0 for schizophrenia. BBB, blood-brain barrier; CRP, C-reactive protein; HDL, high-density lipoprotein; IGF-1, insulin-like growth factor 1; LDL, low-density lipoprotein; NLR, neutrophil-to-lymphocyte ratio; p-uncorr., p-uncorrected value; p-corr., Bonferroni p-corrected value; S.E., standard error; SHBG, sex hormone binding globulin. ***p corr. ≤ 0.001, **p corr. ≤ 0.01, *p corr. ≤ 0.05

| **Biomarker** | **Effect Size (β)** | **S.E.** | **t statistic** | **p-uncorr.** | **p-corr.** |
| --- | --- | --- | --- | --- | --- |
| ***BBB-Permeable*** | | | | | |
| Estradiol | 4.36e-3 | 1.66e-3 | 2.629 | 0.009 | 0.530 |
| Glucose | -0.008 | 1.84e-3 | -4.119 | 3.81e-5 | 2.36e-3** |
| HDL cholesterol | 0.011 | 1.68e-3 | 6.682 | 2.36e-11 | 1.46e-9*** |
| IGF-1 | 4.34e-3 | 1.71e-3 | 2.541 | 0.011 | 0.685 |
| SHBG | 0.013 | 1.70e-3 | 7.906 | 2.67e-15 | 1.66e-13*** |
| Testosterone | 4.73e-3 | 8.15e-4 | 5.808 | 6.31e-9 | 3.91e-7*** |
| Triglycerides | -0.006 | 1.73e-3 | -3.443 | 5.76e-4 | 0.036* |
| Vitamin D | -0.014 | 1.80e-3 | -7.635 | 2.26e-14 | 1.40e-12*** |
| ***Inflammatory and Hematological*** | | | | | |
| Basophil count | 0.005 | 1.73e-3 | 3.080 | 2.07e-3 | 0.128 |
| Basophil percentage | 0.006 | 1.76e-3 | 3.315 | 9.16e-4 | 0.057 |
| CRP | -0.007 | 1.77e-3 | -4.206 | 2.60e-5 | 1.61e-3** |
| Eosinophil count | 0.008 | 1.75e-3 | 4.675 | 2.94e-6 | 1.82e-4*** |
| Eosinophil percentage | 0.005 | 1.75e-3 | 3.011 | 2.61e-3 | 0.162 |
| Hematocrit percentage | 6.94e-4 | 1.43e-3 | 0.485 | 0.628 | 1.000 |
| Hemoglobin concentration | 1.35e-3 | 1.39e-3 | 0.964 | 0.335 | 1.000 |
| High light scatter reticulocyte count | -0.010 | 1.76e-3 | -5.907 | 3.48e-9 | 2.16e-7*** |
| High light scatter reticulocyte percentage | -0.010 | 1.77e-3 | -5.631 | 1.79e-8 | 1.11e-6*** |
| Immature reticulocyte fraction | -0.008 | 1.76e-3 | -4.310 | 1.63e-5 | 1.01e-3** |
| Lymphocyte count | 0.015 | 1.74e-3 | 8.584 | 9.20e-18 | 5.71e-16*** |
| Lymphocyte percentage | 0.009 | 1.74e-3 | 5.220 | 1.79e-7 | 1.11e-5*** |
| Mean corpuscular hemoglobin | 0.009 | 1.74e-3 | 5.020 | 5.17e-7 | 3.20e-5*** |
| Mean corpuscular hemoglobin concentration | 1.25e-3 | 1.74e-3 | 0.718 | 0.473 | 1.000 |
| Mean corpuscular volume | 0.009 | 1.74e-3 | 5.213 | 1.86e-7 | 1.15e-5*** |
| Mean platelet thrombocyte volume | -6.80e-4 | 1.75e-3 | -0.388 | 0.698 | 1.000 |
| Mean reticulocyte volume | 0.006 | 1.76e-3 | 3.604 | 3.14e-4 | 0.019* |
| Mean sphered cell volume | 0.012 | 1.76e-3 | 6.611 | 3.81e-11 | 2.36e-9*** |
| Monocyte count | 4.20e-3 | 1.70e-3 | 2.468 | 0.014 | 0.842 |
| Monocyte percentage | -2.86e-3 | 1.70e-3 | -1.683 | 0.092 | 1.000 |
| Neutrophil count | 4.00e-3 | 1.75e-3 | 2.283 | 0.022 | 1.000 |
| Neutrophil percentage | -0.009 | 1.75e-3 | -4.928 | 8.32e-7 | 5.16e-5*** |
| NLR | -0.008 | 1.75e-3 | -4.430 | 9.44e-6 | 5.85e-4*** |
| Nucleated red blood cell count | -2.11e-3 | 1.76e-3 | -1.196 | 0.232 | 1.000 |
| Nucleated red blood cell percentage | -1.32e-3 | 1.76e-3 | -0.747 | 0.455 | 1.000 |
| Platelet count | 2.32e-3 | 1.69e-3 | 1.374 | 0.169 | 1.000 |
| Platelet crit | 2.57e-3 | 1.66e-3 | 1.550 | 0.121 | 1.000 |
| Platelet distribution width | 2.45e-3 | 1.74e-3 | 1.414 | 0.157 | 1.000 |
| Red blood cell erythrocyte count | -4.24e-3 | 1.50e-3 | -2.821 | 4.79e-3 | 0.297 |
| Red blood cell erythrocyte distribution width | -0.006 | 1.75e-3 | -3.190 | 1.42e-3 | 0.088 |
| Reticulocyte count | -0.009 | 1.75e-3 | -5.360 | 8.33e-8 | 5.17e-6*** |
| Reticulocyte percentage | -0.009 | 1.77e-3 | -4.843 | 1.28e-6 | 7.95e-5*** |
| White blood cell leukocyte count | 0.009 | 1.75e-3 | 5.397 | 6.79e-8 | 4.21e-6*** |
| ***Liver*** | | | | | |
| Albumin | 3.97e-3 | 1.82e-3 | 2.188 | 0.029 | 1.000 |
| Alkaline phosphatase | 1.27e-3 | 1.74e-3 | 0.731 | 0.465 | 1.000 |
| Alanine aminotransferase | -3.46e-3 | 1.69e-3 | -2.043 | 0.041 | 1.000 |
| Aspartate aminotransferase | 0.010 | 1.72e-3 | 5.843 | 5.12e-9 | 3.18e-7*** |
| Direct bilirubin | 2.25e-3 | 1.87e-3 | 1.204 | 0.229 | 1.000 |
| Gamma glutamyltransferase | -2.39e-3 | 1.72e-3 | -1.388 | 0.165 | 1.000 |
| Total bilirubin | 1.62e-3 | 1.72e-3 | 0.941 | 0.347 | 1.000 |
| ***Renal*** | | | | | |
| Creatinine | -0.014 | 1.44e-3 | -9.983 | 1.82e-23 | 1.13e-21*** |
| Cystatin C | -0.011 | 1.62e-3 | -6.589 | 4.42e-11 | 2.74e-9*** |
| Phosphate | 0.009 | 1.79e-3 | 4.913 | 8.95e-7 | 5.55e-5*** |
| Total protein | 0.011 | 1.85e-3 | 5.975 | 2.31e-9 | 1.43e-7*** |
| Urate | -0.013 | 1.50e-3 | -8.761 | 1.94e-18 | 1.20e-16*** |
| Urea | -0.007 | 1.70e-3 | -4.310 | 1.63e-5 | 1.01e-3** |
| ***Cardiovascular*** | | | | | |
| Apolipoprotein A | 0.007 | 1.70e-3 | 3.902 | 9.55e-5 | 0.006** |
| Apolipoprotein B | 2.01e-3 | 1.76e-3 | 1.141 | 0.254 | 1.000 |
| Cholesterol | 0.005 | 1.73e-3 | 2.956 | 3.12e-3 | 0.193 |
| LDL direct | 2.42e-3 | 1.76e-3 | 1.377 | 0.169 | 1.000 |
| Lipoprotein A | 0.005 | 1.98e-3 | 2.703 | 0.007 | 0.426 |
| ***Other Non-BBB-Permeable*** | | | | | |
| Calcium | -4.49e-3 | 1.84e-3 | -2.442 | 0.015 | 0.907 |
| Glycated hemoglobin | -0.007 | 1.72e-3 | -4.249 | 2.15e-5 | 1.33e-3** |
| Rheumatoid factor | -4.98e-3 | 1.78e-3 | -2.790 | 0.005 | 0.326 |

**Table S23.** Peripheral marker fully adjusted associations with polygenic risk scores (PRS) at threshold ≤ 0.01 for schizophrenia. BBB, blood-brain barrier; CRP, C-reactive protein; HDL, high-density lipoprotein; IGF-1, insulin-like growth factor 1; LDL, low-density lipoprotein; NLR, neutrophil-to-lymphocyte ratio; p-uncorr., p-uncorrected value; p-corr., Bonferroni p-corrected value; S.E., standard error; SHBG, sex hormone binding globulin. ***p corr. ≤ 0.001, **p corr. ≤ 0.01, *p corr. ≤ 0.05

| **Biomarker** | **Effect Size (β)** | **S.E.** | **t statistic** | **p-uncorr.** | **p-corr.** |
| --- | --- | --- | --- | --- | --- |
| ***BBB-Permeable*** | | | | | |
| Estradiol | 3.55e-3 | 1.60e-3 | 2.210 | 0.027 | 1.000 |
| Glucose | -2.09e-3 | 1.75e-3 | -1.190 | 0.234 | 1.000 |
| HDL cholesterol | 0.008 | 1.51e-3 | 5.147 | 2.64e-7 | 1.64e-5*** |
| IGF-1 | 0.007 | 1.64e-3 | 3.974 | 7.06e-5 | 4.38e-3** |
| SHBG | 0.006 | 1.52e-3 | 3.711 | 2.06e-4 | 0.013* |
| Testosterone | 2.66e-3 | 7.75e-4 | 3.434 | 5.95e-4 | 0.037* |
| Triglycerides | 1.47e-3 | 1.60e-3 | 0.923 | 0.356 | 1.000 |
| Vitamin D | -0.013 | 1.71e-3 | -7.865 | 3.69e-15 | 2.29e-13*** |
| ***Inflammatory and Hematological*** | | | | | |
| Basophil count | 0.005 | 1.66e-3 | 3.264 | 1.10e-3 | 0.068 |
| Basophil percentage | 3.21e-3 | 1.70e-3 | 1.886 | 0.059 | 1.000 |
| CRP | -1.71e-3 | 1.60e-3 | -1.063 | 0.288 | 1.000 |
| Eosinophil count | 0.016 | 1.68e-3 | 9.449 | 3.45e-21 | 2.14e-19*** |
| Eosinophil percentage | 0.011 | 1.69e-3 | 6.524 | 6.86e-11 | 4.25e-9*** |
| Hematocrit percentage | 0.006 | 1.37e-3 | 4.014 | 5.97e-5 | 3.70e-3** |
| Hemoglobin concentration | 0.007 | 1.34e-3 | 5.169 | 2.36e-7 | 1.46e-5*** |
| High light scatter reticulocyte count | 1.03e-3 | 1.55e-3 | 0.665 | 0.506 | 1.000 |
| High light scatter reticulocyte percentage | -6.81e-4 | 1.57e-3 | -0.433 | 0.665 | 1.000 |
| Immature reticulocyte fraction | -3.07e-3 | 1.62e-3 | -1.897 | 0.058 | 1.000 |
| Lymphocyte count | 0.023 | 1.63e-3 | 14.139 | 2.26e-45 | 1.40e-43*** |
| Lymphocyte percentage | 0.012 | 1.68e-3 | 6.918 | 4.57e-12 | 2.83e-10*** |
| Mean corpuscular hemoglobin | -1.16e-3 | 1.65e-3 | -0.702 | 0.483 | 1.000 |
| Mean corpuscular hemoglobin concentration | 4.41e-3 | 1.68e-3 | 2.624 | 0.009 | 0.538 |
| Mean corpuscular volume | -4.06e-3 | 1.65e-3 | -2.464 | 0.014 | 0.852 |
| Mean platelet thrombocyte volume | 1.66e-3 | 1.70e-3 | 0.975 | 0.330 | 1.000 |
| Mean reticulocyte volume | -0.007 | 1.69e-3 | -4.284 | 1.84e-5 | 1.14e-3** |
| Mean sphered cell volume | -2.82e-3 | 1.66e-3 | -1.694 | 0.090 | 1.000 |
| Monocyte count | 0.011 | 1.62e-3 | 6.749 | 1.49e-11 | 9.21e-10*** |
| Monocyte percentage | -1.60e-3 | 1.64e-3 | -0.976 | 0.329 | 1.000 |
| Neutrophil count | 0.008 | 1.64e-3 | 4.609 | 4.05e-6 | 2.51e-4*** |
| Neutrophil percentage | -0.012 | 1.70e-3 | -7.308 | 2.71e-13 | 1.68e-11*** |
| NLR | -0.012 | 1.69e-3 | -6.846 | 7.63e-12 | 4.73e-10*** |
| Nucleated red blood cell count | -3.68e-3 | 1.71e-3 | -2.159 | 0.031 | 1.000 |
| Nucleated red blood cell percentage | -3.51e-3 | 1.70e-3 | -2.061 | 0.039 | 1.000 |
| Platelet count | 3.94e-3 | 1.63e-3 | 2.410 | 0.016 | 0.990 |
| Platelet crit | 0.006 | 1.60e-3 | 3.575 | 3.50e-4 | 0.022* |
| Platelet distribution width | 2.94e-3 | 1.68e-3 | 1.752 | 0.080 | 1.000 |
| Red blood cell erythrocyte count | 0.007 | 1.43e-3 | 5.148 | 2.63e-7 | 1.63e-5*** |
| Red blood cell erythrocyte distribution width | -0.005 | 1.68e-3 | -3.112 | 1.86e-3 | 0.115 |
| Reticulocyte count | 2.89e-3 | 1.57e-3 | 1.840 | 0.066 | 1.000 |
| Reticulocyte percentage | 1.18e-3 | 1.60e-3 | 0.735 | 0.462 | 1.000 |
| White blood cell leukocyte count | 0.017 | 1.61e-3 | 10.442 | 1.61e-25 | 1.00e-23*** |
| ***Liver*** | | | | | |
| Albumin | -3.88e-4 | 1.73e-3 | -0.224 | 0.823 | 1.000 |
| Alkaline phosphatase | 1.19e-3 | 1.65e-3 | 0.724 | 0.469 | 1.000 |
| Alanine aminotransferase | 1.81e-3 | 1.57e-3 | 1.150 | 0.250 | 1.000 |
| Aspartate aminotransferase | 0.014 | 1.66e-3 | 8.631 | 6.11e-18 | 3.79e-16*** |
| Direct bilirubin | -8.64e-4 | 1.80e-3 | -0.480 | 0.631 | 1.000 |
| Gamma glutamyltransferase | 1.64e-3 | 1.63e-3 | 1.009 | 0.313 | 1.000 |
| Total bilirubin | 2.35e-4 | 1.65e-3 | 0.142 | 0.887 | 1.000 |
| ***Renal*** | | | | | |
| Creatinine | -0.010 | 1.38e-3 | -7.408 | 1.28e-13 | 7.94e-12*** |
| Cystatin C | -0.013 | 1.47e-3 | -8.641 | 5.60e-18 | 3.47e-16*** |
| Phosphate | 0.006 | 1.72e-3 | 3.757 | 1.72e-4 | 0.011* |
| Total protein | 0.016 | 1.78e-3 | 9.142 | 6.19e-20 | 3.84e-18*** |
| Urate | -0.009 | 1.34e-3 | -6.877 | 6.14e-12 | 3.81e-10*** |
| Urea | -2.75e-3 | 1.63e-3 | -1.682 | 0.093 | 1.000 |
| ***Cardiovascular*** | | | | | |
| Apolipoprotein A | 4.83e-3 | 1.58e-3 | 3.066 | 2.17e-3 | 0.135 |
| Apolipoprotein B | 4.34e-3 | 1.70e-3 | 2.550 | 0.011 | 0.669 |
| Cholesterol | 0.007 | 1.67e-3 | 4.211 | 2.55e-5 | 1.58e-3** |
| LDL direct | 4.30e-3 | 1.70e-3 | 2.534 | 0.011 | 0.700 |
| Lipoprotein A | 3.35e-3 | 1.92e-3 | 1.743 | 0.081 | 1.000 |
| ***Other Non-BBB-Permeable*** | | | | | |
| Calcium | -0.007 | 1.78e-3 | -3.748 | 1.78e-4 | 0.011* |
| Glycated hemoglobin | -3.81e-3 | 1.60e-3 | -2.380 | 0.017 | 1.000 |
| Rheumatoid factor | -0.014 | 1.73e-3 | -8.094 | 5.79e-16 | 3.59e-14*** |

**Table S24.** Peripheral marker fully adjusted associations with polygenic risk scores (PRS) at threshold ≤ 0.05 for schizophrenia. BBB, blood-brain barrier; CRP, C-reactive protein; HDL, high-density lipoprotein; IGF-1, insulin-like growth factor 1; LDL, low-density lipoprotein; NLR, neutrophil-to-lymphocyte ratio; p-uncorr., p-uncorrected value; p-corr., Bonferroni p-corrected value; S.E., standard error; SHBG, sex hormone binding globulin. ***p corr. ≤ 0.001, **p corr. ≤ 0.01, *p corr. ≤ 0.05

| **Biomarker** | **Effect Size (β)** | **S.E.** | **t statistic** | **p-uncorr.** | **p-corr.** |
| --- | --- | --- | --- | --- | --- |
| ***BBB-Permeable*** | | | | | |
| Estradiol | 3.53e-3 | 1.63e-3 | 2.172 | 0.030 | 1.000 |
| Glucose | -4.38e-3 | 1.77e-3 | -2.467 | 0.014 | 0.844 |
| HDL cholesterol | 0.006 | 1.53e-3 | 4.041 | 5.32e-5 | 3.30e-3** |
| IGF-1 | 4.32e-3 | 1.66e-3 | 2.606 | 0.009 | 0.569 |
| SHBG | 0.006 | 1.54e-3 | 3.621 | 2.94e-4 | 0.018* |
| Testosterone | 2.54e-3 | 7.85e-4 | 3.239 | 1.20e-3 | 0.074 |
| Triglycerides | 1.15e-3 | 1.62e-3 | 0.713 | 0.476 | 1.000 |
| Vitamin D | -0.017 | 1.73e-3 | -9.674 | 3.92e-22 | 2.43e-20*** |
| ***Inflammatory and Hematological*** | | | | | |
| Basophil count | 0.006 | 1.69e-3 | 3.574 | 3.51e-4 | 0.022* |
| Basophil percentage | 4.22e-3 | 1.72e-3 | 2.452 | 0.014 | 0.880 |
| CRP | -1.72e-3 | 1.63e-3 | -1.057 | 0.291 | 1.000 |
| Eosinophil count | 0.013 | 1.70e-3 | 7.582 | 3.42e-14 | 2.12e-12*** |
| Eosinophil percentage | 0.008 | 1.72e-3 | 4.426 | 9.60e-6 | 5.95e-4*** |
| Hematocrit percentage | 4.37e-3 | 1.39e-3 | 3.147 | 1.65e-3 | 0.102 |
| Hemoglobin concentration | 0.006 | 1.35e-3 | 4.170 | 3.05e-5 | 1.89e-3** |
| High light scatter reticulocyte count | -7.07e-5 | 1.57e-3 | -0.045 | 0.964 | 1.000 |
| High light scatter reticulocyte percentage | -1.36e-3 | 1.59e-3 | -0.850 | 0.395 | 1.000 |
| Immature reticulocyte fraction | -2.49e-3 | 1.64e-3 | -1.523 | 0.128 | 1.000 |
| Lymphocyte count | 0.020 | 1.66e-3 | 12.119 | 8.54e-34 | 5.29e-32*** |
| Lymphocyte percentage | 0.009 | 1.70e-3 | 5.502 | 3.77e-8 | 2.34e-6*** |
| Mean corpuscular hemoglobin | 2.66e-5 | 1.67e-3 | 0.016 | 0.987 | 1.000 |
| Mean corpuscular hemoglobin concentration | 3.35e-3 | 1.70e-3 | 1.965 | 0.049 | 1.000 |
| Mean corpuscular volume | -2.22e-3 | 1.67e-3 | -1.330 | 0.184 | 1.000 |
| Mean platelet thrombocyte volume | 2.12e-4 | 1.72e-3 | 0.123 | 0.902 | 1.000 |
| Mean reticulocyte volume | -3.07e-3 | 1.71e-3 | -1.791 | 0.073 | 1.000 |
| Mean sphered cell volume | -3.65e-4 | 1.69e-3 | -0.216 | 0.829 | 1.000 |
| Monocyte count | 0.009 | 1.64e-3 | 5.756 | 8.61e-9 | 5.34e-7*** |
| Monocyte percentage | -1.77e-3 | 1.66e-3 | -1.065 | 0.287 | 1.000 |
| Neutrophil count | 0.008 | 1.66e-3 | 4.670 | 3.02e-6 | 1.87e-4*** |
| Neutrophil percentage | -0.010 | 1.72e-3 | -5.558 | 2.72e-8 | 1.69e-6*** |
| NLR | -0.009 | 1.71e-3 | -5.333 | 9.64e-8 | 5.98e-6*** |
| Nucleated red blood cell count | -2.01e-3 | 1.73e-3 | -1.165 | 0.244 | 1.000 |
| Nucleated red blood cell percentage | -8.25e-4 | 1.73e-3 | -0.478 | 0.633 | 1.000 |
| Platelet count | 4.28e-3 | 1.65e-3 | 2.586 | 0.010 | 0.602 |
| Platelet crit | 0.005 | 1.62e-3 | 3.393 | 6.92e-4 | 0.043* |
| Platelet distribution width | 2.21e-3 | 1.70e-3 | 1.302 | 0.193 | 1.000 |
| Red blood cell erythrocyte count | 0.005 | 1.45e-3 | 3.620 | 2.94e-4 | 0.018* |
| Red blood cell erythrocyte distribution width | -4.29e-3 | 1.70e-3 | -2.521 | 0.012 | 0.726 |
| Reticulocyte count | 9.52e-4 | 1.59e-3 | 0.598 | 0.550 | 1.000 |
| Reticulocyte percentage | -1.81e-4 | 1.63e-3 | -0.111 | 0.911 | 1.000 |
| White blood cell leukocyte count | 0.015 | 1.63e-3 | 9.410 | 4.98e-21 | 3.09e-19*** |
| ***Liver*** | | | | | |
| Albumin | -6.00e-4 | 1.76e-3 | -0.342 | 0.733 | 1.000 |
| Alkaline phosphatase | 2.16e-3 | 1.67e-3 | 1.293 | 0.196 | 1.000 |
| Alanine aminotransferase | 2.38e-3 | 1.59e-3 | 1.493 | 0.135 | 1.000 |
| Aspartate aminotransferase | 0.013 | 1.68e-3 | 7.986 | 1.40e-15 | 8.65e-14*** |
| Direct bilirubin | 1.46e-3 | 1.82e-3 | 0.800 | 0.424 | 1.000 |
| Gamma glutamyltransferase | 7.80e-4 | 1.65e-3 | 0.473 | 0.636 | 1.000 |
| Total bilirubin | 2.16e-3 | 1.67e-3 | 1.291 | 0.197 | 1.000 |
| ***Renal*** | | | | | |
| Creatinine | -0.011 | 1.40e-3 | -7.918 | 2.42e-15 | 1.50e-13*** |
| Cystatin C | -0.009 | 1.49e-3 | -5.810 | 6.24e-9 | 3.87e-7*** |
| Phosphate | 0.007 | 1.74e-3 | 4.020 | 5.83e-5 | 3.62e-3** |
| Total protein | 0.014 | 1.80e-3 | 7.772 | 7.74e-15 | 4.80e-13*** |
| Urate | -0.007 | 1.36e-3 | -5.325 | 1.01e-7 | 6.26e-6*** |
| Urea | -4.41e-3 | 1.65e-3 | -2.665 | 0.008 | 0.478 |
| ***Cardiovascular*** | | | | | |
| Apolipoprotein A | 3.38e-3 | 1.60e-3 | 2.118 | 0.034 | 1.000 |
| Apolipoprotein B | 4.89e-3 | 1.72e-3 | 2.838 | 4.55e-3 | 0.282 |
| Cholesterol | 0.006 | 1.69e-3 | 3.697 | 2.18e-4 | 0.014* |
| LDL direct | 4.27e-3 | 1.72e-3 | 2.485 | 0.013 | 0.804 |
| Lipoprotein A | 0.005 | 1.95e-3 | 2.811 | 4.94e-3 | 0.306 |
| ***Other Non-BBB-Permeable*** | | | | | |
| Calcium | -0.006 | 1.80e-3 | -3.472 | 5.17e-4 | 0.032* |
| Glycated hemoglobin | -4.32e-3 | 1.62e-3 | -2.666 | 0.008 | 0.476 |
| Rheumatoid factor | -0.010 | 1.75e-3 | -5.717 | 1.08e-8 | 6.72e-7*** |

**Table S25.** Peripheral marker fully adjusted associations with polygenic risk scores (PRS) at threshold ≤ 0.1 for schizophrenia. BBB, blood-brain barrier; CRP, C-reactive protein; HDL, high-density lipoprotein; IGF-1, insulin-like growth factor 1; LDL, low-density lipoprotein; NLR, neutrophil-to-lymphocyte ratio; p-uncorr., p-uncorrected value; p-corr., Bonferroni p-corrected value; S.E., standard error; SHBG, sex hormone binding globulin. ***p corr. ≤ 0.001, **p corr. ≤ 0.01, *p corr. ≤ 0.05

| **Biomarker** | **Effect Size (β)** | **S.E.** | **t statistic** | **p-uncorr.** | **p-corr.** |
| --- | --- | --- | --- | --- | --- |
| ***BBB-Permeable*** | | | | | |
| Estradiol | 4.08e-3 | 1.63e-3 | 2.496 | 0.013 | 0.778 |
| Glucose | -3.48e-3 | 1.78e-3 | -1.949 | 0.051 | 1.000 |
| HDL cholesterol | 0.006 | 1.54e-3 | 4.015 | 5.95e-5 | 3.69e-3** |
| IGF-1 | 4.77e-3 | 1.67e-3 | 2.858 | 4.27e-3 | 0.264 |
| SHBG | 0.006 | 1.55e-3 | 4.141 | 3.45e-5 | 2.14e-3** |
| Testosterone | 2.83e-3 | 7.90e-4 | 3.583 | 3.40e-4 | 0.021* |
| Triglycerides | 2.06e-4 | 1.63e-3 | 0.127 | 0.899 | 1.000 |
| Vitamin D | -0.016 | 1.74e-3 | -9.429 | 4.17e-21 | 2.59e-19*** |
| ***Inflammatory and Hematological*** | | | | | |
| Basophil count | 0.006 | 1.70e-3 | 3.465 | 5.30e-4 | 0.033* |
| Basophil percentage | 4.34e-3 | 1.73e-3 | 2.508 | 0.012 | 0.753 |
| CRP | -1.49e-3 | 1.64e-3 | -0.909 | 0.364 | 1.000 |
| Eosinophil count | 0.011 | 1.71e-3 | 6.241 | 4.35e-10 | 2.69e-8*** |
| Eosinophil percentage | 0.006 | 1.73e-3 | 3.345 | 8.23e-4 | 0.051 |
| Hematocrit percentage | 4.19e-3 | 1.40e-3 | 2.998 | 2.72e-3 | 0.169 |
| Hemoglobin concentration | 0.005 | 1.36e-3 | 3.910 | 9.25e-5 | 0.006** |
| High light scatter reticulocyte count | -1.26e-3 | 1.58e-3 | -0.798 | 0.425 | 1.000 |
| High light scatter reticulocyte percentage | -2.38e-3 | 1.60e-3 | -1.485 | 0.138 | 1.000 |
| Immature reticulocyte fraction | -3.21e-3 | 1.65e-3 | -1.949 | 0.051 | 1.000 |
| Lymphocyte count | 0.018 | 1.67e-3 | 10.668 | 1.45e-26 | 9.02e-25*** |
| Lymphocyte percentage | 0.008 | 1.71e-3 | 4.448 | 8.67e-6 | 5.38e-4*** |
| Mean corpuscular hemoglobin | 1.03e-3 | 1.68e-3 | 0.612 | 0.541 | 1.000 |
| Mean corpuscular hemoglobin concentration | 3.12e-3 | 1.71e-3 | 1.823 | 0.068 | 1.000 |
| Mean corpuscular volume | -8.09e-4 | 1.68e-3 | -0.482 | 0.630 | 1.000 |
| Mean platelet thrombocyte volume | 8.10e-4 | 1.73e-3 | 0.468 | 0.640 | 1.000 |
| Mean reticulocyte volume | -8.33e-4 | 1.72e-3 | -0.483 | 0.629 | 1.000 |
| Mean sphered cell volume | 1.43e-3 | 1.69e-3 | 0.844 | 0.399 | 1.000 |
| Monocyte count | 0.009 | 1.65e-3 | 5.242 | 1.59e-7 | 9.86e-6*** |
| Monocyte percentage | -2.09e-3 | 1.67e-3 | -1.250 | 0.211 | 1.000 |
| Neutrophil count | 0.008 | 1.67e-3 | 4.713 | 2.44e-6 | 1.51e-4*** |
| Neutrophil percentage | -0.008 | 1.73e-3 | -4.430 | 9.42e-6 | 5.84e-4*** |
| NLR | -0.007 | 1.72e-3 | -4.189 | 2.80e-5 | 1.74e-3** |
| Nucleated red blood cell count | -2.63e-3 | 1.74e-3 | -1.514 | 0.130 | 1.000 |
| Nucleated red blood cell percentage | -1.50e-3 | 1.74e-3 | -0.861 | 0.389 | 1.000 |
| Platelet count | 3.34e-3 | 1.66e-3 | 2.007 | 0.045 | 1.000 |
| Platelet crit | 4.78e-3 | 1.63e-3 | 2.931 | 3.37e-3 | 0.209 |
| Platelet distribution width | 2.39e-3 | 1.71e-3 | 1.399 | 0.162 | 1.000 |
| Red blood cell erythrocyte count | 4.50e-3 | 1.46e-3 | 3.088 | 2.01e-3 | 0.125 |
| Red blood cell erythrocyte distribution width | -4.74e-3 | 1.71e-3 | -2.768 | 0.006 | 0.350 |
| Reticulocyte count | -4.23e-5 | 1.60e-3 | -0.026 | 0.979 | 1.000 |
| Reticulocyte percentage | -1.05e-3 | 1.63e-3 | -0.641 | 0.522 | 1.000 |
| White blood cell leukocyte count | 0.014 | 1.64e-3 | 8.741 | 2.32e-18 | 1.44e-16*** |
| ***Liver*** | | | | | |
| Albumin | -1.22e-4 | 1.77e-3 | -0.069 | 0.945 | 1.000 |
| Alkaline phosphatase | 2.89e-3 | 1.68e-3 | 1.720 | 0.085 | 1.000 |
| Alanine aminotransferase | 2.57e-3 | 1.60e-3 | 1.601 | 0.109 | 1.000 |
| Aspartate aminotransferase | 0.013 | 1.69e-3 | 7.448 | 9.51e-14 | 5.90e-12*** |
| Direct bilirubin | 2.04e-3 | 1.83e-3 | 1.109 | 0.267 | 1.000 |
| Gamma glutamyltransferase | 8.45e-4 | 1.66e-3 | 0.509 | 0.610 | 1.000 |
| Total bilirubin | 2.87e-3 | 1.68e-3 | 1.707 | 0.088 | 1.000 |
| ***Renal*** | | | | | |
| Creatinine | -0.011 | 1.41e-3 | -7.638 | 2.22e-14 | 1.38e-12*** |
| Cystatin C | -0.007 | 1.50e-3 | -4.642 | 3.45e-6 | 2.14e-4*** |
| Phosphate | 0.006 | 1.75e-3 | 3.564 | 3.66e-4 | 0.023* |
| Total protein | 0.013 | 1.81e-3 | 6.995 | 2.66e-12 | 1.65e-10*** |
| Urate | -0.007 | 1.37e-3 | -4.772 | 1.82e-6 | 1.13e-4*** |
| Urea | -3.48e-3 | 1.66e-3 | -2.094 | 0.036 | 1.000 |
| ***Cardiovascular*** | | | | | |
| Apolipoprotein A | 2.79e-3 | 1.60e-3 | 1.737 | 0.082 | 1.000 |
| Apolipoprotein B | 3.64e-3 | 1.73e-3 | 2.100 | 0.036 | 1.000 |
| Cholesterol | 4.95e-3 | 1.70e-3 | 2.908 | 3.64e-3 | 0.226 |
| LDL direct | 3.09e-3 | 1.73e-3 | 1.787 | 0.074 | 1.000 |
| Lipoprotein A | 0.006 | 1.96e-3 | 3.073 | 2.12e-3 | 0.132 |
| ***Other Non-BBB-Permeable*** | | | | | |
| Calcium | -0.006 | 1.81e-3 | -3.215 | 1.30e-3 | 0.081 |
| Glycated hemoglobin | -3.25e-3 | 1.63e-3 | -1.996 | 0.046 | 1.000 |
| Rheumatoid factor | -0.007 | 1.76e-3 | -4.087 | 4.36e-5 | 2.70e-3** |

**Table S26.** Peripheral marker fully adjusted associations with polygenic risk scores (PRS) at threshold ≤ 1.0 for schizophrenia. BBB, blood-brain barrier; CRP, C-reactive protein; HDL, high-density lipoprotein; IGF-1, insulin-like growth factor 1; LDL, low-density lipoprotein; NLR, neutrophil-to-lymphocyte ratio; p-uncorr., p-uncorrected value; p-corr., Bonferroni p-corrected value; S.E., standard error; SHBG, sex hormone binding globulin. ***p corr. ≤ 0.001, **p corr. ≤ 0.01, *p corr. ≤ 0.05

| **Biomarker** | **Effect Size (β)** | **S.E.** | **t statistic** | **p-uncorr.** | **p-corr.** |
| --- | --- | --- | --- | --- | --- |
| ***BBB-Permeable*** |  |  |  |  |  |
| Estradiol | 3.91e-3 | 1.66e-3 | 2.354 | 0.019 | 1.000 |
| Glucose | -3.65e-3 | 1.81e-3 | -2.011 | 0.044 | 1.000 |
| HDL cholesterol | 4.45e-3 | 1.56e-3 | 2.848 | 4.40e-3 | 0.273 |
| IGF-1 | 2.68e-3 | 1.70e-3 | 1.579 | 0.114 | 1.000 |
| SHBG | 4.48e-3 | 1.58e-3 | 2.838 | 4.54e-3 | 0.281 |
| Testosterone | 2.77e-3 | 8.03e-4 | 3.448 | 5.66e-4 | 0.035* |
| Triglycerides | -6.70e-4 | 1.65e-3 | -0.405 | 0.686 | 1.000 |
| Vitamin D | -0.017 | 1.77e-3 | -9.522 | 1.70e-21 | 1.05e-19*** |
| ***Inflammatory and Hematological*** |  |  |  |  |  |
| Basophil count | 4.38e-3 | 1.72e-3 | 2.543 | 0.011 | 0.681 |
| Basophil percentage | 4.81e-3 | 1.76e-3 | 2.734 | 0.006 | 0.388 |
| CRP | -1.63e-3 | 1.66e-3 | -0.983 | 0.325 | 1.000 |
| Eosinophil count | 0.009 | 1.74e-3 | 4.943 | 7.69e-7 | 4.77e-5*** |
| Eosinophil percentage | 0.006 | 1.75e-3 | 3.292 | 9.94e-4 | 0.062 |
| Hematocrit percentage | 2.29e-3 | 1.42e-3 | 1.615 | 0.106 | 1.000 |
| Hemoglobin concentration | 3.00e-3 | 1.38e-3 | 2.164 | 0.030 | 1.000 |
| High light scatter reticulocyte count | -1.48e-3 | 1.60e-3 | -0.920 | 0.358 | 1.000 |
| High light scatter reticulocyte percentage | -1.80e-3 | 1.63e-3 | -1.102 | 0.271 | 1.000 |
| Immature reticulocyte fraction | -1.89e-3 | 1.67e-3 | -1.131 | 0.258 | 1.000 |
| Lymphocyte count | 0.015 | 1.69e-3 | 8.850 | 8.75e-19 | 5.43e-17*** |
| Lymphocyte percentage | 0.009 | 1.74e-3 | 5.373 | 7.74e-8 | 4.80e-6*** |
| Mean corpuscular hemoglobin | 3.03e-3 | 1.71e-3 | 1.772 | 0.076 | 1.000 |
| Mean corpuscular hemoglobin concentration | 1.40e-3 | 1.74e-3 | 0.807 | 0.420 | 1.000 |
| Mean corpuscular volume | 2.53e-3 | 1.71e-3 | 1.481 | 0.139 | 1.000 |
| Mean platelet thrombocyte volume | -6.12e-4 | 1.76e-3 | -0.348 | 0.728 | 1.000 |
| Mean reticulocyte volume | 2.85e-3 | 1.75e-3 | 1.627 | 0.104 | 1.000 |
| Mean sphered cell volume | 4.74e-3 | 1.72e-3 | 2.753 | 0.006 | 0.366 |
| Monocyte count | 4.82e-3 | 1.68e-3 | 2.865 | 4.16e-3 | 0.258 |
| Monocyte percentage | -2.13e-3 | 1.70e-3 | -1.256 | 0.209 | 1.000 |
| Neutrophil count | 3.35e-3 | 1.70e-3 | 1.974 | 0.048 | 1.000 |
| Neutrophil percentage | -0.009 | 1.76e-3 | -5.200 | 2.00e-7 | 1.24e-5*** |
| NLR | -0.008 | 1.75e-3 | -4.790 | 1.67e-6 | 1.03e-4*** |
| Nucleated red blood cell count | -2.24e-3 | 1.77e-3 | -1.269 | 0.204 | 1.000 |
| Nucleated red blood cell percentage | -1.40e-3 | 1.76e-3 | -0.791 | 0.429 | 1.000 |
| Platelet count | 2.58e-3 | 1.69e-3 | 1.524 | 0.127 | 1.000 |
| Platelet crit | 2.99e-3 | 1.66e-3 | 1.806 | 0.071 | 1.000 |
| Platelet distribution width | 3.72e-3 | 1.74e-3 | 2.138 | 0.033 | 1.000 |
| Red blood cell erythrocyte count | 6.55e-4 | 1.48e-3 | 0.443 | 0.658 | 1.000 |
| Red blood cell erythrocyte distribution width | -4.61e-3 | 1.74e-3 | -2.649 | 0.008 | 0.501 |
| Reticulocyte count | -9.53e-4 | 1.63e-3 | -0.586 | 0.558 | 1.000 |
| Reticulocyte percentage | -9.48e-4 | 1.66e-3 | -0.570 | 0.568 | 1.000 |
| White blood cell leukocyte count | 0.009 | 1.66e-3 | 5.426 | 5.77e-8 | 3.58e-6*** |
| ***Liver*** |  |  |  |  |  |
| Albumin | 1.43e-3 | 1.80e-3 | 0.794 | 0.427 | 1.000 |
| Alkaline phosphatase | 3.22e-3 | 1.71e-3 | 1.883 | 0.060 | 1.000 |
| Alanine aminotransferase | 3.33e-3 | 1.63e-3 | 2.041 | 0.041 | 1.000 |
| Aspartate aminotransferase | 0.013 | 1.72e-3 | 7.746 | 9.48e-15 | 5.88e-13*** |
| Direct bilirubin | 2.22e-3 | 1.86e-3 | 1.192 | 0.233 | 1.000 |
| Gamma glutamyltransferase | 7.77e-4 | 1.69e-3 | 0.461 | 0.645 | 1.000 |
| Total bilirubin | 1.93e-3 | 1.71e-3 | 1.131 | 0.258 | 1.000 |
| ***Renal*** |  |  |  |  |  |
| Creatinine | -0.011 | 1.43e-3 | -7.936 | 2.10e-15 | 1.30e-13*** |
| Cystatin C | -0.006 | 1.52e-3 | -4.196 | 2.72e-5 | 1.68e-3** |
| Phosphate | 0.005 | 1.78e-3 | 3.010 | 2.61e-3 | 0.162 |
| Total protein | 0.013 | 1.85e-3 | 7.248 | 4.23e-13 | 2.62e-11*** |
| Urate | -0.006 | 1.39e-3 | -4.108 | 4.00e-5 | 2.48e-3** |
| Urea | -3.88e-3 | 1.69e-3 | -2.297 | 0.022 | 1.000 |
| ***Cardiovascular*** |  |  |  |  |  |
| Apolipoprotein A | 1.47e-3 | 1.63e-3 | 0.901 | 0.367 | 1.000 |
| Apolipoprotein B | 4.08e-3 | 1.76e-3 | 2.318 | 0.020 | 1.000 |
| Cholesterol | 0.005 | 1.73e-3 | 2.909 | 3.62e-3 | 0.225 |
| LDL direct | 3.81e-3 | 1.76e-3 | 2.168 | 0.030 | 1.000 |
| Lipoprotein A | 0.005 | 1.99e-3 | 2.639 | 0.008 | 0.515 |
| ***Other Non-BBB-Permeable*** |  |  |  |  |  |
| Calcium | -0.005 | 1.84e-3 | -2.889 | 3.87e-3 | 0.240 |
| Glycated hemoglobin | -3.82e-3 | 1.66e-3 | -2.310 | 0.021 | 1.000 |
| Rheumatoid factor | -0.005 | 1.79e-3 | -2.877 | 4.01e-3 | 0.249 |

**Table S27.** Peripheral marker minimally adjusted associations with polygenic risk scores (PRS) at threshold ≤ 0.01 for bipolar disorder. BBB, blood-brain barrier; CRP, C-reactive protein; HDL, high-density lipoprotein; IGF-1, insulin-like growth factor 1; LDL, low-density lipoprotein; NLR, neutrophil-to-lymphocyte ratio; p-uncorr., p-uncorrected value; p-corr., Bonferroni p-corrected value; S.E., standard error; SHBG, sex hormone binding globulin. ***p corr. ≤ 0.001, **p corr. ≤ 0.01, *p corr. ≤ 0.05

| **Biomarker** | **Effect Size (β)** | **S.E.** | **t statistic** | **p-uncorr.** | **p-corr.** |
| --- | --- | --- | --- | --- | --- |
| ***BBB-Permeable*** | | | | | |
| Estradiol | 2.43e-3 | 1.60e-3 | 1.519 | 0.129 | 1.000 |
| Glucose | -4.93e-3 | 1.77e-3 | -2.788 | 0.005 | 0.328 |
| HDL cholesterol | 0.006 | 1.62e-3 | 3.427 | 6.09e-4 | 0.038* |
| IGF-1 | -1.46e-3 | 1.65e-3 | -0.884 | 0.377 | 1.000 |
| SHBG | 1.38e-3 | 1.64e-3 | 0.844 | 0.399 | 1.000 |
| Testosterone | 6.73e-4 | 7.86e-4 | 0.856 | 0.392 | 1.000 |
| Triglycerides | 2.08e-3 | 1.67e-3 | 1.248 | 0.212 | 1.000 |
| Vitamin D | -0.007 | 1.74e-3 | -3.796 | 1.47e-4 | 0.009** |
| ***Inflammatory and Hematological*** | | | | | |
| Basophil count | -7.25e-4 | 1.67e-3 | -0.434 | 0.664 | 1.000 |
| Basophil percentage | -3.40e-3 | 1.70e-3 | -2.006 | 0.045 | 1.000 |
| CRP | -4.46e-4 | 1.71e-3 | -0.261 | 0.794 | 1.000 |
| Eosinophil count | 1.95e-3 | 1.69e-3 | 1.150 | 0.250 | 1.000 |
| Eosinophil percentage | -2.04e-5 | 1.69e-3 | -0.012 | 0.990 | 1.000 |
| Hematocrit percentage | 1.39e-3 | 1.38e-3 | 1.006 | 0.315 | 1.000 |
| Hemoglobin concentration | 1.91e-3 | 1.35e-3 | 1.415 | 0.157 | 1.000 |
| High light scatter reticulocyte count | -0.006 | 1.70e-3 | -3.483 | 4.97e-4 | 0.031* |
| High light scatter reticulocyte percentage | -0.006 | 1.71e-3 | -3.340 | 8.38e-4 | 0.052 |
| Immature reticulocyte fraction | -4.07e-3 | 1.70e-3 | -2.397 | 0.017 | 1.000 |
| Lymphocyte count | 0.006 | 1.69e-3 | 3.794 | 1.49e-4 | 0.009** |
| Lymphocyte percentage | -7.28e-5 | 1.68e-3 | -0.043 | 0.965 | 1.000 |
| Mean corpuscular hemoglobin | 4.76e-3 | 1.68e-3 | 2.833 | 4.61e-3 | 0.286 |
| Mean corpuscular hemoglobin concentration | 2.21e-3 | 1.68e-3 | 1.316 | 0.188 | 1.000 |
| Mean corpuscular volume | 4.34e-3 | 1.68e-3 | 2.576 | 0.010 | 0.619 |
| Mean platelet thrombocyte volume | -4.12e-3 | 1.69e-3 | -2.435 | 0.015 | 0.924 |
| Mean reticulocyte volume | 6.00e-4 | 1.70e-3 | 0.353 | 0.724 | 1.000 |
| Mean sphered cell volume | 0.005 | 1.70e-3 | 3.214 | 1.31e-3 | 0.081 |
| Monocyte count | 2.18e-3 | 1.64e-3 | 1.327 | 0.184 | 1.000 |
| Monocyte percentage | -3.87e-3 | 1.64e-3 | -2.357 | 0.018 | 1.000 |
| Neutrophil count | 0.006 | 1.69e-3 | 3.822 | 1.33e-4 | 0.008** |
| Neutrophil percentage | 1.64e-3 | 1.69e-3 | 0.966 | 0.334 | 1.000 |
| NLR | 8.44e-4 | 1.69e-3 | 0.500 | 0.617 | 1.000 |
| Nucleated red blood cell count | 1.38e-3 | 1.70e-3 | 0.814 | 0.416 | 1.000 |
| Nucleated red blood cell percentage | 4.65e-4 | 1.70e-3 | 0.273 | 0.785 | 1.000 |
| Platelet count | 3.11e-3 | 1.63e-3 | 1.902 | 0.057 | 1.000 |
| Platelet crit | 1.23e-3 | 1.60e-3 | 0.770 | 0.441 | 1.000 |
| Platelet distribution width | -3.39e-3 | 1.68e-3 | -2.019 | 0.043 | 1.000 |
| Red blood cell erythrocyte count | -1.30e-3 | 1.45e-3 | -0.896 | 0.370 | 1.000 |
| Red blood cell erythrocyte distribution width | -4.34e-3 | 1.69e-3 | -2.564 | 0.010 | 0.642 |
| Reticulocyte count | -0.005 | 1.69e-3 | -3.070 | 2.14e-3 | 0.133 |
| Reticulocyte percentage | -0.005 | 1.71e-3 | -3.111 | 1.86e-3 | 0.116 |
| White blood cell leukocyte count | 0.008 | 1.69e-3 | 4.458 | 8.29e-6 | 5.14e-4*** |
| ***Liver*** | | | | | |
| Albumin | 1.47e-3 | 1.75e-3 | 0.841 | 0.400 | 1.000 |
| Alkaline phosphatase | -2.75e-3 | 1.68e-3 | -1.642 | 0.101 | 1.000 |
| Alanine aminotransferase | -3.89e-4 | 1.63e-3 | -0.238 | 0.812 | 1.000 |
| Aspartate aminotransferase | 0.006 | 1.66e-3 | 3.586 | 3.36e-4 | 0.021* |
| Direct bilirubin | -0.005 | 1.80e-3 | -3.039 | 2.37e-3 | 0.147 |
| Gamma glutamyltransferase | -9.07e-4 | 1.66e-3 | -0.546 | 0.585 | 1.000 |
| Total bilirubin | -3.15e-3 | 1.66e-3 | -1.899 | 0.058 | 1.000 |
| ***Renal*** | | | | | |
| Creatinine | -0.012 | 1.39e-3 | -8.887 | 6.33e-19 | 3.93e-17*** |
| Cystatin C | -0.007 | 1.56e-3 | -4.750 | 2.03e-6 | 1.26e-4*** |
| Phosphate | 0.007 | 1.72e-3 | 3.791 | 1.50e-4 | 0.009** |
| Total protein | 4.38e-3 | 1.78e-3 | 2.460 | 0.014 | 0.861 |
| Urate | -4.08e-3 | 1.45e-3 | -2.818 | 4.83e-3 | 0.299 |
| Urea | -4.16e-3 | 1.64e-3 | -2.539 | 0.011 | 0.689 |
| ***Cardiovascular*** | | | | | |
| Apolipoprotein A | 3.62e-3 | 1.64e-3 | 2.206 | 0.027 | 1.000 |
| Apolipoprotein B | -4.15e-5 | 1.70e-3 | -0.024 | 0.981 | 1.000 |
| Cholesterol | 2.76e-3 | 1.67e-3 | 1.652 | 0.099 | 1.000 |
| LDL direct | 1.15e-3 | 1.69e-3 | 0.677 | 0.498 | 1.000 |
| Lipoprotein A | -1.70e-3 | 1.91e-3 | -0.890 | 0.374 | 1.000 |
| ***Other Non-BBB-Permeable*** | | | | | |
| Calcium | -1.58e-3 | 1.77e-3 | -0.893 | 0.372 | 1.000 |
| Glycated hemoglobin | -1.86e-3 | 1.66e-3 | -1.124 | 0.261 | 1.000 |
| Rheumatoid factor | -3.40e-3 | 1.72e-3 | -1.974 | 0.048 | 1.000 |

**Table S28.** Peripheral marker minimally adjusted associations with polygenic risk scores (PRS) at threshold ≤ 0.05 for bipolar disorder. BBB, blood-brain barrier; CRP, C-reactive protein; HDL, high-density lipoprotein; IGF-1, insulin-like growth factor 1; LDL, low-density lipoprotein; NLR, neutrophil-to-lymphocyte ratio; p-uncorr., p-uncorrected value; p-corr., Bonferroni p-corrected value; S.E., standard error; SHBG, sex hormone binding globulin. ***p corr. ≤ 0.001, **p corr. ≤ 0.01, *p corr. ≤ 0.05

| **Biomarker** | **Effect Size (β)** | **S.E.** | **t statistic** | **p-uncorr.** | **p-corr.** |
| --- | --- | --- | --- | --- | --- |
| ***BBB-Permeable*** | | | | | |
| Estradiol | 2.98e-3 | 1.61e-3 | 1.847 | 0.065 | 1.000 |
| Glucose | -3.92e-3 | 1.78e-3 | -2.198 | 0.028 | 1.000 |
| HDL cholesterol | 0.008 | 1.63e-3 | 5.039 | 4.67e-7 | 2.90e-5*** |
| IGF-1 | -2.77e-4 | 1.66e-3 | -0.167 | 0.868 | 1.000 |
| SHBG | 1.37e-3 | 1.65e-3 | 0.834 | 0.404 | 1.000 |
| Testosterone | 6.68e-4 | 7.92e-4 | 0.844 | 0.399 | 1.000 |
| Triglycerides | -1.31e-3 | 1.68e-3 | -0.778 | 0.437 | 1.000 |
| Vitamin D | -0.006 | 1.75e-3 | -3.260 | 1.11e-3 | 0.069 |
| ***Inflammatory and Hematological*** | | | | | |
| Basophil count | -2.21e-3 | 1.68e-3 | -1.314 | 0.189 | 1.000 |
| Basophil percentage | -2.37e-3 | 1.71e-3 | -1.387 | 0.165 | 1.000 |
| CRP | 5.41e-4 | 1.72e-3 | 0.314 | 0.753 | 1.000 |
| Eosinophil count | 3.30e-3 | 1.70e-3 | 1.937 | 0.053 | 1.000 |
| Eosinophil percentage | 2.08e-3 | 1.70e-3 | 1.224 | 0.221 | 1.000 |
| Hematocrit percentage | 6.56e-4 | 1.39e-3 | 0.471 | 0.637 | 1.000 |
| Hemoglobin concentration | 1.24e-3 | 1.36e-3 | 0.915 | 0.360 | 1.000 |
| High light scatter reticulocyte count | -0.011 | 1.71e-3 | -6.556 | 5.54e-11 | 3.44e-9*** |
| High light scatter reticulocyte percentage | -0.011 | 1.72e-3 | -6.223 | 4.89e-10 | 3.03e-8** |
| Immature reticulocyte fraction | -0.008 | 1.71e-3 | -4.548 | 5.43e-6 | 3.36e-4*** |
| Lymphocyte count | 4.72e-3 | 1.70e-3 | 2.783 | 0.005 | 0.334 |
| Lymphocyte percentage | 1.20e-3 | 1.69e-3 | 0.711 | 0.477 | 1.000 |
| Mean corpuscular hemoglobin | 0.006 | 1.69e-3 | 3.562 | 3.67e-4 | 0.023* |
| Mean corpuscular hemoglobin concentration | 1.89e-3 | 1.69e-3 | 1.120 | 0.263 | 1.000 |
| Mean corpuscular volume | 0.006 | 1.70e-3 | 3.449 | 5.62e-4 | 0.035* |
| Mean platelet thrombocyte volume | 1.61e-3 | 1.71e-3 | 0.944 | 0.345 | 1.000 |
| Mean reticulocyte volume | 1.99e-3 | 1.71e-3 | 1.165 | 0.244 | 1.000 |
| Mean sphered cell volume | 0.007 | 1.71e-3 | 3.879 | 1.05e-4 | 0.006** |
| Monocyte count | -4.28e-4 | 1.65e-3 | -0.259 | 0.796 | 1.000 |
| Monocyte percentage | -4.17e-3 | 1.65e-3 | -2.522 | 0.012 | 0.724 |
| Neutrophil count | 2.77e-3 | 1.71e-3 | 1.627 | 0.104 | 1.000 |
| Neutrophil percentage | 1.29e-4 | 1.71e-3 | 0.076 | 0.940 | 1.000 |
| NLR | -7.77e-4 | 1.70e-3 | -0.457 | 0.647 | 1.000 |
| Nucleated red blood cell count | 2.13e-3 | 1.71e-3 | 1.245 | 0.213 | 1.000 |
| Nucleated red blood cell percentage | 2.80e-3 | 1.71e-3 | 1.632 | 0.103 | 1.000 |
| Platelet count | -2.52e-3 | 1.65e-3 | -1.532 | 0.126 | 1.000 |
| Platelet crit | -2.12e-3 | 1.62e-3 | -1.315 | 0.189 | 1.000 |
| Platelet distribution width | 4.38e-4 | 1.69e-3 | 0.259 | 0.796 | 1.000 |
| Red blood cell erythrocyte count | -2.40e-3 | 1.46e-3 | -1.638 | 0.101 | 1.000 |
| Red blood cell erythrocyte distribution width | -4.15e-3 | 1.70e-3 | -2.435 | 0.015 | 0.924 |
| Reticulocyte count | -0.010 | 1.70e-3 | -5.896 | 3.73e-9 | 2.31e-7*** |
| Reticulocyte percentage | -0.010 | 1.72e-3 | -5.755 | 8.68e-9 | 5.38e-7*** |
| White blood cell leukocyte count | 3.96e-3 | 1.70e-3 | 2.324 | 0.020 | 1.000 |
| ***Liver*** | | | | | |
| Albumin | 1.92e-3 | 1.76e-3 | 1.088 | 0.277 | 1.000 |
| Alkaline phosphatase | -0.006 | 1.69e-3 | -3.558 | 3.73e-4 | 0.023* |
| Alanine aminotransferase | -3.02e-3 | 1.64e-3 | -1.837 | 0.066 | 1.000 |
| Aspartate aminotransferase | 0.005 | 1.68e-3 | 3.252 | 1.14e-3 | 0.071 |
| Direct bilirubin | -0.005 | 1.82e-3 | -2.798 | 0.005 | 0.319 |
| Gamma glutamyltransferase | -3.93e-4 | 1.67e-3 | -0.235 | 0.815 | 1.000 |
| Total bilirubin | -3.37e-3 | 1.67e-3 | -2.018 | 0.044 | 1.000 |
| ***Renal*** | | | | | |
| Creatinine | -0.011 | 1.40e-3 | -7.948 | 1.91e-15 | 1.18e-13*** |
| Cystatin C | -0.009 | 1.58e-3 | -5.920 | 3.22e-9 | 2.00e-7*** |
| Phosphate | 0.009 | 1.74e-3 | 4.921 | 8.60e-7 | 5.33e-5*** |
| Total protein | 3.49e-3 | 1.79e-3 | 1.945 | 0.052 | 1.000 |
| Urate | -0.005 | 1.46e-3 | -3.628 | 2.86e-4 | 0.018* |
| Urea | -2.81e-3 | 1.65e-3 | -1.704 | 0.088 | 1.000 |
| ***Cardiovascular*** | | | | | |
| Apolipoprotein A | 0.006 | 1.66e-3 | 3.511 | 4.46e-4 | 0.028* |
| Apolipoprotein B | -6.08e-4 | 1.72e-3 | -0.355 | 0.723 | 1.000 |
| Cholesterol | 3.38e-3 | 1.68e-3 | 2.012 | 0.044 | 1.000 |
| LDL direct | 1.26e-3 | 1.71e-3 | 0.738 | 0.461 | 1.000 |
| Lipoprotein A | -1.89e-3 | 1.93e-3 | -0.979 | 0.328 | 1.000 |
| ***Other Non-BBB-Permeable*** | | | | | |
| Calcium | -2.58e-3 | 1.79e-3 | -1.445 | 0.149 | 1.000 |
| Glycated hemoglobin | -5.94e-4 | 1.67e-3 | -0.356 | 0.722 | 1.000 |
| Rheumatoid factor | -3.12e-3 | 1.73e-3 | -1.799 | 0.072 | 1.000 |

**Table S29.** Peripheral marker minimally adjusted associations with polygenic risk scores (PRS) at threshold ≤ 0.1 for bipolar disorder. BBB, blood-brain barrier; CRP, C-reactive protein; HDL, high-density lipoprotein; IGF-1, insulin-like growth factor 1; LDL, low-density lipoprotein; NLR, neutrophil-to-lymphocyte ratio; p-uncorr., p-uncorrected value; p-corr., Bonferroni p-corrected value; S.E., standard error; SHBG, sex hormone binding globulin. ***p corr. ≤ 0.001, **p corr. ≤ 0.01, *p corr. ≤ 0.05

| **Biomarker** | **Effect Size (β)** | **S.E.** | **t statistic** | **p-uncorr.** | **p-corr.** |
| --- | --- | --- | --- | --- | --- |
| ***BBB-Permeable*** | | | | | |
| Estradiol | 2.58e-3 | 1.62e-3 | 1.593 | 0.111 | 1.000 |
| Glucose | -4.44e-3 | 1.80e-3 | -2.471 | 0.013 | 0.834 |
| HDL cholesterol | 0.008 | 1.64e-3 | 4.803 | 1.57e-6 | 9.71e-5*** |
| IGF-1 | 1.53e-4 | 1.67e-3 | 0.091 | 0.927 | 1.000 |
| SHBG | 1.51e-3 | 1.66e-3 | 0.908 | 0.364 | 1.000 |
| Testosterone | -1.09e-4 | 7.97e-4 | -0.137 | 0.891 | 1.000 |
| Triglycerides | -1.06e-3 | 1.69e-3 | -0.628 | 0.530 | 1.000 |
| Vitamin D | -0.005 | 1.76e-3 | -3.012 | 2.60e-3 | 0.161 |
| ***Inflammatory and Hematological*** | | | | | |
| Basophil count | -1.99e-3 | 1.69e-3 | -1.174 | 0.240 | 1.000 |
| Basophil percentage | -1.47e-3 | 1.72e-3 | -0.855 | 0.393 | 1.000 |
| CRP | -1.06e-3 | 1.74e-3 | -0.613 | 0.540 | 1.000 |
| Eosinophil count | 4.04e-3 | 1.72e-3 | 2.357 | 0.018 | 1.000 |
| Eosinophil percentage | 2.25e-3 | 1.72e-3 | 1.310 | 0.190 | 1.000 |
| Hematocrit percentage | 2.68e-5 | 1.40e-3 | 0.019 | 0.985 | 1.000 |
| Hemoglobin concentration | 4.98e-4 | 1.37e-3 | 0.365 | 0.715 | 1.000 |
| High light scatter reticulocyte count | -0.009 | 1.72e-3 | -5.398 | 6.74e-8 | 4.18e-6*** |
| High light scatter reticulocyte percentage | -0.009 | 1.73e-3 | -5.054 | 4.33e-7 | 2.69e-5*** |
| Immature reticulocyte fraction | -0.006 | 1.72e-3 | -3.317 | 9.10e-4 | 0.056 |
| Lymphocyte count | 0.007 | 1.71e-3 | 4.172 | 3.02e-5 | 1.87e-3** |
| Lymphocyte percentage | 2.70e-3 | 1.70e-3 | 1.586 | 0.113 | 1.000 |
| Mean corpuscular hemoglobin | 4.52e-3 | 1.70e-3 | 2.654 | 0.008 | 0.493 |
| Mean corpuscular hemoglobin concentration | 1.49e-3 | 1.70e-3 | 0.877 | 0.381 | 1.000 |
| Mean corpuscular volume | 4.59e-3 | 1.71e-3 | 2.688 | 0.007 | 0.446 |
| Mean platelet thrombocyte volume | -7.36e-4 | 1.72e-3 | -0.429 | 0.668 | 1.000 |
| Mean reticulocyte volume | 2.52e-3 | 1.72e-3 | 1.466 | 0.143 | 1.000 |
| Mean sphered cell volume | 0.006 | 1.72e-3 | 3.314 | 9.21e-4 | 0.057 |
| Monocyte count | 1.34e-3 | 1.67e-3 | 0.802 | 0.423 | 1.000 |
| Monocyte percentage | -3.67e-3 | 1.67e-3 | -2.203 | 0.028 | 1.000 |
| Neutrophil count | 2.81e-3 | 1.72e-3 | 1.638 | 0.101 | 1.000 |
| Neutrophil percentage | -1.53e-3 | 1.72e-3 | -0.893 | 0.372 | 1.000 |
| NLR | -2.44e-3 | 1.71e-3 | -1.425 | 0.154 | 1.000 |
| Nucleated red blood cell count | 8.00e-4 | 1.73e-3 | 0.464 | 0.643 | 1.000 |
| Nucleated red blood cell percentage | 1.26e-3 | 1.73e-3 | 0.733 | 0.464 | 1.000 |
| Platelet count | -2.13e-3 | 1.66e-3 | -1.286 | 0.198 | 1.000 |
| Platelet crit | -2.97e-3 | 1.63e-3 | -1.825 | 0.068 | 1.000 |
| Platelet distribution width | 1.28e-3 | 1.70e-3 | 0.753 | 0.451 | 1.000 |
| Red blood cell erythrocyte count | -2.21e-3 | 1.47e-3 | -1.503 | 0.133 | 1.000 |
| Red blood cell erythrocyte distribution width | -0.005 | 1.72e-3 | -3.055 | 2.25e-3 | 0.139 |
| Reticulocyte count | -0.008 | 1.71e-3 | -4.925 | 8.45e-7 | 5.24e-5*** |
| Reticulocyte percentage | -0.008 | 1.73e-3 | -4.806 | 1.54e-6 | 9.55e-5*** |
| White blood cell leukocyte count | 4.86e-3 | 1.71e-3 | 2.833 | 4.62e-3 | 0.286 |
| ***Liver*** | | | | | |
| Albumin | 1.28e-3 | 1.78e-3 | 0.720 | 0.471 | 1.000 |
| Alkaline phosphatase | -0.007 | 1.70e-3 | -4.041 | 5.31e-5 | 3.30e-3** |
| Alanine aminotransferase | -2.31e-3 | 1.66e-3 | -1.394 | 0.163 | 1.000 |
| Aspartate aminotransferase | 0.006 | 1.69e-3 | 3.400 | 6.74e-4 | 0.042* |
| Direct bilirubin | -4.85e-3 | 1.83e-3 | -2.653 | 0.008 | 0.495 |
| Gamma glutamyltransferase | -1.19e-3 | 1.68e-3 | -0.708 | 0.479 | 1.000 |
| Total bilirubin | -3.78e-3 | 1.68e-3 | -2.245 | 0.025 | 1.000 |
| ***Renal*** | | | | | |
| Creatinine | -0.011 | 1.41e-3 | -7.512 | 5.85e-14 | 3.63e-12*** |
| Cystatin C | -0.011 | 1.59e-3 | -6.792 | 1.11e-11 | 6.86e-10*** |
| Phosphate | 0.009 | 1.75e-3 | 5.115 | 3.13e-7 | 1.94e-5*** |
| Total protein | 1.52e-3 | 1.81e-3 | 0.840 | 0.401 | 1.000 |
| Urate | -0.005 | 1.47e-3 | -3.630 | 2.84e-4 | 0.018* |
| Urea | -2.81e-3 | 1.66e-3 | -1.694 | 0.090 | 1.000 |
| ***Cardiovascular*** | | | | | |
| Apolipoprotein A | 0.005 | 1.67e-3 | 3.169 | 1.53e-3 | 0.095 |
| Apolipoprotein B | -9.92e-4 | 1.73e-3 | -0.574 | 0.566 | 1.000 |
| Cholesterol | 2.95e-3 | 1.69e-3 | 1.741 | 0.082 | 1.000 |
| LDL direct | 7.26e-4 | 1.72e-3 | 0.422 | 0.673 | 1.000 |
| Lipoprotein A | -2.15e-3 | 1.94e-3 | -1.104 | 0.269 | 1.000 |
| ***Other Non-BBB-Permeable*** | | | | | |
| Calcium | -3.80e-3 | 1.80e-3 | -2.110 | 0.035 | 1.000 |
| Glycated hemoglobin | 3.71e-4 | 1.68e-3 | 0.221 | 0.825 | 1.000 |
| Rheumatoid factor | -3.13e-3 | 1.75e-3 | -1.791 | 0.073 | 1.000 |

**Table S30.** Peripheral marker minimally adjusted associations with polygenic risk scores (PRS) at threshold ≤ 1.0 for bipolar disorder. BBB, blood-brain barrier; CRP, C-reactive protein; HDL, high-density lipoprotein; IGF-1, insulin-like growth factor 1; LDL, low-density lipoprotein; NLR, neutrophil-to-lymphocyte ratio; p-uncorr., p-uncorrected value; p-corr., Bonferroni p-corrected value; S.E., standard error; SHBG, sex hormone binding globulin. ***p corr. ≤ 0.001, **p corr. ≤ 0.01, *p corr. ≤ 0.05

| **Biomarker** | **Effect Size (β)** | **S.E.** | **t statistic** | **p-uncorr.** | **p-corr.** |
| --- | --- | --- | --- | --- | --- |
| ***BBB-Permeable*** |  |  |  |  |  |
| Estradiol | 2.05e-3 | 1.65e-3 | 1.247 | 0.212 | 1.000 |
| Glucose | -3.25e-3 | 1.82e-3 | -1.781 | 0.075 | 1.000 |
| HDL cholesterol | 0.010 | 1.67e-3 | 5.854 | 4.81e-9 | 2.98e-7*** |
| IGF-1 | -3.12e-4 | 1.70e-3 | -0.184 | 0.854 | 1.000 |
| SHBG | 3.71e-3 | 1.69e-3 | 2.203 | 0.028 | 1.000 |
| Testosterone | -4.93e-4 | 8.09e-4 | -0.609 | 0.542 | 1.000 |
| Triglycerides | -1.60e-3 | 1.72e-3 | -0.932 | 0.351 | 1.000 |
| Vitamin D | -0.006 | 1.79e-3 | -3.287 | 1.01e-3 | 0.063 |
| ***Inflammatory and Hematological*** |  |  |  |  |  |
| Basophil count | -2.19e-3 | 1.72e-3 | -1.270 | 0.204 | 1.000 |
| Basophil percentage | -1.12e-3 | 1.75e-3 | -0.639 | 0.523 | 1.000 |
| CRP | -2.55e-3 | 1.76e-3 | -1.448 | 0.148 | 1.000 |
| Eosinophil count | 3.18e-3 | 1.74e-3 | 1.824 | 0.068 | 1.000 |
| Eosinophil percentage | 1.95e-3 | 1.74e-3 | 1.120 | 0.263 | 1.000 |
| Hematocrit percentage | -1.90e-3 | 1.42e-3 | -1.332 | 0.183 | 1.000 |
| Hemoglobin concentration | -1.14e-3 | 1.39e-3 | -0.824 | 0.410 | 1.000 |
| High light scatter reticulocyte count | -0.010 | 1.75e-3 | -5.996 | 2.03e-9 | 1.26e-7*** |
| High light scatter reticulocyte percentage | -0.010 | 1.76e-3 | -5.555 | 2.77e-8 | 1.72e-6*** |
| Immature reticulocyte fraction | -0.006 | 1.75e-3 | -3.585 | 3.37e-4 | 0.021* |
| Lymphocyte count | 0.006 | 1.73e-3 | 3.207 | 1.34e-3 | 0.083 |
| Lymphocyte percentage | 2.89e-3 | 1.73e-3 | 1.670 | 0.095 | 1.000 |
| Mean corpuscular hemoglobin | 0.007 | 1.73e-3 | 3.863 | 1.12e-4 | 0.007** |
| Mean corpuscular hemoglobin concentration | 1.73e-3 | 1.73e-3 | 1.003 | 0.316 | 1.000 |
| Mean corpuscular volume | 0.007 | 1.73e-3 | 3.883 | 1.03e-4 | 0.006** |
| Mean platelet thrombocyte volume | -2.31e-3 | 1.74e-3 | -1.324 | 0.186 | 1.000 |
| Mean reticulocyte volume | 2.88e-3 | 1.75e-3 | 1.649 | 0.099 | 1.000 |
| Mean sphered cell volume | 0.007 | 1.75e-3 | 4.125 | 3.71e-5 | 2.30e-3** |
| Monocyte count | 1.55e-3 | 1.69e-3 | 0.915 | 0.360 | 1.000 |
| Monocyte percentage | -1.25e-3 | 1.69e-3 | -0.738 | 0.461 | 1.000 |
| Neutrophil count | 1.26e-3 | 1.74e-3 | 0.721 | 0.471 | 1.000 |
| Neutrophil percentage | -2.43e-3 | 1.74e-3 | -1.391 | 0.164 | 1.000 |
| NLR | -2.50e-3 | 1.74e-3 | -1.442 | 0.149 | 1.000 |
| Nucleated red blood cell count | -5.21e-4 | 1.75e-3 | -0.298 | 0.766 | 1.000 |
| Nucleated red blood cell percentage | -3.89e-4 | 1.75e-3 | -0.222 | 0.824 | 1.000 |
| Platelet count | -1.78e-3 | 1.68e-3 | -1.061 | 0.289 | 1.000 |
| Platelet crit | -3.28e-3 | 1.65e-3 | -1.990 | 0.047 | 1.000 |
| Platelet distribution width | -8.02e-4 | 1.73e-3 | -0.464 | 0.643 | 1.000 |
| Red blood cell erythrocyte count | -0.005 | 1.50e-3 | -3.368 | 7.56e-4 | 0.047* |
| Red blood cell erythrocyte distribution width | -4.37e-3 | 1.74e-3 | -2.508 | 0.012 | 0.753 |
| Reticulocyte count | -0.010 | 1.74e-3 | -5.682 | 1.33e-8 | 8.24e-7*** |
| Reticulocyte percentage | -0.009 | 1.76e-3 | -5.079 | 3.80e-7 | 2.36e-5*** |
| White blood cell leukocyte count | 3.03e-3 | 1.74e-3 | 1.741 | 0.082 | 1.000 |
| ***Liver*** |  |  |  |  |  |
| Albumin | -5.28e-5 | 1.81e-3 | -0.029 | 0.977 | 1.000 |
| Alkaline phosphatase | -0.007 | 1.73e-3 | -4.196 | 2.72e-5 | 1.69e-3** |
| Alanine aminotransferase | -1.94e-3 | 1.68e-3 | -1.153 | 0.249 | 1.000 |
| Aspartate aminotransferase | 0.006 | 1.71e-3 | 3.522 | 4.29e-4 | 0.027* |
| Direct bilirubin | -1.79e-3 | 1.86e-3 | -0.963 | 0.335 | 1.000 |
| Gamma glutamyltransferase | -1.28e-3 | 1.71e-3 | -0.748 | 0.454 | 1.000 |
| Total bilirubin | -1.06e-3 | 1.71e-3 | -0.618 | 0.536 | 1.000 |
| ***Renal*** |  |  |  |  |  |
| Creatinine | -0.012 | 1.43e-3 | -8.656 | 4.89e-18 | 3.03e-16*** |
| Cystatin C | -0.014 | 1.61e-3 | -8.411 | 4.07e-17 | 2.52e-15*** |
| Phosphate | 0.009 | 1.78e-3 | 5.126 | 2.97e-7 | 1.84e-5*** |
| Total protein | -2.15e-3 | 1.83e-3 | -1.173 | 0.241 | 1.000 |
| Urate | -0.006 | 1.49e-3 | -4.241 | 2.23e-5 | 1.38e-3** |
| Urea | -0.005 | 1.69e-3 | -3.234 | 1.22e-3 | 0.076 |
| ***Cardiovascular*** |  |  |  |  |  |
| Apolipoprotein A | 0.006 | 1.69e-3 | 3.593 | 3.27e-4 | 0.020* |
| Apolipoprotein B | -8.81e-4 | 1.75e-3 | -0.502 | 0.615 | 1.000 |
| Cholesterol | 3.27e-3 | 1.72e-3 | 1.903 | 0.057 | 1.000 |
| LDL direct | 5.87e-4 | 1.74e-3 | 0.336 | 0.737 | 1.000 |
| Lipoprotein A | 6.01e-4 | 1.97e-3 | 0.305 | 0.761 | 1.000 |
| ***Other Non-BBB-Permeable*** |  |  |  |  |  |
| Calcium | -0.006 | 1.83e-3 | -3.267 | 1.09e-3 | 0.067 |
| Glycated hemoglobin | 3.40e-4 | 1.71e-3 | 0.199 | 0.842 | 1.000 |
| Rheumatoid factor | -1.40e-3 | 1.77e-3 | -0.789 | 0.430 | 1.000 |

**Table S31.** Peripheral marker fully adjusted associations with polygenic risk scores (PRS) at threshold ≤ 0.01 for bipolar disorder. BBB, blood-brain barrier; CRP, C-reactive protein; HDL, high-density lipoprotein; IGF-1, insulin-like growth factor 1; LDL, low-density lipoprotein; NLR, neutrophil-to-lymphocyte ratio; p-uncorr., p-uncorrected value; p-corr., Bonferroni p-corrected value; S.E., standard error; SHBG, sex hormone binding globulin. ***p corr. ≤ 0.001, **p corr. ≤ 0.01, *p corr. ≤ 0.05

| **Biomarker** | **Effect Size (β)** | **S.E.** | **t statistic** | **p-uncorr.** | **p-corr.** |
| --- | --- | --- | --- | --- | --- |
| ***BBB-Permeable*** | | | | | |
| Estradiol | 2.27e-3 | 1.60e-3 | 1.416 | 0.157 | 1.000 |
| Glucose | -3.89e-3 | 1.75e-3 | -2.223 | 0.026 | 1.000 |
| HDL cholesterol | 3.82e-3 | 1.50e-3 | 2.538 | 0.011 | 0.691 |
| IGF-1 | -1.83e-3 | 1.63e-3 | -1.118 | 0.264 | 1.000 |
| SHBG | -1.38e-3 | 1.52e-3 | -0.905 | 0.366 | 1.000 |
| Testosterone | -8.09e-6 | 7.74e-4 | -0.010 | 0.992 | 1.000 |
| Triglycerides | 3.90e-3 | 1.59e-3 | 2.443 | 0.015 | 0.902 |
| Vitamin D | -0.007 | 1.71e-3 | -4.315 | 1.60e-5 | 9.89e-4*** |
| ***Inflammatory and Hematological*** | | | | | |
| Basophil count | -1.01e-3 | 1.66e-3 | -0.605 | 0.545 | 1.000 |
| Basophil percentage | -3.61e-3 | 1.70e-3 | -2.125 | 0.034 | 1.000 |
| CRP | 1.07e-3 | 1.60e-3 | 0.665 | 0.506 | 1.000 |
| Eosinophil count | 2.06e-3 | 1.68e-3 | 1.225 | 0.221 | 1.000 |
| Eosinophil percentage | 1.69e-4 | 1.69e-3 | 0.100 | 0.921 | 1.000 |
| Hematocrit percentage | 2.29e-3 | 1.37e-3 | 1.670 | 0.095 | 1.000 |
| Hemoglobin concentration | 2.85e-3 | 1.34e-3 | 2.132 | 0.033 | 1.000 |
| High light scatter reticulocyte count | -2.66e-3 | 1.55e-3 | -1.715 | 0.086 | 1.000 |
| High light scatter reticulocyte percentage | -2.83e-3 | 1.57e-3 | -1.800 | 0.072 | 1.000 |
| Immature reticulocyte fraction | -2.13e-3 | 1.62e-3 | -1.318 | 0.187 | 1.000 |
| Lymphocyte count | 0.006 | 1.63e-3 | 3.905 | 9.44e-5 | 0.006** |
| Lymphocyte percentage | 1.95e-4 | 1.68e-3 | 0.116 | 0.907 | 1.000 |
| Mean corpuscular hemoglobin | 2.83e-3 | 1.65e-3 | 1.713 | 0.087 | 1.000 |
| Mean corpuscular hemoglobin concentration | 2.37e-3 | 1.68e-3 | 1.413 | 0.158 | 1.000 |
| Mean corpuscular volume | 2.15e-3 | 1.65e-3 | 1.306 | 0.192 | 1.000 |
| Mean platelet thrombocyte volume | -4.22e-3 | 1.70e-3 | -2.487 | 0.013 | 0.799 |
| Mean reticulocyte volume | -6.74e-4 | 1.69e-3 | -0.399 | 0.690 | 1.000 |
| Mean sphered cell volume | 3.02e-3 | 1.66e-3 | 1.818 | 0.069 | 1.000 |
| Monocyte count | 2.42e-3 | 1.62e-3 | 1.491 | 0.136 | 1.000 |
| Monocyte percentage | -3.43e-3 | 1.64e-3 | -2.092 | 0.036 | 1.000 |
| Neutrophil count | 0.006 | 1.64e-3 | 3.696 | 2.19e-4 | 0.014* |
| Neutrophil percentage | 1.26e-3 | 1.70e-3 | 0.745 | 0.456 | 1.000 |
| NLR | 4.44e-4 | 1.69e-3 | 0.263 | 0.793 | 1.000 |
| Nucleated red blood cell count | 1.48e-3 | 1.70e-3 | 0.869 | 0.385 | 1.000 |
| Nucleated red blood cell percentage | 6.20e-4 | 1.70e-3 | 0.364 | 0.716 | 1.000 |
| Platelet count | 3.20e-3 | 1.63e-3 | 1.962 | 0.050 | 1.000 |
| Platelet crit | 1.32e-3 | 1.60e-3 | 0.829 | 0.407 | 1.000 |
| Platelet distribution width | -2.91e-3 | 1.68e-3 | -1.731 | 0.083 | 1.000 |
| Red blood cell erythrocyte count | 6.90e-4 | 1.43e-3 | 0.483 | 0.629 | 1.000 |
| Red blood cell erythrocyte distribution width | -4.25e-3 | 1.68e-3 | -2.531 | 0.011 | 0.706 |
| Reticulocyte count | -2.10e-3 | 1.57e-3 | -1.338 | 0.181 | 1.000 |
| Reticulocyte percentage | -2.59e-3 | 1.60e-3 | -1.613 | 0.107 | 1.000 |
| White blood cell leukocyte count | 0.007 | 1.61e-3 | 4.474 | 7.69e-6 | 4.77e-4*** |
| ***Liver*** | | | | | |
| Albumin | 9.73e-4 | 1.73e-3 | 0.562 | 0.574 | 1.000 |
| Alkaline phosphatase | -2.25e-3 | 1.65e-3 | -1.369 | 0.171 | 1.000 |
| Alanine aminotransferase | 2.16e-3 | 1.57e-3 | 1.374 | 0.170 | 1.000 |
| Aspartate aminotransferase | 0.007 | 1.65e-3 | 4.365 | 1.27e-5 | 7.87e-4*** |
| Direct bilirubin | -0.005 | 1.80e-3 | -2.876 | 4.02e-3 | 0.249 |
| Gamma glutamyltransferase | 3.46e-4 | 1.63e-3 | 0.213 | 0.831 | 1.000 |
| Total bilirubin | -2.76e-3 | 1.65e-3 | -1.676 | 0.094 | 1.000 |
| ***Renal*** | | | | | |
| Creatinine | -0.011 | 1.38e-3 | -8.060 | 7.65e-16 | 4.74e-14*** |
| Cystatin C | -0.006 | 1.47e-3 | -4.419 | 9.92e-6 | 6.15e-4*** |
| Phosphate | 0.005 | 1.72e-3 | 3.114 | 1.84e-3 | 0.114 |
| Total protein | 0.005 | 1.78e-3 | 3.022 | 2.51e-3 | 0.156 |
| Urate | -1.49e-3 | 1.34e-3 | -1.112 | 0.266 | 1.000 |
| Urea | -2.57e-3 | 1.63e-3 | -1.579 | 0.114 | 1.000 |
| ***Cardiovascular*** | | | | | |
| Apolipoprotein A | 2.50e-3 | 1.57e-3 | 1.590 | 0.112 | 1.000 |
| Apolipoprotein B | 9.40e-4 | 1.70e-3 | 0.554 | 0.580 | 1.000 |
| Cholesterol | 3.12e-3 | 1.67e-3 | 1.870 | 0.061 | 1.000 |
| LDL direct | 1.96e-3 | 1.69e-3 | 1.156 | 0.248 | 1.000 |
| Lipoprotein A | -1.77e-3 | 1.92e-3 | -0.922 | 0.357 | 1.000 |
| ***Other Non-BBB-Permeable*** | | | | | |
| Calcium | -1.81e-3 | 1.78e-3 | -1.022 | 0.307 | 1.000 |
| Glycated hemoglobin | -8.36e-4 | 1.60e-3 | -0.523 | 0.601 | 1.000 |
| Rheumatoid factor | -3.45e-3 | 1.72e-3 | -2.000 | 0.045 | 1.000 |

**Table S32.** Peripheral marker fully adjusted associations with polygenic risk scores (PRS) at threshold ≤ 0.05 for bipolar disorder. BBB, blood-brain barrier; CRP, C-reactive protein; HDL, high-density lipoprotein; IGF-1, insulin-like growth factor 1; LDL, low-density lipoprotein; NLR, neutrophil-to-lymphocyte ratio; p-uncorr., p-uncorrected value; p-corr., Bonferroni p-corrected value; S.E., standard error; SHBG, sex hormone binding globulin. ***p corr. ≤ 0.001, **p corr. ≤ 0.01, *p corr. ≤ 0.05

| **Biomarker** | **Effect Size (β)** | **S.E.** | **t statistic** | **p-uncorr.** | **p-corr.** |
| --- | --- | --- | --- | --- | --- |
| ***BBB-Permeable*** | | | | | |
| Estradiol | 2.80e-3 | 1.61e-3 | 1.737 | 0.082 | 1.000 |
| Glucose | -2.68e-3 | 1.76e-3 | -1.520 | 0.129 | 1.000 |
| HDL cholesterol | 0.006 | 1.52e-3 | 4.107 | 4.01e-5 | 2.49e-3** |
| IGF-1 | -6.10e-4 | 1.65e-3 | -0.371 | 0.711 | 1.000 |
| SHBG | -1.73e-3 | 1.53e-3 | -1.128 | 0.259 | 1.000 |
| Testosterone | 2.57e-5 | 7.79e-4 | 0.033 | 0.974 | 1.000 |
| Triglycerides | 5.01e-4 | 1.61e-3 | 0.312 | 0.755 | 1.000 |
| Vitamin D | -0.007 | 1.72e-3 | -3.903 | 9.50e-5 | 0.006** |
| ***Inflammatory and Hematological*** | | | | | |
| Basophil count | -2.61e-3 | 1.68e-3 | -1.556 | 0.120 | 1.000 |
| Basophil percentage | -2.72e-3 | 1.71e-3 | -1.592 | 0.111 | 1.000 |
| CRP | 2.25e-3 | 1.61e-3 | 1.397 | 0.162 | 1.000 |
| Eosinophil count | 3.24e-3 | 1.69e-3 | 1.913 | 0.056 | 1.000 |
| Eosinophil percentage | 2.11e-3 | 1.71e-3 | 1.237 | 0.216 | 1.000 |
| Hematocrit percentage | 1.70e-3 | 1.38e-3 | 1.232 | 0.218 | 1.000 |
| Hemoglobin concentration | 2.31e-3 | 1.35e-3 | 1.716 | 0.086 | 1.000 |
| High light scatter reticulocyte count | -0.008 | 1.56e-3 | -4.988 | 6.10e-7 | 3.78e-5*** |
| High light scatter reticulocyte percentage | -0.008 | 1.58e-3 | -4.848 | 1.25e-6 | 7.75e-5*** |
| Immature reticulocyte fraction | -0.006 | 1.63e-3 | -3.534 | 4.10e-4 | 0.025* |
| Lymphocyte count | 4.58e-3 | 1.65e-3 | 2.783 | 0.005 | 0.334 |
| Lymphocyte percentage | 1.27e-3 | 1.69e-3 | 0.752 | 0.452 | 1.000 |
| Mean corpuscular hemoglobin | 3.93e-3 | 1.66e-3 | 2.366 | 0.018 | 1.000 |
| Mean corpuscular hemoglobin concentration | 1.97e-3 | 1.69e-3 | 1.166 | 0.244 | 1.000 |
| Mean corpuscular volume | 3.49e-3 | 1.66e-3 | 2.106 | 0.035 | 1.000 |
| Mean platelet thrombocyte volume | 1.64e-3 | 1.71e-3 | 0.962 | 0.336 | 1.000 |
| Mean reticulocyte volume | 5.93e-4 | 1.70e-3 | 0.348 | 0.728 | 1.000 |
| Mean sphered cell volume | 4.01e-3 | 1.68e-3 | 2.392 | 0.017 | 1.000 |
| Monocyte count | -2.43e-4 | 1.63e-3 | -0.149 | 0.882 | 1.000 |
| Monocyte percentage | -3.79e-3 | 1.65e-3 | -2.297 | 0.022 | 1.000 |
| Neutrophil count | 2.48e-3 | 1.65e-3 | 1.502 | 0.133 | 1.000 |
| Neutrophil percentage | -2.03e-5 | 1.71e-3 | -0.012 | 0.991 | 1.000 |
| NLR | -1.04e-3 | 1.70e-3 | -0.610 | 0.542 | 1.000 |
| Nucleated red blood cell count | 2.24e-3 | 1.72e-3 | 1.304 | 0.192 | 1.000 |
| Nucleated red blood cell percentage | 2.95e-3 | 1.72e-3 | 1.719 | 0.086 | 1.000 |
| Platelet count | -2.58e-3 | 1.64e-3 | -1.571 | 0.116 | 1.000 |
| Platelet crit | -2.18e-3 | 1.61e-3 | -1.356 | 0.175 | 1.000 |
| Platelet distribution width | 1.01e-3 | 1.69e-3 | 0.598 | 0.550 | 1.000 |
| Red blood cell erythrocyte count | -1.95e-4 | 1.44e-3 | -0.135 | 0.892 | 1.000 |
| Red blood cell erythrocyte distribution width | -3.94e-3 | 1.69e-3 | -2.329 | 0.020 | 1.000 |
| Reticulocyte count | -0.007 | 1.58e-3 | -4.289 | 1.80e-5 | 1.11e-3** |
| Reticulocyte percentage | -0.007 | 1.62e-3 | -4.362 | 1.29e-5 | 8.00e-4*** |
| White blood cell leukocyte count | 3.66e-3 | 1.62e-3 | 2.261 | 0.024 | 1.000 |
| ***Liver*** | | | | | |
| Albumin | 1.30e-3 | 1.74e-3 | 0.747 | 0.455 | 1.000 |
| Alkaline phosphatase | -0.005 | 1.66e-3 | -3.264 | 1.10e-3 | 0.068 |
| Alanine aminotransferase | -5.07e-4 | 1.58e-3 | -0.320 | 0.749 | 1.000 |
| Aspartate aminotransferase | 0.007 | 1.67e-3 | 4.011 | 6.04e-5 | 3.74e-3** |
| Direct bilirubin | -4.81e-3 | 1.81e-3 | -2.655 | 0.008 | 0.491 |
| Gamma glutamyltransferase | 7.55e-4 | 1.64e-3 | 0.461 | 0.645 | 1.000 |
| Total bilirubin | -2.96e-3 | 1.66e-3 | -1.783 | 0.075 | 1.000 |
| ***Renal*** | | | | | |
| Creatinine | -0.010 | 1.39e-3 | -7.040 | 1.93e-12 | 1.20e-10*** |
| Cystatin C | -0.008 | 1.48e-3 | -5.565 | 2.62e-8 | 1.63e-6*** |
| Phosphate | 0.007 | 1.73e-3 | 4.097 | 4.18e-5 | 2.59e-3** |
| Total protein | 4.51e-3 | 1.79e-3 | 2.517 | 0.012 | 0.733 |
| Urate | -2.65e-3 | 1.35e-3 | -1.962 | 0.050 | 1.000 |
| Urea | -1.28e-3 | 1.64e-3 | -0.780 | 0.435 | 1.000 |
| ***Cardiovascular*** | | | | | |
| Apolipoprotein A | 4.48e-3 | 1.58e-3 | 2.827 | 4.69e-3 | 0.291 |
| Apolipoprotein B | 5.16e-4 | 1.71e-3 | 0.301 | 0.763 | 1.000 |
| Cholesterol | 3.82e-3 | 1.68e-3 | 2.273 | 0.023 | 1.000 |
| LDL direct | 2.20e-3 | 1.71e-3 | 1.292 | 0.196 | 1.000 |
| Lipoprotein A | -1.90e-3 | 1.93e-3 | -0.983 | 0.326 | 1.000 |
| ***Other Non-BBB-Permeable*** | | | | | |
| Calcium | -2.89e-3 | 1.79e-3 | -1.614 | 0.107 | 1.000 |
| Glycated hemoglobin | 4.36e-4 | 1.61e-3 | 0.271 | 0.787 | 1.000 |
| Rheumatoid factor | -3.37e-3 | 1.74e-3 | -1.942 | 0.052 | 1.000 |

**Table S33.** Peripheral marker fully adjusted associations with polygenic risk scores (PRS) at threshold ≤ 0.1 for bipolar disorder. BBB, blood-brain barrier; CRP, C-reactive protein; HDL, high-density lipoprotein; IGF-1, insulin-like growth factor 1; LDL, low-density lipoprotein; NLR, neutrophil-to-lymphocyte ratio; p-uncorr., p-uncorrected value; p-corr., Bonferroni p-corrected value; S.E., standard error; SHBG, sex hormone binding globulin. ***p corr. ≤ 0.001, **p corr. ≤ 0.01, *p corr. ≤ 0.05

| **Biomarker** | **Effect Size (β)** | **S.E.** | **t statistic** | **p-uncorr.** | **p-corr.** |
| --- | --- | --- | --- | --- | --- |
| ***BBB-Permeable*** | | | | | |
| Estradiol | 2.43e-3 | 1.62e-3 | 1.497 | 0.135 | 1.000 |
| Glucose | -3.24e-3 | 1.77e-3 | -1.828 | 0.068 | 1.000 |
| HDL cholesterol | 0.006 | 1.53e-3 | 3.963 | 7.40e-5 | 4.59e-3** |
| IGF-1 | -1.54e-4 | 1.66e-3 | -0.093 | 0.926 | 1.000 |
| SHBG | -1.44e-3 | 1.54e-3 | -0.935 | 0.350 | 1.000 |
| Testosterone | -6.41e-4 | 7.85e-4 | -0.817 | 0.414 | 1.000 |
| Triglycerides | 5.61e-4 | 1.62e-3 | 0.347 | 0.729 | 1.000 |
| Vitamin D | -0.006 | 1.73e-3 | -3.580 | 3.43e-4 | 0.021* |
| ***Inflammatory and Hematological*** | | | | | |
| Basophil count | -2.40e-3 | 1.69e-3 | -1.420 | 0.155 | 1.000 |
| Basophil percentage | -1.87e-3 | 1.72e-3 | -1.086 | 0.277 | 1.000 |
| CRP | 4.55e-4 | 1.63e-3 | 0.280 | 0.780 | 1.000 |
| Eosinophil count | 3.91e-3 | 1.71e-3 | 2.294 | 0.022 | 1.000 |
| Eosinophil percentage | 2.24e-3 | 1.72e-3 | 1.304 | 0.192 | 1.000 |
| Hematocrit percentage | 9.84e-4 | 1.39e-3 | 0.707 | 0.479 | 1.000 |
| Hemoglobin concentration | 1.50e-3 | 1.36e-3 | 1.104 | 0.269 | 1.000 |
| High light scatter reticulocyte count | -0.006 | 1.57e-3 | -3.851 | 1.18e-4 | 0.007** |
| High light scatter reticulocyte percentage | -0.006 | 1.60e-3 | -3.713 | 2.05e-4 | 0.013* |
| Immature reticulocyte fraction | -3.89e-3 | 1.64e-3 | -2.376 | 0.018 | 1.000 |
| Lymphocyte count | 0.007 | 1.66e-3 | 4.181 | 2.90e-5 | 1.80e-3** |
| Lymphocyte percentage | 2.79e-3 | 1.70e-3 | 1.640 | 0.101 | 1.000 |
| Mean corpuscular hemoglobin | 2.37e-3 | 1.67e-3 | 1.420 | 0.156 | 1.000 |
| Mean corpuscular hemoglobin concentration | 1.64e-3 | 1.70e-3 | 0.963 | 0.335 | 1.000 |
| Mean corpuscular volume | 2.16e-3 | 1.67e-3 | 1.294 | 0.196 | 1.000 |
| Mean platelet thrombocyte volume | -6.69e-4 | 1.72e-3 | -0.389 | 0.697 | 1.000 |
| Mean reticulocyte volume | 1.01e-3 | 1.71e-3 | 0.590 | 0.555 | 1.000 |
| Mean sphered cell volume | 3.01e-3 | 1.69e-3 | 1.785 | 0.074 | 1.000 |
| Monocyte count | 1.43e-3 | 1.65e-3 | 0.868 | 0.385 | 1.000 |
| Monocyte percentage | -3.30e-3 | 1.66e-3 | -1.985 | 0.047 | 1.000 |
| Neutrophil count | 2.43e-3 | 1.66e-3 | 1.460 | 0.144 | 1.000 |
| Neutrophil percentage | -1.70e-3 | 1.72e-3 | -0.990 | 0.322 | 1.000 |
| NLR | -2.72e-3 | 1.71e-3 | -1.587 | 0.112 | 1.000 |
| Nucleated red blood cell count | 1.01e-3 | 1.73e-3 | 0.587 | 0.557 | 1.000 |
| Nucleated red blood cell percentage | 1.55e-3 | 1.73e-3 | 0.897 | 0.369 | 1.000 |
| Platelet count | -2.20e-3 | 1.66e-3 | -1.329 | 0.184 | 1.000 |
| Platelet crit | -3.03e-3 | 1.62e-3 | -1.867 | 0.062 | 1.000 |
| Platelet distribution width | 1.88e-3 | 1.70e-3 | 1.103 | 0.270 | 1.000 |
| Red blood cell erythrocyte count | -6.20e-5 | 1.45e-3 | -0.043 | 0.966 | 1.000 |
| Red blood cell erythrocyte distribution width | -0.005 | 1.70e-3 | -2.978 | 2.90e-3 | 0.180 |
| Reticulocyte count | -0.005 | 1.59e-3 | -3.366 | 7.62e-4 | 0.047* |
| Reticulocyte percentage | -0.006 | 1.63e-3 | -3.458 | 5.45e-4 | 0.034* |
| White blood cell leukocyte count | 4.46e-3 | 1.63e-3 | 2.736 | 0.006 | 0.385 |
| ***Liver*** | | | | | |
| Albumin | 7.33e-4 | 1.76e-3 | 0.417 | 0.676 | 1.000 |
| Alkaline phosphatase | -0.006 | 1.67e-3 | -3.789 | 1.52e-4 | 0.009** |
| Alanine aminotransferase | 3.97e-5 | 1.59e-3 | 0.025 | 0.980 | 1.000 |
| Aspartate aminotransferase | 0.007 | 1.68e-3 | 4.087 | 4.37e-5 | 2.71e-3** |
| Direct bilirubin | -4.53e-3 | 1.82e-3 | -2.484 | 0.013 | 0.806 |
| Gamma glutamyltransferase | -2.11e-4 | 1.65e-3 | -0.128 | 0.898 | 1.000 |
| Total bilirubin | -3.27e-3 | 1.67e-3 | -1.956 | 0.050 | 1.000 |
| ***Renal*** | | | | | |
| Creatinine | -0.009 | 1.40e-3 | -6.623 | 3.51e-11 | 2.18e-9*** |
| Cystatin C | -0.010 | 1.49e-3 | -6.580 | 4.71e-11 | 2.92e-9*** |
| Phosphate | 0.008 | 1.74e-3 | 4.328 | 1.50e-5 | 9.32e-4*** |
| Total protein | 2.58e-3 | 1.80e-3 | 1.430 | 0.153 | 1.000 |
| Urate | -2.94e-3 | 1.36e-3 | -2.155 | 0.031 | 1.000 |
| Urea | -1.41e-3 | 1.65e-3 | -0.855 | 0.392 | 1.000 |
| ***Cardiovascular*** | | | | | |
| Apolipoprotein A | 4.08e-3 | 1.60e-3 | 2.556 | 0.011 | 0.656 |
| Apolipoprotein B | 8.34e-5 | 1.72e-3 | 0.048 | 0.961 | 1.000 |
| Cholesterol | 3.41e-3 | 1.69e-3 | 2.013 | 0.044 | 1.000 |
| LDL direct | 1.65e-3 | 1.72e-3 | 0.957 | 0.339 | 1.000 |
| Lipoprotein A | -2.18e-3 | 1.95e-3 | -1.119 | 0.263 | 1.000 |
| ***Other Non-BBB-Permeable*** | | | | | |
| Calcium | -4.09e-3 | 1.80e-3 | -2.270 | 0.023 | 1.000 |
| Glycated hemoglobin | 1.12e-3 | 1.62e-3 | 0.692 | 0.489 | 1.000 |
| Rheumatoid factor | -3.36e-3 | 1.75e-3 | -1.918 | 0.055 | 1.000 |

**Table S34.** Peripheral marker fully adjusted associations with polygenic risk scores (PRS) at threshold ≤ 1.0 for bipolar disorder. BBB, blood-brain barrier; CRP, C-reactive protein; HDL, high-density lipoprotein; IGF-1, insulin-like growth factor 1; LDL, low-density lipoprotein; NLR, neutrophil-to-lymphocyte ratio; p-uncorr., p-uncorrected value; p-corr., Bonferroni p-corrected value; S.E., standard error; SHBG, sex hormone binding globulin. ***p corr. ≤ 0.001, **p corr. ≤ 0.01, *p corr. ≤ 0.05

| **Biomarker** | **Effect Size (β)** | **S.E.** | **t statistic** | **p-uncorr.** | **p-corr.** |
| --- | --- | --- | --- | --- | --- |
| ***BBB-Permeable*** | | | | | |
| Estradiol | 1.90e-3 | 1.65e-3 | 1.149 | 0.250 | 1.000 |
| Glucose | -1.51e-3 | 1.80e-3 | -0.840 | 0.401 | 1.000 |
| HDL cholesterol | 0.007 | 1.55e-3 | 4.209 | 2.57e-5 | 1.59e-3** |
| IGF-1 | -1.06e-3 | 1.68e-3 | -0.628 | 0.530 | 1.000 |
| SHBG | -5.40e-4 | 1.57e-3 | -0.344 | 0.730 | 1.000 |
| Testosterone | -1.35e-3 | 7.97e-4 | -1.700 | 0.089 | 1.000 |
| Triglycerides | 1.08e-3 | 1.64e-3 | 0.660 | 0.510 | 1.000 |
| Vitamin D | -0.007 | 1.76e-3 | -4.175 | 2.97e-5 | 1.84e-3** |
| ***Inflammatory and Hematological*** | | | | | |
| Basophil count | -2.46e-3 | 1.71e-3 | -1.436 | 0.151 | 1.000 |
| Basophil percentage | -1.48e-3 | 1.75e-3 | -0.848 | 0.396 | 1.000 |
| CRP | 2.41e-4 | 1.65e-3 | 0.146 | 0.884 | 1.000 |
| Eosinophil count | 3.47e-3 | 1.73e-3 | 2.003 | 0.045 | 1.000 |
| Eosinophil percentage | 2.19e-3 | 1.74e-3 | 1.254 | 0.210 | 1.000 |
| Hematocrit percentage | -6.79e-4 | 1.41e-3 | -0.481 | 0.631 | 1.000 |
| Hemoglobin concentration | 1.05e-4 | 1.38e-3 | 0.076 | 0.939 | 1.000 |
| High light scatter reticulocyte count | -0.006 | 1.59e-3 | -3.708 | 2.09e-4 | 0.013* |
| High light scatter reticulocyte percentage | -0.006 | 1.62e-3 | -3.489 | 4.85e-4 | 0.030* |
| Immature reticulocyte fraction | -3.43e-3 | 1.66e-3 | -2.061 | 0.039 | 1.000 |
| Lymphocyte count | 0.006 | 1.68e-3 | 3.484 | 4.93e-4 | 0.031* |
| Lymphocyte percentage | 3.06e-3 | 1.73e-3 | 1.770 | 0.077 | 1.000 |
| Mean corpuscular hemoglobin | 3.92e-3 | 1.70e-3 | 2.306 | 0.021 | 1.000 |
| Mean corpuscular hemoglobin concentration | 1.83e-3 | 1.73e-3 | 1.057 | 0.291 | 1.000 |
| Mean corpuscular volume | 3.60e-3 | 1.70e-3 | 2.121 | 0.034 | 1.000 |
| Mean platelet thrombocyte volume | -2.16e-3 | 1.75e-3 | -1.234 | 0.217 | 1.000 |
| Mean reticulocyte volume | 1.08e-3 | 1.74e-3 | 0.618 | 0.537 | 1.000 |
| Mean sphered cell volume | 3.71e-3 | 1.71e-3 | 2.169 | 0.030 | 1.000 |
| Monocyte count | 1.95e-3 | 1.67e-3 | 1.170 | 0.242 | 1.000 |
| Monocyte percentage | -9.81e-4 | 1.69e-3 | -0.581 | 0.561 | 1.000 |
| Neutrophil count | 1.21e-3 | 1.69e-3 | 0.718 | 0.473 | 1.000 |
| Neutrophil percentage | -2.70e-3 | 1.75e-3 | -1.546 | 0.122 | 1.000 |
| NLR | -2.94e-3 | 1.74e-3 | -1.691 | 0.091 | 1.000 |
| Nucleated red blood cell count | -3.08e-4 | 1.75e-3 | -0.175 | 0.861 | 1.000 |
| Nucleated red blood cell percentage | -9.10e-5 | 1.75e-3 | -0.052 | 0.959 | 1.000 |
| Platelet count | -1.67e-3 | 1.68e-3 | -0.995 | 0.320 | 1.000 |
| Platelet crit | -3.07e-3 | 1.65e-3 | -1.863 | 0.062 | 1.000 |
| Platelet distribution width | -1.06e-4 | 1.73e-3 | -0.062 | 0.951 | 1.000 |
| Red blood cell erythrocyte count | -2.27e-3 | 1.47e-3 | -1.540 | 0.123 | 1.000 |
| Red blood cell erythrocyte distribution width | -3.92e-3 | 1.73e-3 | -2.265 | 0.024 | 1.000 |
| Reticulocyte count | -0.006 | 1.62e-3 | -3.429 | 6.07e-4 | 0.038* |
| Reticulocyte percentage | -0.005 | 1.65e-3 | -3.058 | 2.23e-3 | 0.138 |
| White blood cell leukocyte count | 3.13e-3 | 1.65e-3 | 1.891 | 0.059 | 1.000 |
| ***Liver*** | | | | | |
| Albumin | -1.25e-3 | 1.79e-3 | -0.700 | 0.484 | 1.000 |
| Alkaline phosphatase | -0.006 | 1.70e-3 | -3.607 | 3.10e-4 | 0.019* |
| Alanine aminotransferase | 1.47e-3 | 1.62e-3 | 0.909 | 0.364 | 1.000 |
| Aspartate aminotransferase | 0.008 | 1.70e-3 | 4.452 | 8.50e-6 | 5.27e-4*** |
| Direct bilirubin | -1.65e-3 | 1.85e-3 | -0.889 | 0.374 | 1.000 |
| Gamma glutamyltransferase | 3.47e-4 | 1.67e-3 | 0.207 | 0.836 | 1.000 |
| Total bilirubin | -8.16e-4 | 1.70e-3 | -0.481 | 0.631 | 1.000 |
| ***Renal*** | | | | | |
| Creatinine | -0.011 | 1.42e-3 | -7.517 | 5.61e-14 | 3.48e-12*** |
| Cystatin C | -0.012 | 1.51e-3 | -7.622 | 2.51e-14 | 1.56e-12*** |
| Phosphate | 0.007 | 1.77e-3 | 4.148 | 3.36e-5 | 2.08e-3** |
| Total protein | -1.02e-3 | 1.83e-3 | -0.556 | 0.578 | 1.000 |
| Urate | -2.64e-3 | 1.38e-3 | -1.906 | 0.057 | 1.000 |
| Urea | -3.76e-3 | 1.68e-3 | -2.236 | 0.025 | 1.000 |
| ***Cardiovascular*** | | | | | |
| Apolipoprotein A | 3.78e-3 | 1.62e-3 | 2.334 | 0.020 | 1.000 |
| Apolipoprotein B | 4.69e-4 | 1.75e-3 | 0.268 | 0.789 | 1.000 |
| Cholesterol | 3.58e-3 | 1.72e-3 | 2.083 | 0.037 | 1.000 |
| LDL direct | 1.58e-3 | 1.75e-3 | 0.906 | 0.365 | 1.000 |
| Lipoprotein A | 5.94e-4 | 1.98e-3 | 0.301 | 0.764 | 1.000 |
| ***Other Non-BBB-Permeable*** | | | | | |
| Calcium | -0.006 | 1.83e-3 | -3.516 | 4.38e-4 | 0.027* |
| Glycated hemoglobin | 1.94e-3 | 1.65e-3 | 1.182 | 0.237 | 1.000 |
| Rheumatoid factor | -1.69e-3 | 1.78e-3 | -0.954 | 0.340 | 1.000 |
